# Sleep Health Behind Bars: A Global Scoping Review of Sleep in Carceral Settings

**DOI:** 10.1101/2025.10.08.25337513

**Authors:** Victoria Hummel, Jerimiah Kouka, Patrick Li, Alyssa Grimshaw, Chante-Colleen Lewis, Ella Suh, Gabriel Paglia, Cassandra Michel, Johanna E. Elumn

**Author notes:** Corresponding Author Johanna E. Elumn, Ph.D, M.S.W. SEICHE Center for Health and Justice Department of Internal Medicine, Yale University School of Medicine, New Haven, Connecticut, 60 Temple Street, New Haven, CT 06515, USA.

## Abstract

**Background:** Sleep is a fundamental determinant of health, yet little is known about sleep in carceral environments where structural, environmental, and psychosocial factors uniquely disrupt rest. Incarcerated individuals face disproportionately high burdens of illness, with sleep disturbances contributing to these disparities. Understanding the evidence on sleep in incarceration is essential for informing care, policy, and research.

**Methods:** We conducted a scoping review of peer-reviewed literature on sleep in incarcerated populations, following the Joanna Briggs Institute Manual and PRISMA-ScR guidelines. Ten databases were searched from inception to March 2025 for studies addressing sleep among currently incarcerated individuals. Two reviewers screened and extracted data, with quality appraisal adapted from Joanna Briggs Institute tools.

**Results:** Of 5,518 records identified, 63 studies published between 1978 and 2025 met inclusion criteria. Most were observational (75%), cross-sectional (59%), and published after 2014, with studies conducted mostly outside the United States. Research clustered into three domains: (1) descriptions of sleep quality and insomnia; (2) sleep and health outcomes, particularly depression, anxiety, PTSD, aggression, and suicidality, with limited study of cardiometabolic risk; and (3) interventions including cognitive behavioral therapy for insomnia, mindfulness, yoga, and relaxation, which showed feasibility but were limited by small samples, lack of controls, or short follow-up. Objective measures were rarely used.

**Conclusions:** Evidence demonstrates compromised sleep health in carceral settings due to psychiatric comorbidity, environmental stressors, and restricted care. Gaps remain in physical health outcomes, women’s experiences, and post-release effects. Interventions tailored to carceral realities and adapted evidence-based interventions showed promise.

## Introduction

The incarceration rate in the United States is the highest in the world, with about two million people behind bars and 11 million people cycling in and out of carceral facilities annually (Fair & Walmsley, 2024). While incarcerated people have a constitutional right to healthcare (“Estelle v. Gamble”, 1976), the reality within carceral facilities often creates barriers to receiving it, especially for sleep-related issues. These limitations exist in environments that are stressful, with aspects of the social and physical environment that adversely impact sleep. The lack of research into the sleep health of currently and formerly incarcerated individuals results in a limited understanding of how these environments aRect sleep, feasible interventions for sleep issues in highly restrictive environments, and the broader impact on health outcomes, including mental health and cardiovascular disease risk. Just as food, water, air, and shelter are necessary for human survival, so too is sleep a basic physiological need (Maslow, 1943; NHLBI-NIH, 2022). Poor sleep quality and sleep deprivation are known to negatively impact growth, immune health, memory, mood, cardiovascular health, hormone regulation, metabolism, and more (Lloyd-Jones et al., 2022; NHLBI-NIH, 2022; Thompson et al., 2022). Both acute total and chronic partial sleep deprivation have both been shown to impair cognitive function (Alhola & Polo-Kantola, 2007; Belenky et al., 2003; Thompson et al., 2022). While minor sleep deprivation in the modern workplace has become banal, extreme forms of sleep deprivation nonetheless constitute cruel and unusual punishment and torture (Cakal, 2021; Jaech, 2022). Long sleep, defined as sleeping greater than nine hours per night, similarly constitutes a slew of negative health measures ranging from depression to impaired fertility and higher all-cause mortality (Jike et al., 2018; Medic et al., 2017; Osmun, 2023).

Incarcerated people may have unique *individual-level* reasons for being sleep deficient. For instance, incarcerated people have higher rates of mood disorders and chronic pain, which amplify sleep deficiency, compared with those who have never been incarcerated (Herrero Babiloni et al., 2020; Kundermann et al., 2004; Williams et al., 2014). Sleep deficiency, in return, aRects stress, anger, and suicidality, which can further influence. Qualitative exploration of trauma of incarceration and sleep shows that fear of violence, untreated health conditions, and lack of access to appropriate treatment all contribute to pervasive sleep problems during incarceration and persisting after (Elumn et al., 2024).

Within carceral settings where artificial lights are on day and night, and violence, noise, and surveillance abound, sleep may be especially fraught (Dewa et al., 2015; Elumn et al., 2024). Life within carceral facilities is often marked by little autonomy over one’s sleep environment, daily schedule, and medical care, and is often the site of physical and emotional trauma, which can lead to sleep problems (Dewa et al., 2017; Elger, 2009; Elumn et al., 2024). In addition, limited access to physical activity, meal timing and content, boredom, and stress can all contribute to poor sleep quality, including not enough sleep, oversleeping, and sleep disturbance(Dewa et al., 2015).

Incarcerated individuals experience a disproportionately high prevalence of both mental and physical health disorders, with sleep disturbances exacerbating these conditions. Sleep health is intricately linked to cardiovascular, metabolic, and psychiatric outcomes, yet remains an underexplored aspect of incarceration-related health disparities. The Sleep Justice Study, a prospective cohort of individuals recently released from incarceration, assessed sleep using both subjective measures and actigraphy to examine associations with cardiometabolic risk factor control (Elumn et al., 2023). This work recognizes sleep as a determinant of cardiometabolic health in justice-involved populations and demonstrates the need for complementary reviews to map the current evidence base.

Despite the importance of sleep in maintaining good health and wellbeing, our review revealed a limited literature on experiences and health implications of sleep in carceral settings. Existing scoping and systematic reviews on sleep and incarceration have primarily focused on sleep quality or specific disorders such as insomnia, with some attention to sleep quality and mental health outcomes (Dewa et al., 2015; GriRiths & Hina, 2022; Sheppard & Hogan, 2022). Previous studies rarely focus on the connection between sleep and physical health conditions or the sleep.

Many health concerns are associated with being incarcerated, but while there is currently a growing body of literature on sleep and incarceration, these studies focused on sleep within prisons in the United States. This scoping review will examine existing literature on sleep during incarceration regardless of the region where the study took place. The articles highlighted focus on a range of topics including sleep quality, insomnia, and the role of environmental factors and mental health on sleep health.

In this scoping review of the literature, we focused on any article related to sleep, not just insomnia, to see what other types of sleep problems have been studied in this population and what interventions have been tested in carceral settings to address sleep. Our review is intended to orient future research on sleep in carceral settings and health care to improve the sleep health of people living in carceral facilities.

## Methods

This scoping review was conducted and reported in accordance with Levac et al’s recommendations for scoping review methodology and the JBI Manual for Evidence Synthesis, Preferred Reporting Items for Systematic Reviews and Meta-Analyses (PRISMA) 2020 and the PRISMA Extension for Scoping Reviews (Supplementary Table 1).(*JBI Manual for Evidence Synthesis*, 2020; Levac et al., 2010; Page et al., 2021; Tricco et al., 2018) The study protocol was published on Open Science Framework (osf.io/hn5tp/).

### Data Sources and Search Strategy

An exhaustive search of the literature was conducted by a medical research librarian (AAG) in Cochrane Library, Criminal Justice Abstracts, Ovid Embase, Google Scholar, Ovid Medline, National Criminal Justice Reference Services Abstracts, PsycExtra, PsycInfo, PubMed, Scopus, and Web of Science Core Collection databases to find relevant articles published from the inception of each database to March 5, 2025. Databases were searched using a combination of keywords and controlled vocabulary for justice involved individuals and sleep disturbances. The search was not limited by language, publication type, or year (full search strategies available in Supplementary Table 2). The search was peer-reviewed by a second medical librarian using the Peer Review of Electronic Search Strategies (PRESS).(McGowan et al., 2016) Forward and backward citation chasing was performed using CitationChaser to identify additional relevant studies not retrieved by the database search.(Haddaway et al., 2021)

### Eligibility criteria and Study Selection

Articles needed to be investigative and have a primary focus on sleep, sleep interventions, or include sleep as a secondary variable. Studies must be full articles published in peer reviewed journals between 1978 and 2025, written in English, involve human participants, and measure some aspect of sleep (e.g. duration, quality). The 1978 cut oR marks the year the U.S. Department of Health and Human Services (HHS) placed regulations on research involving prisoners, designating them as a vulnerable research population. Studies, however, were not limited to the U.S. Participants were required to be incarcerated during the duration of the study in a jail or prison. Forensic use of sleep as defense for a crime and studies on individuals in psychiatric hospitals and group homes were excluded. Papers were excluded if they were about the sleep of carceral staR.

Search results from all databases were imported into an Endnote 25 library. Duplicates were removed using the Yale Reference Deduplicator.(Yale University Harvey Cushing/John Hay Whitney Medical, 2021) The deduplicated results were then imported into Covidence, a systematic review software for screening and data extraction. Two independent screeners performed title/abstract review followed by full text review. Screening disagreements were resolved by a third investigator.

### Data Extraction and Risk of Bias

Data extraction was completed in Qualtrics using a standardized form to capture study characteristics (design, setting, sample size, participant demographics), sleep measures (Sylvia et al., 2021), exposures, and reported outcomes. Two reviewers independently extracted data and assessed study quality using criteria adapted from the Joanna Briggs Institute critical appraisal tools, with a third investigator reviewing and resolving discrepancies. Risk of bias domains included clarity of aims, appropriateness of methods, validity of sleep measures, and transparency of reporting, and extracted data were synthesized narratively according to scoping review methodology.

## Results

Database searches resulted in 5,518 citations (Figure 1). After removing duplicates, 3,342 citations underwent title and abstract screening. Of these, 330 citations met the criteria for full-text review. Subsequently, 62 studies met the inclusion criteria for the study. An additional study was found through citation chasing for a total of 63 manuscripts included. We excluded 268 studies from the search results and one study in citation chasing results due to the focus not being on sleep, wrong study population, non-peer reviewed publications, no original data, wrong study design, non-English language, wrong setting, sleep not discussed, year limitation, and wrong outcomes (Supplementary Table 3).

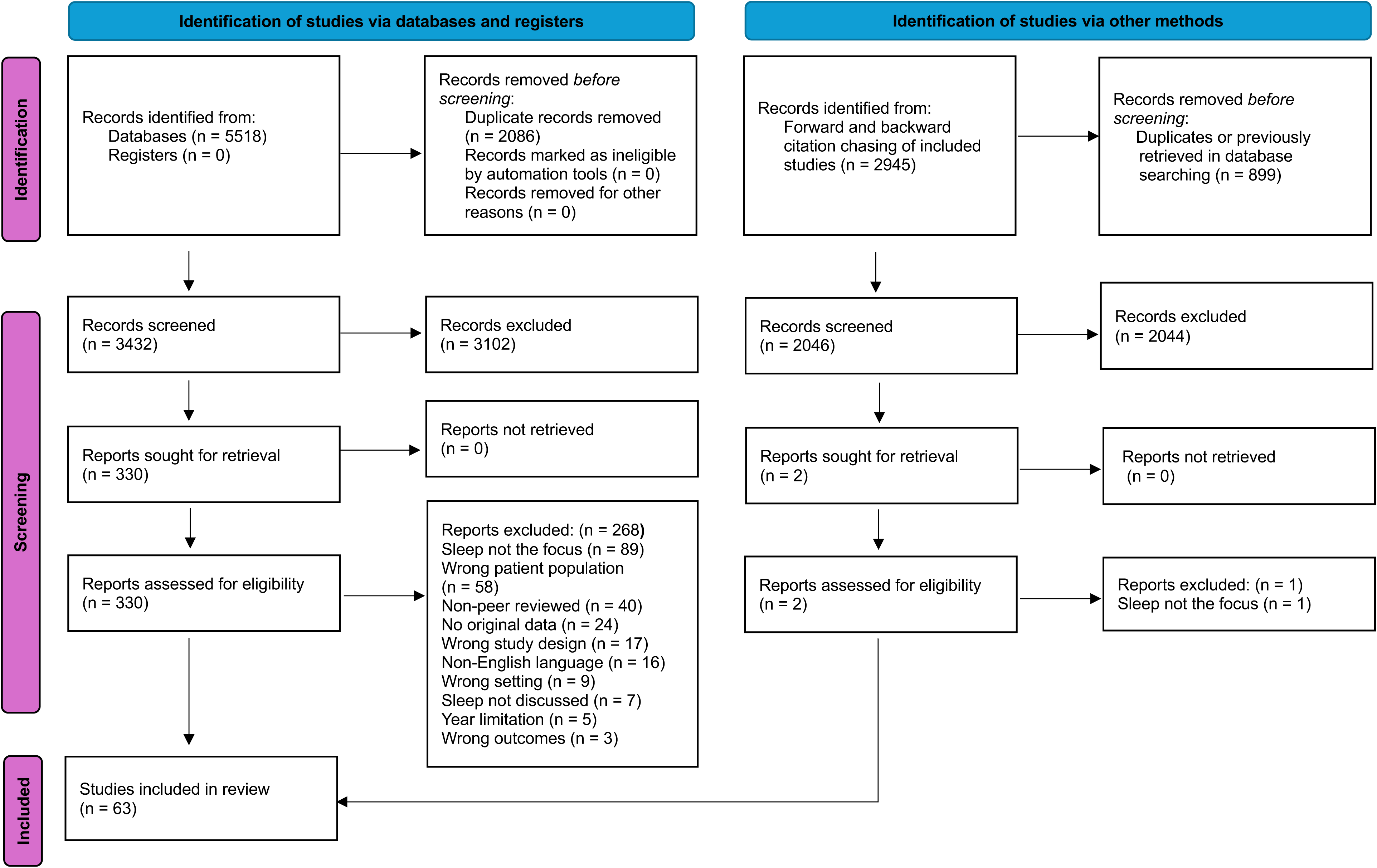

### Study Characteristics

All 63 of the studies included in this review were published between 1978 and 2025, with most occurring after 2014 (n=48), indicating that this is a growing body of research. Most of the studies were observational (75%). More than half of the studies were cross-sectional (59%), and randomized/quasi-randomized control trials made up 13% of the studies. Qualitative studies made up 6% of the studies included in the scoping review.

### Study Populations

The studies all took place among people who were currently incarcerated, with most taking place in prisons and the remainder taking place in a jail or carceral psychiatric unit. The studies were mostly conducted among adults (83%), 56% were among incarcerated males, 14% incarcerated females, and 27% of studies included both genders.

### Study Locations

The studies took place across the world, with 37% from Europe, 21% from North America (12 U.S. and 1 in Canada), 14% from Asia, 14% from the Middle East, 11% from Africa, and 1 study each in South America and New Zealand.

### Study focus

The studies described sleep among incarcerated people, including the quality, types of sleep problems, and causes. They fit into three main categories: 1) descriptive of sleep problems and causes, 2) sleep and associated health outcomes, and 4) interventions that assessed their impact on sleep.

## Sleep Quality and Insomnia Among Incarcerated Individuals

### Sleep Quality and Insomnia

The articles reviewed mainly focused on sleep quality and insomnia among incarcerated individuals, showing a consistent pattern of poor sleep and widespread sleep disturbances in this group. Goudard et al. (2017) carried out a study in a French prison using questionnaires and medical chart reviews to evaluate sleep quality, causes of sleep problems, and insomnia treatments. They found that 56% of participants were dissatisfied with their sleep, with 57% reporting that sleep issues began after incarceration and 31% experiencing a deterioration of pre-existing sleep problems. Hypnotic drugs were prescribed to 25% of the incarcerated individuals, mostly benzodiazepines (Adornetti et al., 2023), but only 42% believed these medications were effective. Similarly, Fakorede et al. (2023) reported a high rate of insomnia (45.7%) among 300 male inmates in a Nigerian maximum-security prison, which was closely linked to poor quality of life.

### Sleep and Circadian Patterns in Youth

Adornetti et al. (2023) explored sleep and circadian patterns in juveniles across 11 carceral facilities using various sleep measures, including the Sleep-Wake Behavior Problems Scale and a daily sleep diary. Despite the juveniles obtaining the recommended 8-10 hours of sleep on average, they took significantly longer to fall asleep (47 minutes compared to the recommended 10-20 minutes). This delay was attributed to environmental factors such as constant lighting and rigid facility schedules. The study’s recommendations included adjusting schedules and lighting conditions, providing eye shades, and offering sleep health education to staff.

### Environmental and Social Determinants of Sleep

Elger (2004) examined insomnia prevalence in Switzerland through a year-long study of medical consultations, finding that 44.3% of prisoners reported insomnia, often linked to psychiatric disorders such as depression and prison-related stress. In another intervention study, Elger (2003) tracked the sleep quality of 52 incarcerated individuals with insomnia treated with hypnotics over two months. Initial findings showed significant improvements in sleep quality, latency, and duration within the first two weeks, but many participants continued to experience clinically significant insomnia, underscoring the need for comprehensive mental health interventions.

Other studies provide broader insights into sleep issues in prisons. Geng et al. (2020) investigated the correlation between psychosocial factors and sleep patterns in a Chinese carceral setting. They found that although 47% of participants slept seven or more hours, poor sleep quality was reported by 45.9%. Insomnia was most prevalent among older individuals, those in poor physical health, and those with PTSD or depression.

However, one study by Camplain et. al (2022), showed that sleep improved in a carceral setting when examining sleep in relation to pre-carceral housing environments of incarcerated individuals while in a rural county jail in Flagstaff, Arizona. Using cross-sectional survey methods, 194 individuals incarcerated in jail reported sleep quality prior to and during incarceration (two question Likert scale) and pre-incarceration housing status. Results showed that individuals in non-permanent housing before incarceration were less likely to experience worsening sleep quality during incarceration, compared to those in permanent housing (Camplain et al., 2022).

### Objective Sleep Assessment

Methods involving actigraphy provided objective data on sleep-wake patterns. D’Aurizio et al. (2020) assessed sleep quality in Italian prisoners using actigraphy, comparing their sleep to a control group of non-incarcerated males. The prisoners exhibited poorer sleep quality, longer sleep latency, and shorter total sleep time compared to the control group, reinforcing the challenges of sleeping in a highly controlled environment.

### Summary

Collectively, these studies illustrate the significant issue of poor sleep quality and high insomnia rates among incarcerated individuals, with contributing factors including psychiatric disorders, environmental conditions, and pre-existing health issues. The findings suggest that improving carceral environments, along with expanding access to comprehensive mental health care and targeted insomnia treatments, is crucial for enhancing sleep quality in this population.

## Sleep and Health Outcomes

Mental health disorders such as depression, anxiety, post-traumatic stress disorder (PTSD), and personality disorders are significantly more prevalent among incarcerated individuals than in the general population. These conditions have direct effects on sleep quality, including nightmares and insomnia, and are exacerbated by associated behaviors such as substance use.

Among the studies reviewed, several articles addressed the relationship between sleep and health conditions, emphasizing the significant co-occurrence of sleep disturbances with depression, anxiety, PTSD, personality disorders, and substance use. Two studies examined sleep and cardiovascular disease risk.

### Depression and Anxiety

Several studies across diverse contexts consistently identify strong associations between sleep disturbances and depression, anxiety, and related mood disorders in incarcerated populations (Raha et al., 2018)(D’Aurizio et al., 2020; Zhang et al., 2023)(Carli et al., 2011; Oniszczenko & Romsicka, 2020; Vogler et al., 2014). Depression is especially prevalent among incarcerated women. Raha et al. (2018) identified that 85% of women at Guwahati Central Jail exhibit depressive symptoms, compared to 62.5% of men. Childhood trauma, pervasive in prison populations, is strongly associated with PTSD and depression (Carli et al., 2011). The compounding impact of incarceration, coupled with additional stressors like the COVID-19 pandemic (Zhang et al., 2023), exacerbates these conditions. D’Aurizio et al. (2020) reported that, in Italian prisoners, poor sleep quality corresponded with increased anxiety and mood disorders, highlighting the complex emotional regulation required by incarceration.

### Post-Traumatic Stress Disorder and Sleep Disturbances

Sleep disturbances are often early indicators of PTSD, with affected individuals experiencing chronic nightmares and various other sleep disorders (Acar et al., 2019; Dos Santos et al., 2017; Eichelman & Dorava, 2021; Eichelman & Dorava, 2022; Sylvia et al., 2021; Tussey et al., 2024; Woolhouse et al., 2018). Neurobiological findings suggest that disrupted REM sleep and heightened limbic activity contribute to PTSD in incarcerated populations (Acar et al., 2019). One study of women in a Wisconsin facility, found high incidences of traumatic dreams in incarcerated women, beyond the incidence of PTSD, indicating potential undertreatment of these symptoms by prison mental health professionals (Eichelman & Dorava, 2022).

Two subpopulations of incarcerated people stood out in the discussion connection between sleep and PTSD, veterans and women. Incarcerated veterans face unique mental health challenges, including high rates of PTSD, depression, and related sleep disturbances. They report more nightmares, insomnia, and sleep fragmentation compared to non-veterans, often due to pre-incarceration trauma and military stressors (Sylvia et al., 2021). The transition from military service to incarceration further heightens sleep-related health disparities.

Studies focusing on incarcerated women also reveal significant connections between trauma and sleep disturbances. A qualitative study in Rio De Janeiro found chronic insomnia, hypersomnia, and nightmares prevalent among incarcerated women and attributable to prior trauma and ongoing psychological distress, with some women using excessive sleep as a maladaptive coping mechanism (Dos Santos et al., 2017). Factors like depressive symptoms and PTSD were significantly associated with insomnia and poor sleep quality in a study of incarcerated women in Idaho with excessive noise, poor bedding, and mental health being commonly cited factors disrupting sleep (Tussey et al., 2024).

Additionally, women with traumatic brain injuries in New Zealand also reported poor sleep quality, stemming from both structural regulations and the prison environment (Woolhouse et al., 2018).

### Sleep Deprivation and Aggression

Sleep deprivation in prisons is linked to increased psychological distress and aggression, with findings emerging primarily from studies of men (Barker et al., 2016; Ireland & Culpin, 2006; Meijers et al., 2015; Vogler et al., 2014). In one of/the only study that used actigraphy, nocturnal restlessness was strongly correlated with daytime aggression, likely reflecting impaired inhibitory control from sleep loss (Meijers et al., 2015). Other work has shown that perceived poor sleep quality predicts aggressive behavior even when sleep duration does not (Barker et al., 2016), and that both short sleep and poor quality are associated with greater aggression, as well as co-occurring symptoms such as ADHD, rumination, and somatic complaints (Vogler et al., 2014). Among incarcerated adolescents and young adults (14-21 years), aggression was linked to both reduced sleep duration and poorer sleep quality, with heightened hostility particularly predictive of fewer hours of sleep and lower sleep quality (Ireland & Culpin, 2006).

### Substance Use

Substance use complicates the relationship between poor sleep and health outcomes in incarcerated populations with use as self-medication for sleep and as a contributor to poor sleep (Chinichian & Alemohammad, 2020; Cope, 2003; Haghighat & Tabatabaee, 2014).

Many incarcerated people use substances like cannabis to help them cope with sleep disturbances or to counteract boredom and inactivity, which can destabilize natural sleep patterns. Abrupt cessation of substance use upon incarceration further disrupts sleep, particularly in people dependent on substances like alcohol, where withdrawal is linked to higher rates of self-harm and aggression. Chinchian and Alemohammad (2020) reported that 87.7% of prisoners with substance use disorders experienced poor sleep quality, which was associated with total duration of imprisonment, the remainder of the prison term, history of sedative use, and poly-substance use in an Iranian prison.

### Personality Disorders

Some of the studies focused specifically on sleep and personality disorders, linking antisocial personality disorder (Oniszczenko & Romsicka, 2020) and borderline personality disorder (Harty et al., 2010) to distinct patterns of sleep disturbances. Lindberg et al. (2003) found that incarcerated individuals with ASP exhibited increased slow-wave sleep, particularly in the deepest stage (S4), suggesting a compensatory response to the intense psychological and social difficulties they face. Sleep disturbances in ASP also overlap with those commonly observed in BPD, including frequent awakenings and poor sleep efficiency (Harty et al., 2010). Despite these shared symptoms, sleep disruptions in ASP cannot be fully attributed to comorbid conditions such as depression or substance dependence, indicating that ASP itself plays a distinct role in altering sleep patterns.

### Sleep and Physical Health

Only two studies focus specifically on the relationship between sleep and physical health in incarcerated populations. In Ghana, a cardiovascular risk assessment reported an average sleep duration of 5.5 hours per night among incarcerated individuals, with nearly 90% having trouble sleeping (Abukari et al., 2024). These disturbances were associated with elevated rates of pre-hypertension (54.5%), dyslipidemia (48.1%), and overweight/obesity (24.6%). In Turkey, 43% of incarcerated men were found to have experienced poor sleep quality, which was significantly associated with obesity and cardiometabolic disease risk (Atan et al., 2025). While both studies demonstrate links between disrupted sleep and cardiometabolic vulnerability, the Ghana study’s reliance on self-reported sleep duration without a standardized instrument limits comparability with research using validated measures.

### Interventions

Several interventions sought to improve sleep through mind-body practices, including mindfulness and relaxation programs focused on improving sleep and reducing anxiety and depression. Five of these studies involved mindfulness techniques (An et al., 2019; Ferszt et al., 2015; Poorebrahim et al., 2022; Sylvia et al., 2021; Zhang et al., 2022), three focused on relaxation (Jacob & Sharma, 2019; Lutz, 1990; Toler, 1978), and two assessing yoga programs (Habibzadeh & Nosratabad, 2024; Kerekes et al., 2017). Across these studies, such approaches were generally effective in improving sleep and reducing anxiety and depression. A randomized control trial by Kerekes et al (2017) examined the outcomes of a 10-week yoga program in Sweden on participants mental health and sleep. Within group analysis showed some improvements in the PSQI global score for those who were in the yoga group and when compared to the control group, there was a close to significant (0.06) decrease in PSQI scores. Participants also showed improvements in the other outcomes measured including reduced perceived stress, increased psychological and emotional well-being, decreased aggressive and antisocial behavior, and improved attention and impulse control.

Two studies focused on assessing whether using physical activity could improve sleep (Adri et al., 2022; Johnson et al., 2019). One study by Adri et al. (2022) enrolled a small sample of adolescents evaluating whether individual and group-based activities improved sleep. They found that group games did improve sleep quality, however, the study only included three sessions over a one-week period. The authors posited that engaging with peers as part of physical activity may have improved sleep by reducing stress and improving mood.

Johnson et al. (2019) was the only intervention study to look at a physical health outcome, focusing on whether physical activity, screen time and sleep impacted body mass index (BMI). While they found no significant association between sleep duration and weight gain, long sleepers gained more weight per year.

Three studies in the United Kingdom looked at the treatment of insomnia with cognitive behavioral therapy (CBT-I) in carceral settings: a single-session CBT-I with self-management materials for men in prison (Randall et al., 2019); a stakeholder-informed design of a prison-specific stepped-care insomnia pathway (Dewa et al., 2018); and a feasibility evaluation of the stepped-care pathway that incorporates peer support, environmental aids, selective hypnotic prescribing, and CBT-I (Dewa et al., 2024).Collectively, these studies indicates that CBT-I-based approaches are feasible and acceptable in prisons and, in uncontrolled evaluations, are associated with clinically meaningful reductions in insomnia severity and improvements in mood symptoms.

In a single-session trial, Randall et al. (2019) observed short-term decreases in insomnia among men after one CBT-I session supplemented by a self-management pamphlet and optional weekly support sessions. This approach was designed to address insomnia in a setting where the carceral environment limits access to treatment. Among the limitations were the lack of a control group and the short follow-up period of the study. Dewa et al (2018) sought stakeholder feedback from prison staff, incarcerated people, and sleep researchers about the design of a treatment pathway for insomnia in prisons and building on this work, Dewa et al. (2024) found the stepped-care pathway to be feasible and acceptable in a high-secure prison, with 96.4% of participants reporting improvements in their sleep and also improved depression, anxiety, cognitive function, and general wellbeing. Both studies were done with men in prison and neither of the studies to test the intervention effectiveness used a control group. Testing these two approaches in different settings and with women is an important next step.

Each of these studies examines different types of interventions across various global settings. Collectively, they show that interventions in carceral environments—ranging from mindfulness and relaxation to yoga programs, physical activities, and prison-adapted CBT-I—are practical and produce short-term gains in self-reported sleep and mood. To identify what truly works for sleep problems, replicating studies in other settings with control groups is an essential next step. Since individual-level programs cannot fully address the structural causes of sleep loss in prisons, future efforts should combine behavioral strategies with environmental changes—such as noise and light reduction, schedule modifications—and medication stewardship policies, developed collaboratively with incarcerated peer facilitators and staff. Where sleep deprivation, whether intentional or unintentional, exists, prison leadership and staff must work to transform the facility culture to tackle the underlying causes and effects of poor sleep health.

## Discussion

Even in community settings, individuals often face challenges accessing specialized sleep care (Collen et al., 2020; Singh et al., 2023). These barriers are amplified in carceral systems, where access to primary care, specialty services, pharmacological treatments, and behavioral interventions is highly restricted. Evidence consistently shows that sleep health in these environments is compromised, supporting the need for interventions that simultaneously address environmental conditions (e.g., lights, noise, schedules, violence), access to medical care, and individual-level supports (D’Aurizio et al., 2023; Dewa et al., 2015; Sheppard & Hogan, 2022).

Most research on sleep in carceral settings has focused narrowly on insomnia and poor sleep quality, with limited attention to other clinically significant disorders such as sleep apnea, parasomnias, PTSD, and hypersomnolence. Mental health outcomes have been most consistently examined, particularly depression and anxiety, with some studies addressing aggression and suicidality, often among men, and personality disorders. Where women were included, samples were typically small. Only one study enrolled a larger women’s sample (Eichelman & Dorava, 2022), which limits precision and generalizability for women and gender-diverse people. The concentration on psychiatric correlates reflects both the high prevalence of mental illness in this population and the relative feasibility of assessing mental health outcomes through self-report and short-term designs.

By contrast, the relationship between sleep and physical health has been examined far less extensively. Although community-based data show that inadequate or poor-quality sleep is a determinant of obesity, hypertension, diabetes, cardiometabolic risk, and impair immune function (Itani et al., 2017; St-Onge et al., 2016), only a small subset of prison-based studies directly assesses these outcomes. This limited attention is significant given the elevated burden of chronic disease in incarcerated populations and the likelihood that carceral conditions exacerbate these risks. Sleep apnea, for example, is presumed to be common due to high rates of obesity, substance use, and psychiatric comorbidity, yet its long-term consequences in prisons and jails remain poorly characterized. The imbalance between mental and physical health research leaves substantial uncertainty about how sleep contributes to chronic disease disparities in justice-involved populations.

Across studies, environmental stressors – including noise, constant or bright night lighting, limited autonomy over routines, and safety concerns - were consistently identified as barriers to sleep and as constraints on the effectiveness of behavioral interventions (Dewa et al., 2017; Elumn et al., 2023; Sheppard & Hogan, 2022; Tussey et al., 2024). Because incarcerated people have little control over their sleep environment, conventional “sleep hygiene” approaches are poorly suited to these contexts. Importantly, no studies assessed how exposures during incarceration extend into the post-release period, leaving questions about the persistence of sleep-related harms and their contribution to reentry challenges and long-term health outcomes. Emerging work focused on the post-release period points to this as a critical gap ((Elumn et al., 2023). Future research—both quantitative and qualitative—should examine how carceral conditions shape sleep and identify and test interventions at both individual and systemic levels.

Measurement approaches also vary widely by country, facility type, and security level, limiting comparability. Few studies incorporated objective measures such as actigraphy or polysomnography (D’Aurizio et al., 2023; Meijers et al., 2015). For example, Camplain et al. (2022) demonstrated that pre-incarceration housing instability influenced sleep during incarceration, illustrating the importance of collecting data across pre-, peri-, and post-incarceration periods to capture continuity in sleep patterns. Without more standardized measures and longitudinal designs, it is difficult to draw conclusions about causal pathways or the durability of intervention effects.

The U.S. context warrants particular attention. Much of the existing literature originates from international settings with distinct legal, cultural, and institutional systems. U.S. prisons, by contrast, are shaped by the legacies of slavery, racialized punishment, and Jim Crow segregation. Because racial and ethnic minorities are disproportionately incarcerated, understanding how incarceration contributes to racial disparities in sleep, and, by extension, disparities in cardiovascular and metabolic disease, remains a research priority. Positioning sleep as both an outcome of carceral conditions and a mechanism through which incarceration drives broader health inequities can inform more targeted policy and health interventions.

Finally, methodological and ethical considerations merit attention. Person-first language is encouraged across health and social science fields but remains inconsistently used in sleep research on incarceration; adopting it can reduce stigma and aligns with best practices in correctional health epidemiology (Bedell et al., 2018). Dewa et al. (2018) explicitly included people who are incarcerated, sleep researchers, and prison staff from the participating facility in a structured co-design (Modified Delphi) of a prison-specific stepped-care pathway for insomnia and subsequent feasibility work showed promise (Dewa et al., 2024). Without the engagement of incarcerated and formerly incarcerated individuals, alongside carceral staff and sleep experts, interventions risk being ill-suited to carceral realities and perpetuating inequities. Addressing these methodological and ethical limitations is essential to building an evidence base that informs structural reforms, clinical interventions, and policies to protect the sleep and health of justice involved populations.

## Data Availability

All data produced in the present work are contained in the manuscript.

## Acknowledgements

We thank everyone who contributed to this scoping review, including Dylan Balter, Kathryn Thomas, Amy Du, and Gul Jana Saeed.

Funding: This work was supported by the National Institutes of Health (NHLBI grant #5K01HL164760-02) and the American Heart Association (grant# 24HERNCDRAP1341360).

## Preferred Reporting Items for Systematic reviews and Meta-Analyses extension for Scoping Reviews (PRISMA-ScR) Checklist

| SECTION | ITEM | PRISMA-ScR CHECKLIST ITEM | REPORTED ON PAGE # |
| --- | --- | --- | --- |
| **TITLE** |  |  |  |
| Title | 1 | Identify the report as a scoping review. | 1-2 |
| Structured summary | 2 | Provide a structured summary that includes (as applicable): background, objectives, eligibility criteria, sources of evidence, charting methods, results, and conclusions that relate to the review questions and objectives. | 3 |
| Rationale | 3 | Describe the rationale for the review in the context of what is already known. Explain why the review questions/objectives lend themselves to a scoping review approach. | 4-5 |
| Objectives | 4 | Provide an explicit statement of the questions and objectives being addressed with reference to their key elements (e.g., population or participants, concepts, and context) or other relevant key elements used to conceptualize the review questions and/or objectives. | 5 |
| Protocol and registration | 5 | Indicate whether a review protocol exists; state if and where it can be accessed (e.g., a Web address); and if available, provide registration information, including the registration number. | 5-6 |
| Eligibility criteria | 6 | Specify characteristics of the sources of evidence used as eligibility criteria (e.g., years considered, language, and publication status), and provide a rationale. | 6-7 |
| Information sources* | 7 | Describe all information sources in the search (e.g., databases with dates of coverage and contact with authors to identify additional sources), as well as the date the most recent search was executed. | 6-7 |
| Search | 8 | Present the full electronic search strategy for at least 1 database, including any limits used, such that it could be repeated. | 7 |
| Selection of sources of evidence† | 9 | State the process for selecting sources of evidence (i.e., screening and eligibility) included in the scoping review. | 7 |
| Data charting process‡ | 10 | Describe the methods of charting data from the included sources of evidence (e.g., calibrated forms or forms that have been tested by the team before their use, and whether data charting was done independently or in duplicate) and any processes for obtaining and confirming data from investigators. | 7 |
| Data items | 11 | List and define all variables for which data were sought and any assumptions and simplifications made. | 7 |
| Critical appraisal of individual sources of evidence§ | 12 | If done, provide a rationale for conducting a critical appraisal of included sources of evidence; describe the methods used and how this information was used in any data synthesis (if appropriate). |  |
| Synthesis of results | 13 | Describe the methods of handling and summarizing the data that were charted. | 7-8 |
| **RESULTS** |  |  |  |
| Selection of sources of evidence | 14 | Give numbers of sources of evidence screened, assessed for eligibility, and included in the review, with reasons for exclusions at each stage, ideally using a flow diagram. | 7-8 |
| Characteristics of sources of evidence | 15 | For each source of evidence, present characteristics for which data were charted and provide the citations. | Supplementary Figure 3 (pg. 65 – 83) |
| Critical appraisal within sources of evidence | 16 | If done, present data on critical appraisal of included sources of evidence (see item 12). |  |
| Results of individual sources of evidence | 17 | For each included source of evidence, present the relevant data that were charted that relate to the review questions and objectives. | 8-15 |
| Synthesis of results | 18 | Summarize and/or present the charting results as they relate to the review questions and objectives. | Supplementary Figure 3 (pg. 65 – 83) |
| Summary of evidence | 19 | Summarize the main results (including an overview of concepts, themes, and types of evidence available), link to the review questions and objectives, and consider the relevance to key groups. | 15-16 |
| Limitations | 20 | Discuss the limitations of the scoping review process. | 16 |
| Conclusions | 21 | Provide a general interpretation of the results with respect to the review questions and objectives, as well as potential implications and/or next steps. | 16-17 |
| Funding | 22 | Describe sources of funding for the included sources of evidence, as well as sources of funding for the scoping review. Describe the role of the funders of the scoping review. | 2 |

JBIC = Joanna Briggs Institute; PRISMA-ScR = Preferred Reporting Items for Systematic reviews and Meta-Analyses extension for Scoping Reviews.
From: Tricco AC, Lillie E, Zarin W, O'Brien KK, Colquhoun H, Levac D, et al. PRISMA Extension for Scoping Reviews (PRISMA-ScR): Checklist and Explanation. *Ann Intern Med*. 2018;169:467–473. [doi: 10.7326/M18-0850](https://doi.org/10.7326/M18-0850)

## Supplementary Digital Appendix 2: Search Strategies

### Ovid Embase

1. Prison/
2. Prisoner/
3. Offender/
4. (incarcerat* or decarcerat* or prison* or jail* or criminal* or correctional or corrections officer* or inmate* or convict* or offender* or detained or detainee or imprison* or behind bars or penitentiar* or detention*).mp.
5. (justice-involve* or justice involve* or ex-con or ex-cons or felon* or parol* or probat*).mp.
6. or/1-5
7. exp Sleep Disorders/
8. (sleep* or narcoleps* or somnolen* or drowsiness or exploding head syndrome* or hypersomni* or hypnagogic hallucination* or hypnic headache* or insomni* or nightmare* or parasomnia* or paroniria* or periodic limb movement disorder* or sopor or agrypnia* or hyposomnia* or nocturnal apnea* or nocturnal apnoea* or dyssomnia* or night terror* or pavor nocturnus).mp.
9. ((abnormal or excessive or unpleasant or vivid or fragmented) adj3 dream*).mp.
10. or/7-9
11. 6 and 10

### Ovid MEDLINE(R) ALL

1. Prisons/
2. Prisoners/
3. Criminals/
4. (incarcerat* or decarcerat* or prison* or jail* or criminal* or correctional or corrections officer* or inmate* or convict* or offender* or detained or detainee or imprison* or behind bars or penitentiar* or detention*).mp.
5. (justice-involve* or justice involve* or ex-con or ex-cons or felon* or parol* or probat*).mp.
6. or/1-5
7. exp Sleep Wake Disorders/
8. (sleep* or narcoleps* or somnolen* or drowsiness or exploding head syndrome* or hypersomni* or hypnagogic hallucination* or hypnic headache* or insomni* or nightmare* or parasomnia* or paroniria* or periodic limb movement disorder* or sopor or agrypnia* or hyposomnia* or nocturnal apnea* or nocturnal apnoea* or dyssomnia* or night terror* or pavor nocturnus).mp.
9. ((abnormal or excessive or unpleasant or vivid or fragmented) adj3 dream*).mp.
10. or/7-9
11. 6 and 10

### Ovid APA PsycInfo

1. exp Prisons/
2. exp incarcerated/
3. exp criminal offenders/
4. (incarcerat* or decarcerat* or prison* or jail* or criminal* or correctional or corrections officer* or inmate* or convict* or offender* or detained or detainee or imprison* or behind bars or penitentiar* or detention*).mp.
5. (justice-involve* or justice involve* or ex-con or ex-cons or felon* or parol* or probat*).mp.
6. or/1-5
7. exp Sleep Wake Disorders/
8. (sleep* or narcoleps* or somnolen* or drowsiness or exploding head syndrome* or hypersomni* or hypnagogic hallucination* or hypnic headache* or insomni* or nightmare* or parasomnia* or paroniria* or periodic limb movement disorder* or sopor or agrypnia* or hyposomnia* or nocturnal apnea* or nocturnal apnoea* or dyssomnia* or night terror* or pavor nocturnus).mp.
9. ((abnormal or excessive or unpleasant or vivid or fragmented) adj3 dream*).mp.
10. 7 or 8 or 9
11. 6 and 10

### Ovid APA PsycExtra

1. exp Prisons/
2. exp incarcerated/
3. exp criminal offenders/
4. (incarcerat* or decarcerat* or prison* or jail* or criminal* or correctional or corrections officer* or inmate* or convict* or offender* or detained or detainee or imprison* or behind bars or penitentiar* or detention*).mp.
5. (justice-involve* or justice involve* or ex-con or ex-cons or felon* or parol* or probat*).mp.
6. 6 or/1-5
7. exp Sleep Wake Disorders/
8. (sleep* or narcoleps* or somnolen* or drowsiness or exploding head syndrome* or hypersomni* or hypnagogic hallucination* or hypnic headache* or insomni* or nightmare* or parasomnia* or paroniria* or periodic limb movement disorder* or sopor or agrypnia* or hyposomnia* or nocturnal apnea* or nocturnal apnoea* or dyssomnia* or night terror* or pavor nocturnus).mp.
9. ((abnormal or excessive or unpleasant or vivid or fragmented) adj3 dream*).mp.
10. 7 or 8 or 9
11. 6 and 10

### PubMed

(sleep*[Title/Abstract] OR narcoleps*[Title/Abstract] OR somnolen*[Title/Abstract] OR drowsiness[Title/Abstract] OR exploding head syndrome*[Title/Abstract] OR hypersomni*[Title/Abstract] OR hypnagogic hallucination*[Title/Abstract] OR hypnic headache*[Title/Abstract] OR insomni*[Title/Abstract] OR nightmare*[Title/Abstract] OR parasomnia*[Title/Abstract] OR paroniria*[Title/Abstract] OR periodic limb movement disorder*[Title/Abstract] OR sopor[Title/Abstract] OR agrypnia*[Title/Abstract] OR hyposomnia*[Title/Abstract] OR nocturnal apnea*[Title/Abstract] OR nocturnal apnoea*[Title/Abstract] OR dyssomnia*[Title/Abstract] OR night terror*[Title/Abstract] OR pavor nocturnus[Title/Abstract] OR abnormal dream*[Title/Abstract] OR excessive dream*[Title/Abstract] OR unpleasant dream*[Title/Abstract] OR vivid dream*[Title/Abstract] OR fragmented dream*[Title/Abstract]) AND (incarcerat*[Title/Abstract] OR decarcerat*[Title/Abstract] OR prison*[Title/Abstract] OR jail*[Title/Abstract] OR criminal*[Title/Abstract] OR correctional[Title/Abstract] OR corrections officer*[Title/Abstract] OR inmate*[Title/Abstract] OR convict*[Title/Abstract] OR offender*[Title/Abstract] OR detained[Title/Abstract] OR detainee[Title/Abstract] OR imprison*[Title/Abstract] OR behind bars[Title/Abstract] OR penitentiar*[Title/Abstract] OR detention*[Title/Abstract] OR justice- involve*[Title/Abstract] OR justice involve*[Title/Abstract] OR ex-con[Title/Abstract] OR ex- cons[Title/Abstract] OR felon*[Title/Abstract] OR parol*[Title/Abstract] OR probat[Title/Abstract])

### Criminal Justice Abstracts

S1 (TI food-secur* OR food-insecur* OR “food basket*” OR “food bank*” OR foodbank* OR “food pantr*” OR “food suppl*” OR “food insufficien*” OR AB food-secur* OR food-insecur* OR “food basket*” OR “food bank*” OR foodbank* OR “food pantr*” OR “food suppl*” OR “food insufficien*” OR TI (food secur* or food insecur* or nutrition secur* or nutrition insecur* or urban garden* or urban farmer* or community garden* or community farmer* or community agriculture or home garden*) OR AB (food secur* or food insecur* or nutrition secur* or nutrition insecur* or urban garden* or urban farmer* or community garden* or community farmer* or community agriculture or home garden*))

S2 TX ((Caribbean or Carib or “West Indies” or Anguilla* or Antilles or Antigua* or Aruba* or Barbuda or Bahamas or Bahamian* or Barbados or Barbadian* or Barthelemy or “Saint Barthelemois” or Barts or Bermuda or Bermudian* or Bonaire or Bonairian* or Cuba* or Cayman or Caymanian* or Curacao* or Caicos or Belonger* or croix or Crucian* or Dominica or “Dominican Republic” or Dominican* or “Santo Domingo” or Eustatius or Grenada or Grenadian* or Guadeloupe* or grenadines or Haiti* or Hispaniola or Jamaica* or Martinique or Martiniquais or Martinican* or “Puerto Rico” or “Puerto Rican*” or Nevis or Nevisian* or

Montserrat* or “Virgin Island” or “Virgin Islands” or “Virgin Islander S1 TI (incarcerat* or decarcerat* or prison* or jail* or criminal* or correctional or corrections officer* or inmate* or convict* or offender* or detained or detainee or imprison* or behind bars or penitentiar* or detention* or justice-involve* or justice involve* or ex-con or ex-cons or felon* or parol* or probat*) OR AB (incarcerat* or decarcerat* or prison* or jail* or criminal* or correctional or corrections officer* or inmate* or convict* or offender* or detained or detainee or imprison* or behind bars or penitentiar* or detention* or justice-involve* or justice involve* or ex-con or ex- cons or felon* or parol* or probat*)

S2 TI (sleep* or narcoleps* or somnolen* or drowsiness or exploding head syndrome* or hypersomni* or hypnagogic hallucination* or hypnic headache* or insomni* or nightmare* or parasomnia* or paroniria* or periodic limb movement disorder* or sopor or agrypnia* or hyposomnia* or nocturnal apnea* or nocturnal apnoea* or dyssomnia* or night terror* or pavor nocturnus or abnormal dream* or excessive dream* or unpleasant dream* or vivid dream* or fragmented dream*) OR AB (sleep* or narcoleps* or somnolen* or drowsiness or exploding head syndrome* or hypersomni* or hypnagogic hallucination* or hypnic headache* or insomni* or nightmare* or parasomnia* or paroniria* or periodic limb movement disorder* or sopor or agrypnia* or hyposomnia* or nocturnal apnea* or nocturnal apnoea* or dyssomnia* or night terror* or pavor nocturnus or abnormal dream* or excessive dream* or unpleasant dream* or vivid dream* or fragmented dream*)

S3 S1 and S2

### National Criminal Justice Reference Service Abstracts Database

abstract((sleep* or narcoleps* or somnolen* or drowsiness or exploding head syndrome* or hypersomni* or hypnagogic hallucination* or hypnic headache* or insomni* or nightmare* or parasomnia* or paroniria* or periodic limb movement disorder* or sopor or agrypnia* or hyposomnia* or nocturnal apnea* or nocturnal apnoea* or dyssomnia* or night terror* or pavor nocturnus or abnormal dream* or excessive dream* or unpleasant dream* or vivid dream* or fragmented dream*)) AND abstract((incarcerat* or decarcerat* or prison* or jail* or criminal* or correctional or corrections officer* or inmate* or convict* or offender* or detained or detainee or imprison* or behind bars or penitentiar* or detention* or justice-involve* or justice involve* or ex-con or ex-cons or felon* or parol* or probat*))

### Cochrane Library

#1 (incarcerat* or decarcerat* or prison* or jail* or criminal* or correctional or corrections officer* or inmate* or convict* or offender* or detained or detainee or imprison* or behind bars or penitentiar* or detention*):ti,ab or (justice-involve* or justice involve* or ex-con or ex-cons or felon* or parol* or probat*):ti,ab

#2 (sleep* or narcoleps* or somnolen* or drowsiness or exploding head syndrome* or hypersomni* or hypnagogic hallucination* or hypnic headache* or insomni* or nightmare* or parasomnia* or paroniria* or periodic limb movement disorder* or sopor or agrypnia* or hyposomnia* or nocturnal apnea* or nocturnal apnoea* or dyssomnia* or night terror* or pavor nocturnus):ti,ab or ((abnormal or excessive or unpleasant or vivid or fragmented) near/3 dream*):ti,ab

#3 #1 and #2

The Cochrane Library database includes:

1. Cochrane Database of Systematic Reviews
2. Cochrane Central Register of Controlled Trials
3. Cochrane Clinical Answers

### Google Scholar (Via Harzig’s Publish or Perlish)

prison sleep disorders

### Scopus

(TITLE-ABS-KEY (incarcerat* OR decarcerat* OR prison* OR jail* OR criminal* OR correctional OR corrections officer* OR inmate* OR convict* OR offender* OR detained OR detainee OR imprison* OR “behind bars” OR penitentiar* OR detention*) OR TITLE-ABS-KEY (justice-involve* OR “justice involve*” OR ex-con OR ex-cons OR felon* OR parol* OR probat*)) AND (TITLE-ABS-KEY (sleep* OR narcoleps* OR somnolen* OR drowsiness OR “exploding head syndrome*” OR hypersomni* OR “hypnagogic hallucination*” OR “hypnic headache*” OR insomni* OR nightmare* OR parasomnia* OR paroniria* OR “periodic limb movement disorder*” OR sopor OR agrypnia* OR hyposomnia* OR “nocturnal apnea*” OR “nocturnal apnoea*” OR dyssomnia* OR “night terror*” OR “pavor nocturnus”) OR TITLE-ABS-KEY ((abnormal OR excessive OR unpleasant OR vivid OR fragmented) W/3 dream*))

### Web of Science Core Collection

TS=(incarcerat* or decarcerat* or prison* or jail* or criminal* or correctional or corrections officer* or inmate* or convict* or offender* or detained or detainee or imprison* or behind bars or penitentiar* or detention*) or TS=(justice-involve* or justice involve* or ex-con or ex-cons or felon* or parol* or probat*)

#2 TS=(sleep* or narcoleps* or somnolen* or drowsiness or “exploding head syndrome*” or hypersomni* or “hypnagogic hallucination*” or “hypnic headache*” or insomni* or nightmare* or parasomnia* or paroniria* or “periodic limb movement disorder*” or sopor or agrypnia* or hyposomnia* or “nocturnal apnea*” or “nocturnal apnoea*” or dyssomnia* or “night terror*” or “pavor nocturnus”) or TS=((abnormal or excessive or unpleasant or vivid or fragmented) near/3 dream*)

#3 #1 and #2

The Core Collection included in this review is:

1. Science Citation Index Expanded (1900 - Data Searched)
2. Social Sciences Citation Index (1900 - Date Searched)
3. Art & Humanities - (1975 - Date Searched)
4. Conference Proceedings Citation Index - Science (1991 - Date Searched)
5. Conference Proceedings Citation Index - Social Sciences and Humanities (1991 - Date Searched)
6. Book Citation Index - Science (2005 - Date Searched)
7. Book Citation Index - Social Sciences and Humanities (2005 - Date Searched)
8. Emerging Source Citation Index - (2018 - Date Searched)
9. Current Chemical Reactions (1985 - Date Searched)
10. Index Chemicus (1993 - Date Searched)

---

**Supplementary Table 3:**
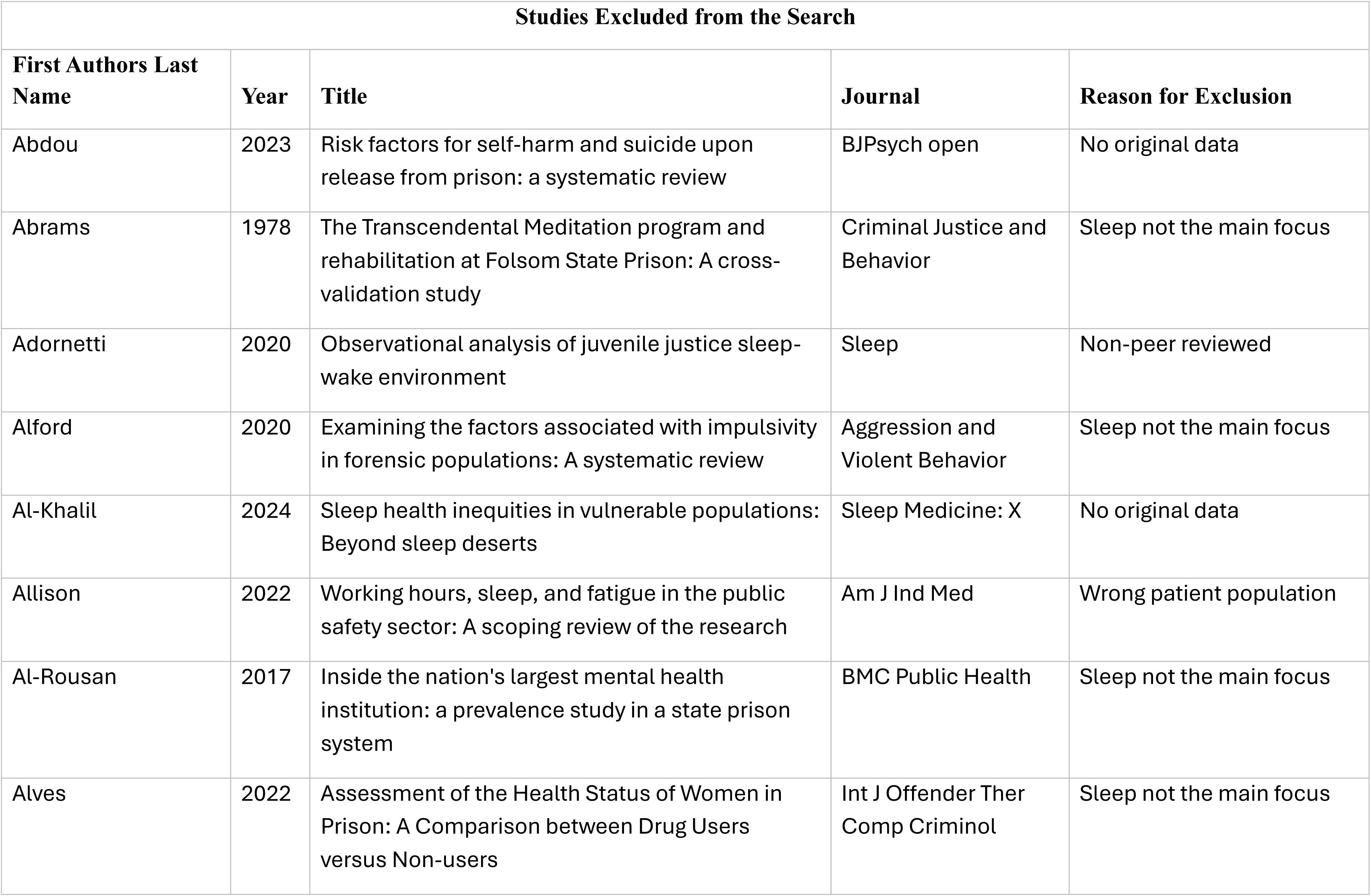
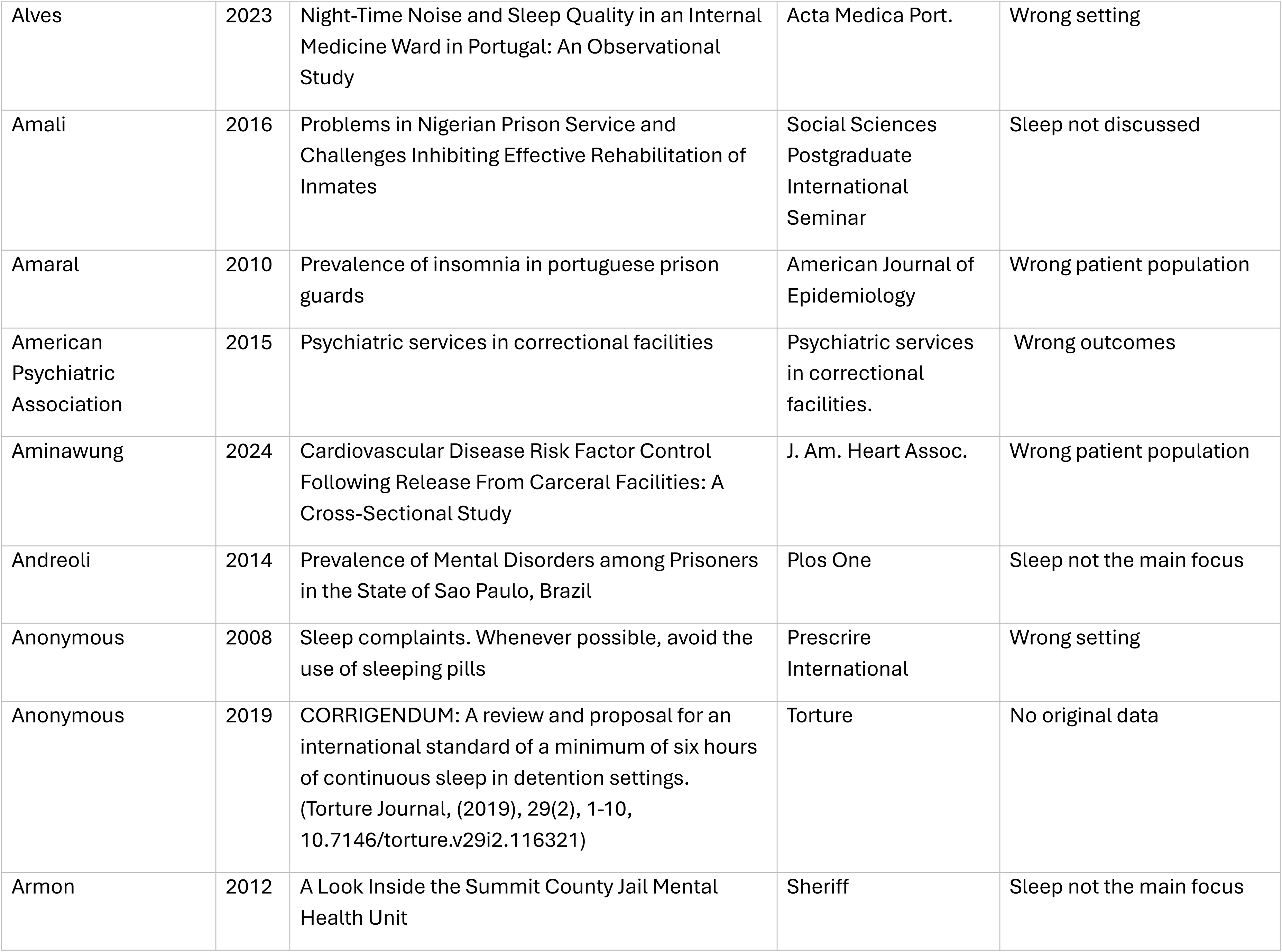
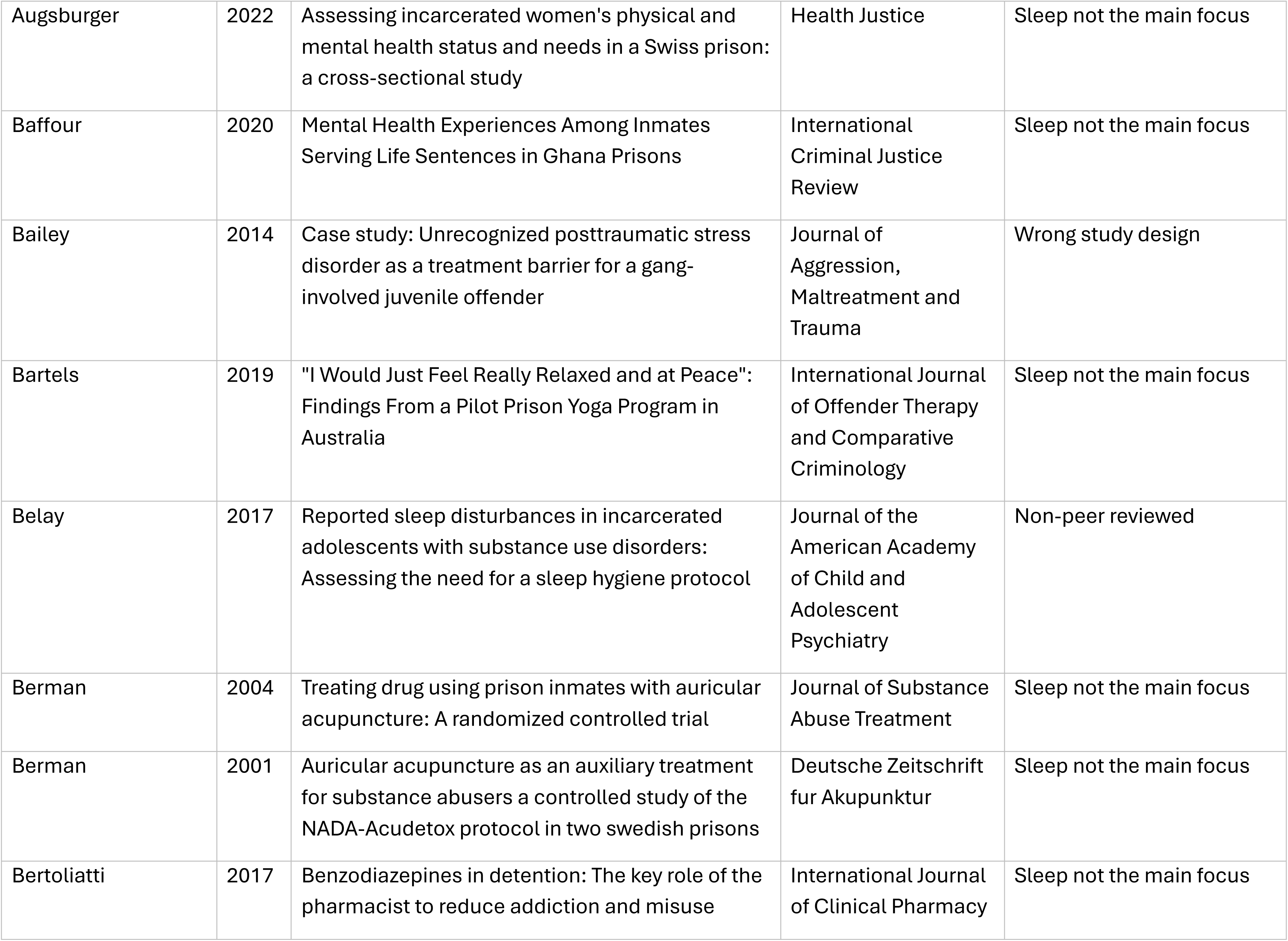
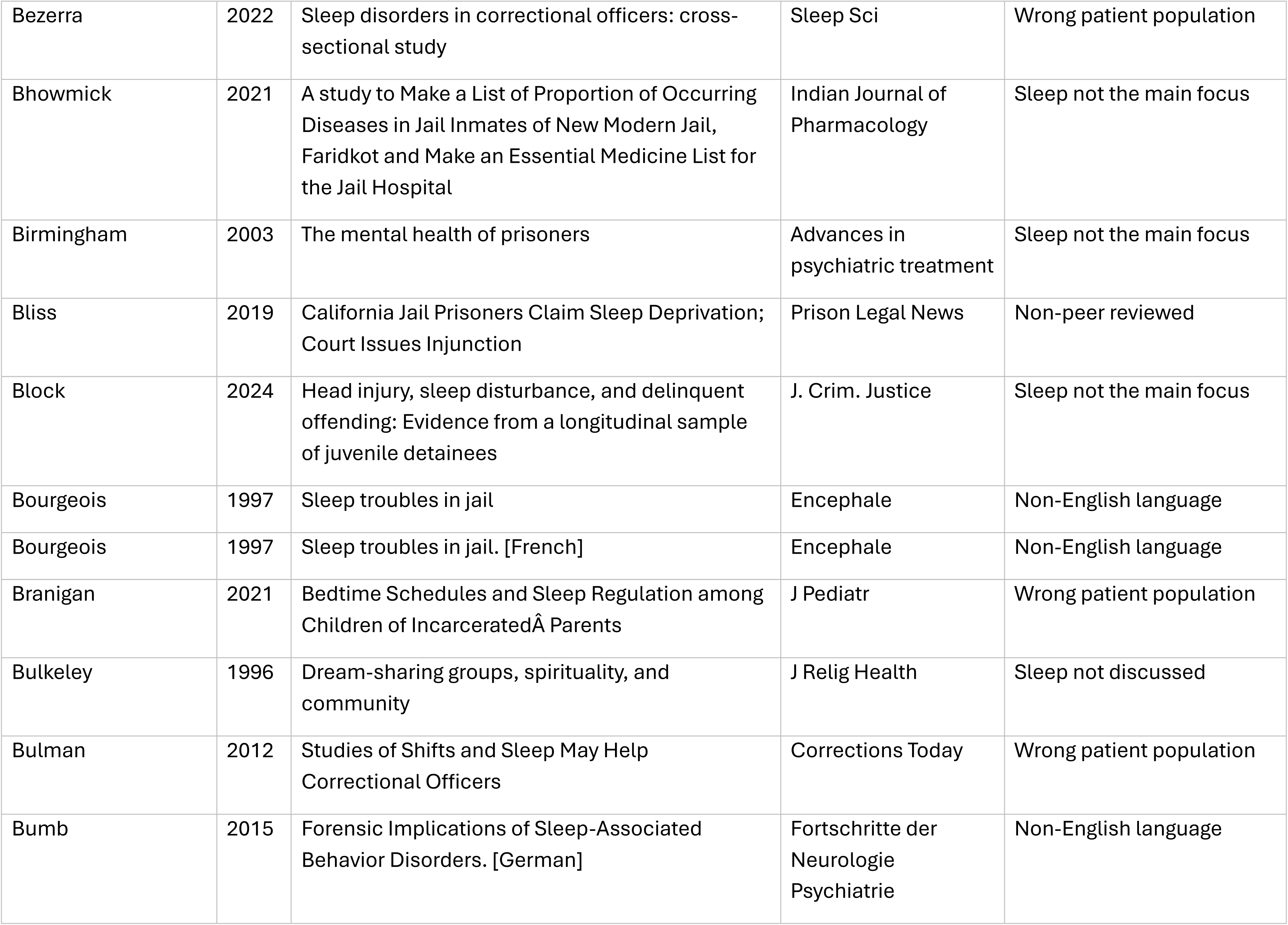
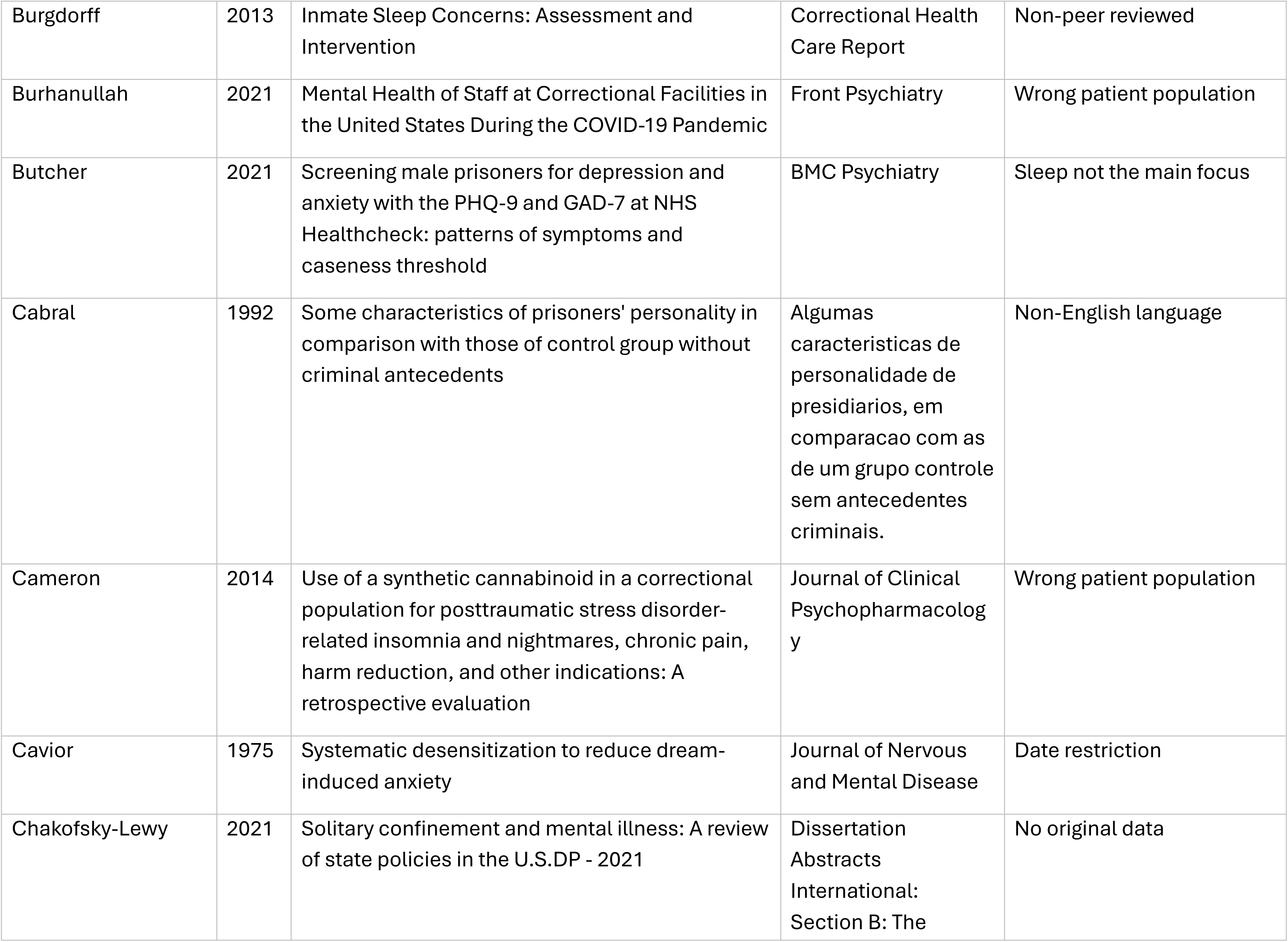
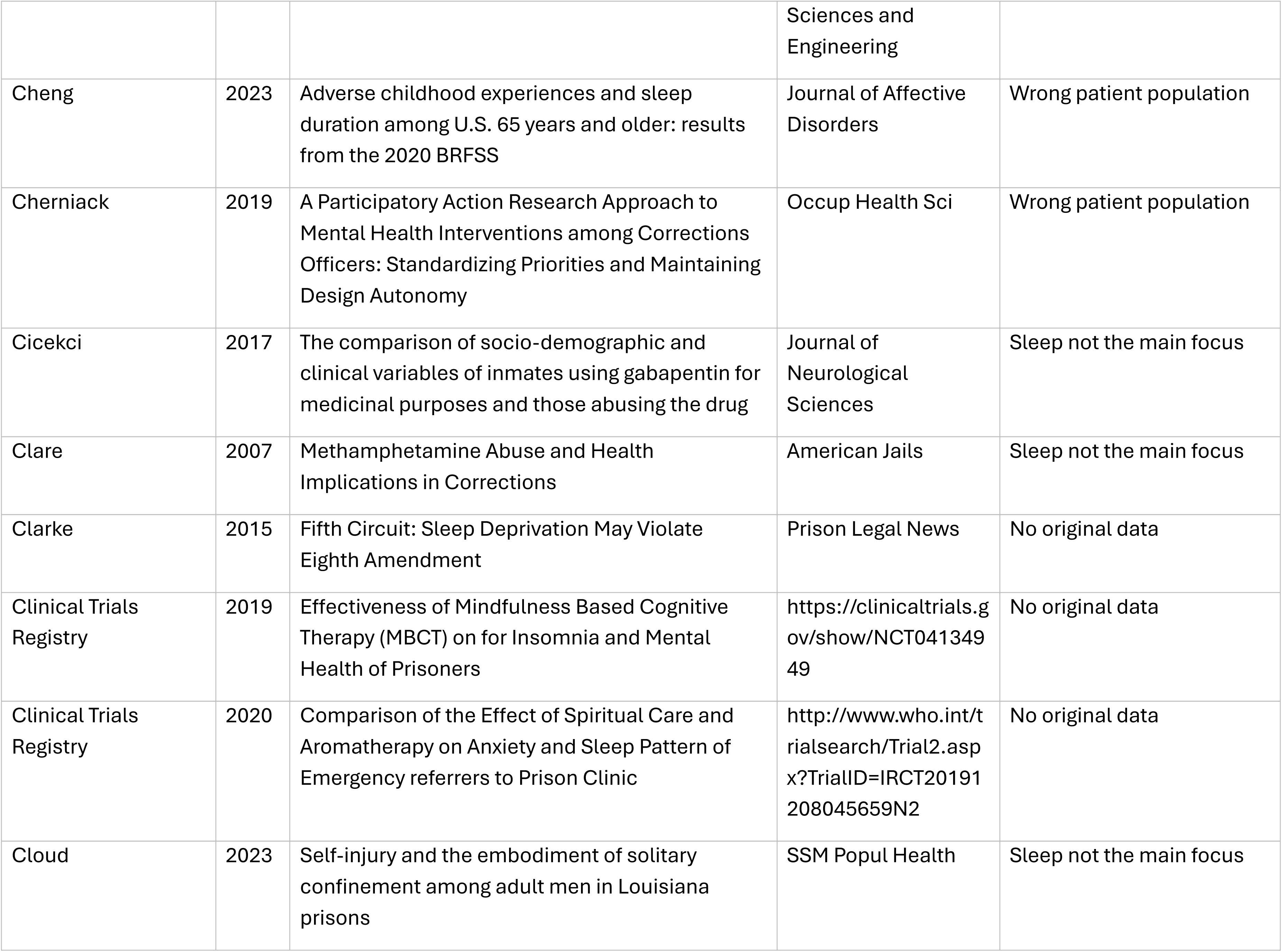
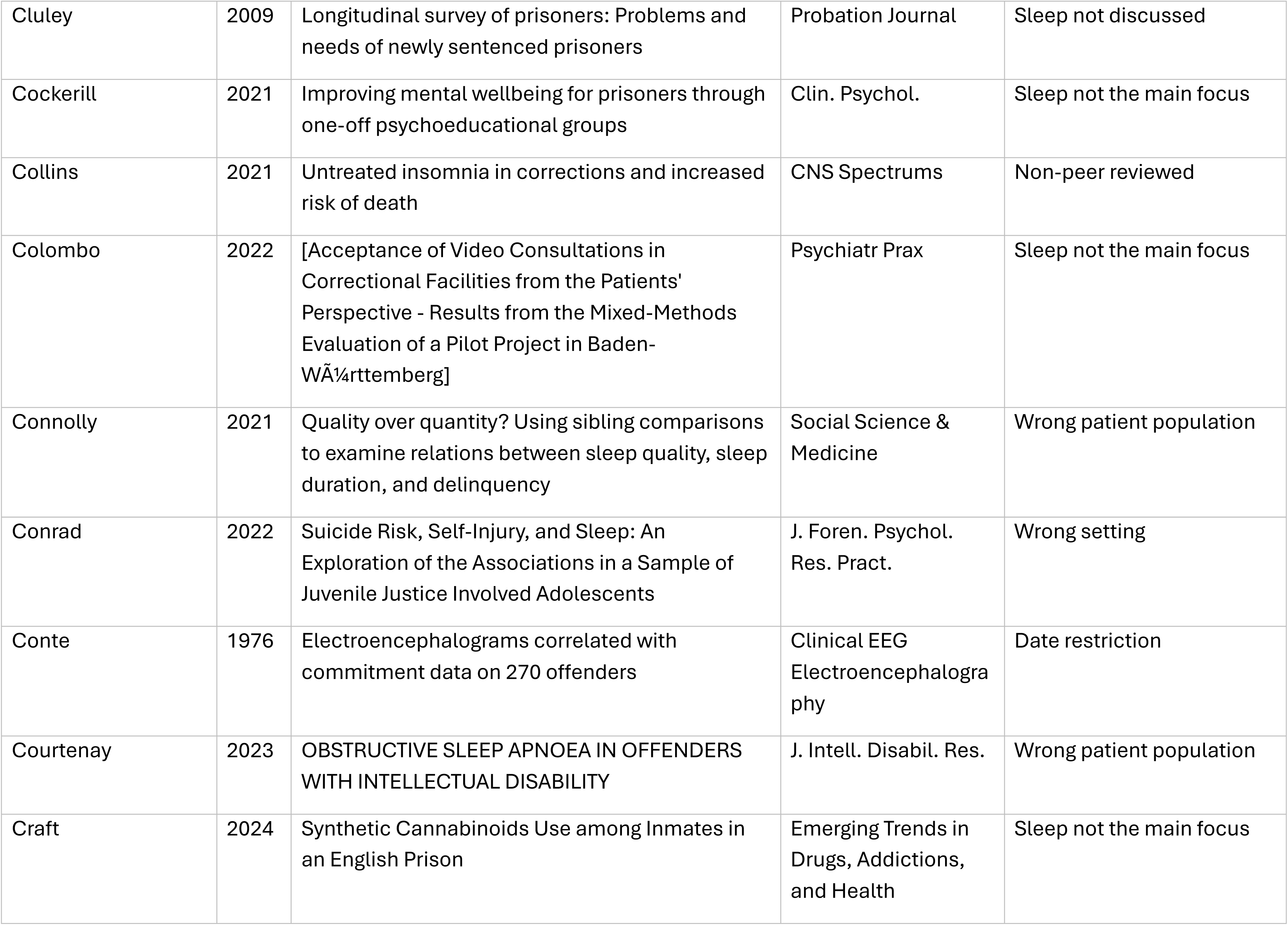
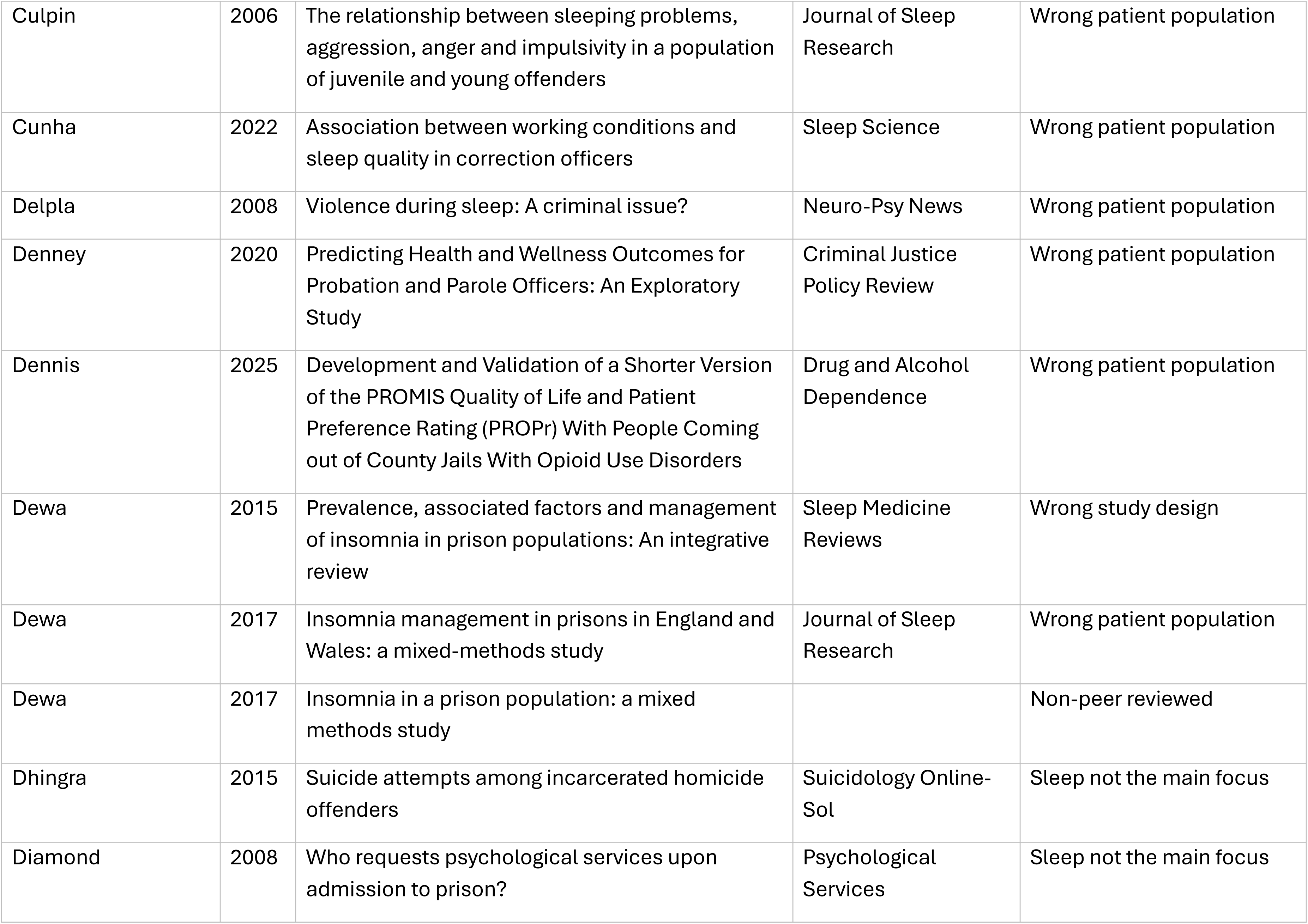
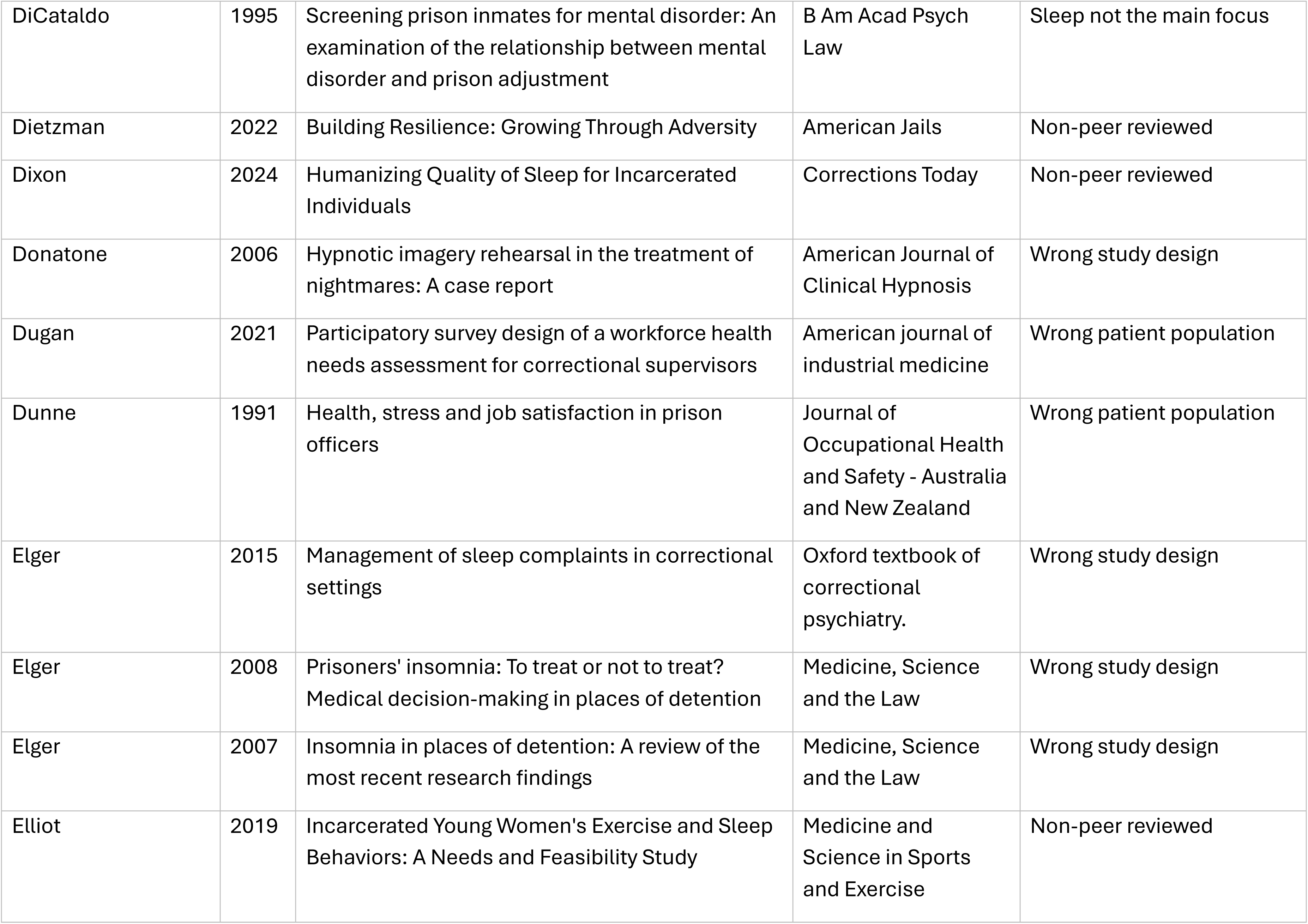
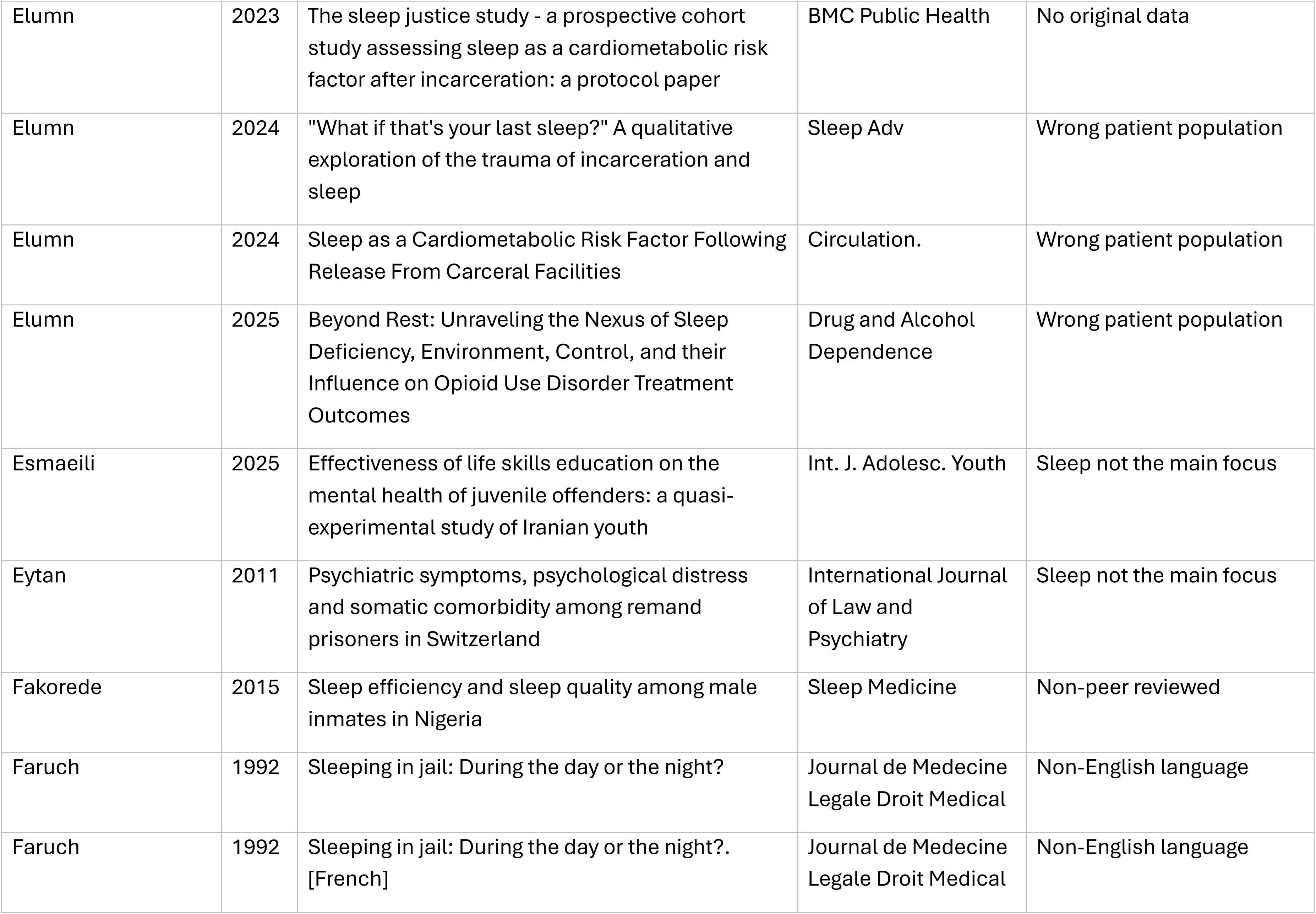
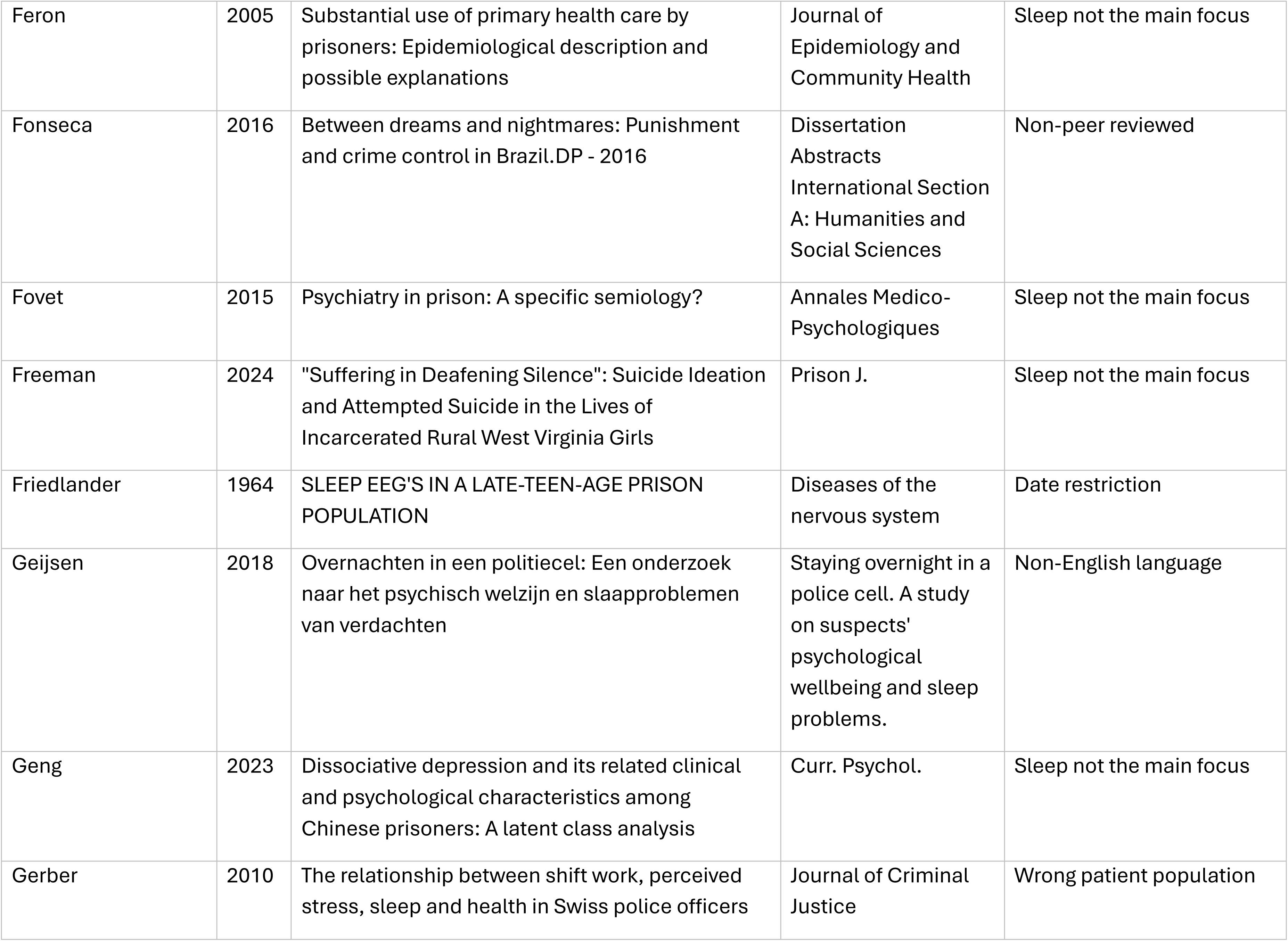
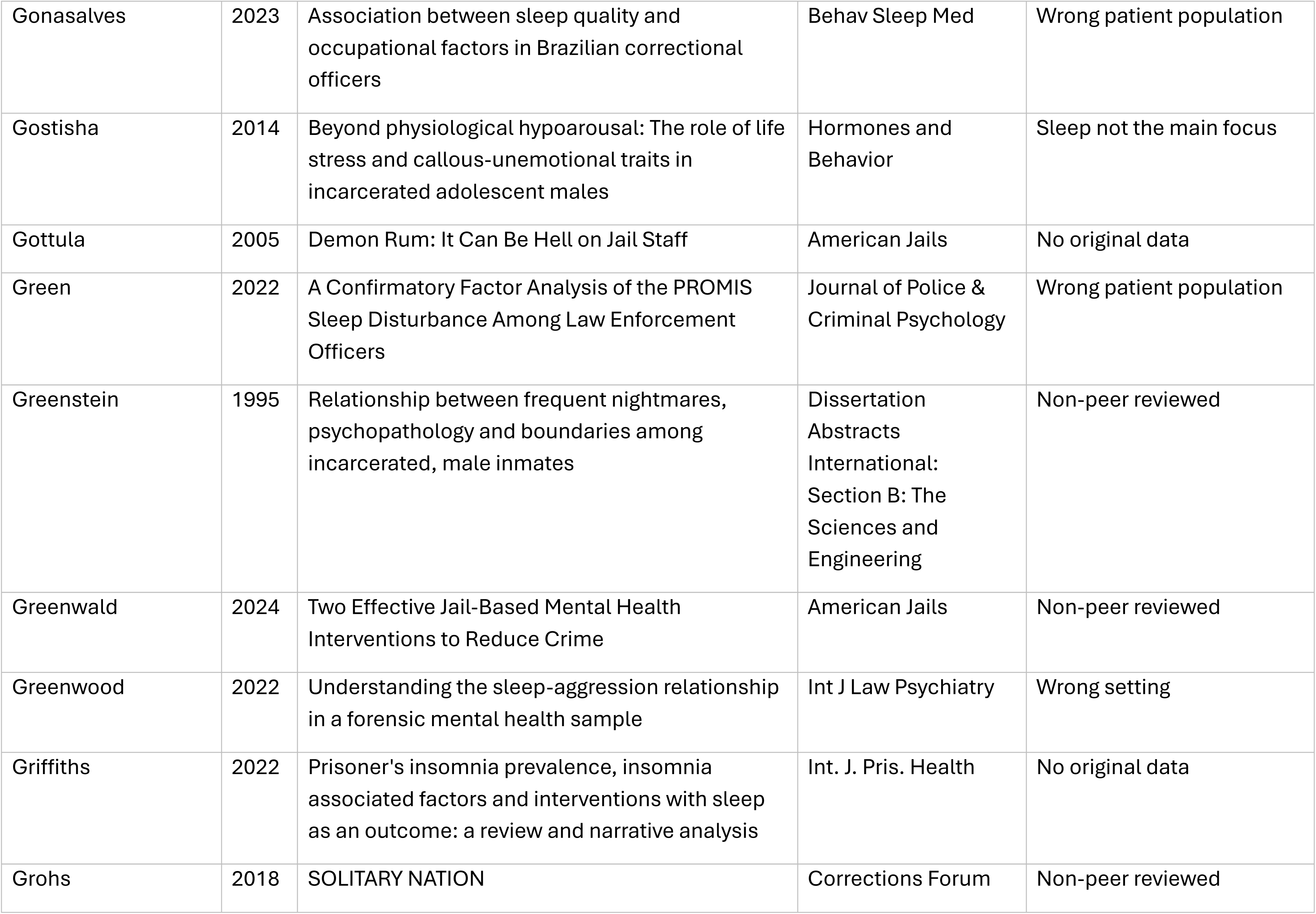
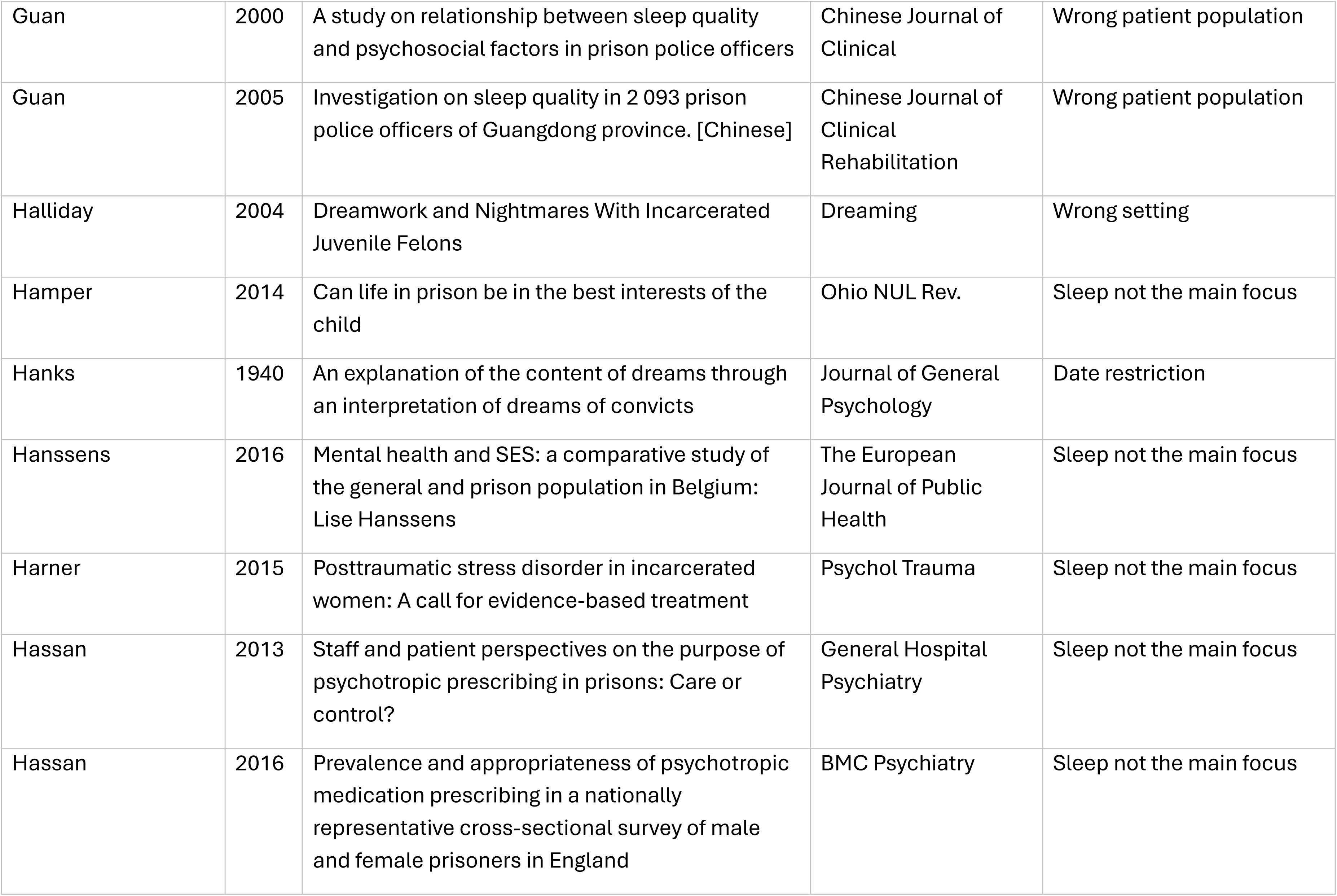
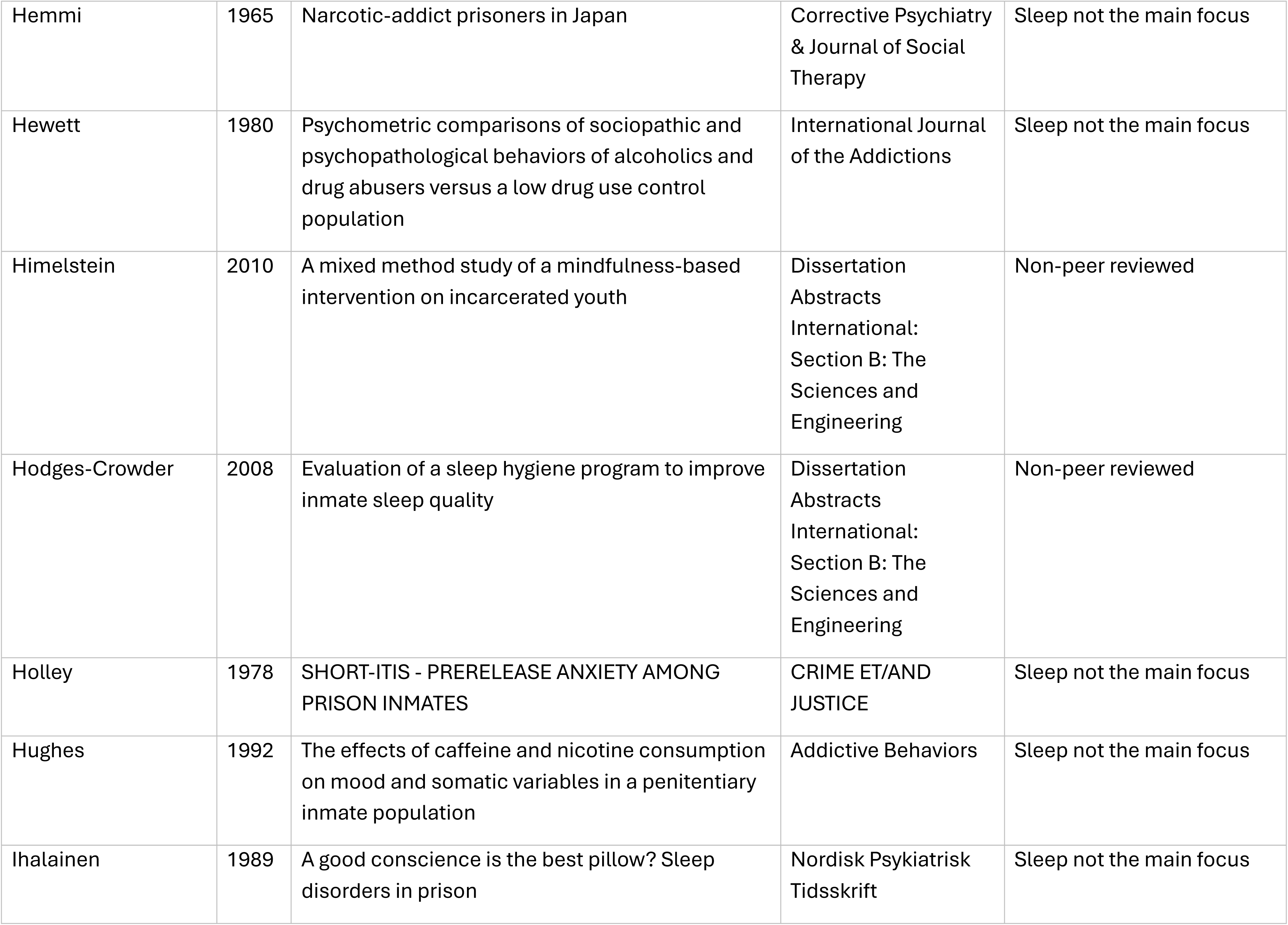
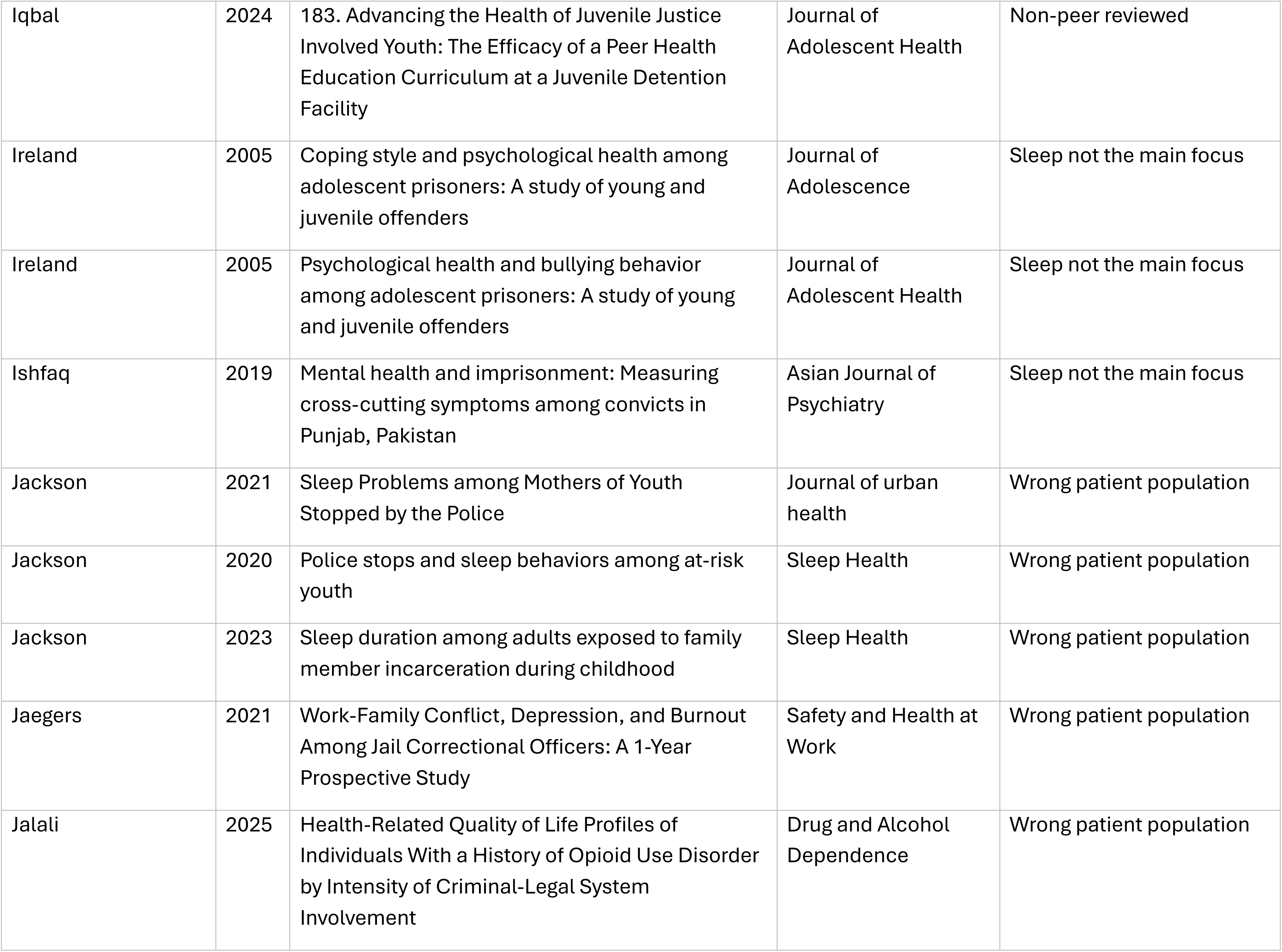
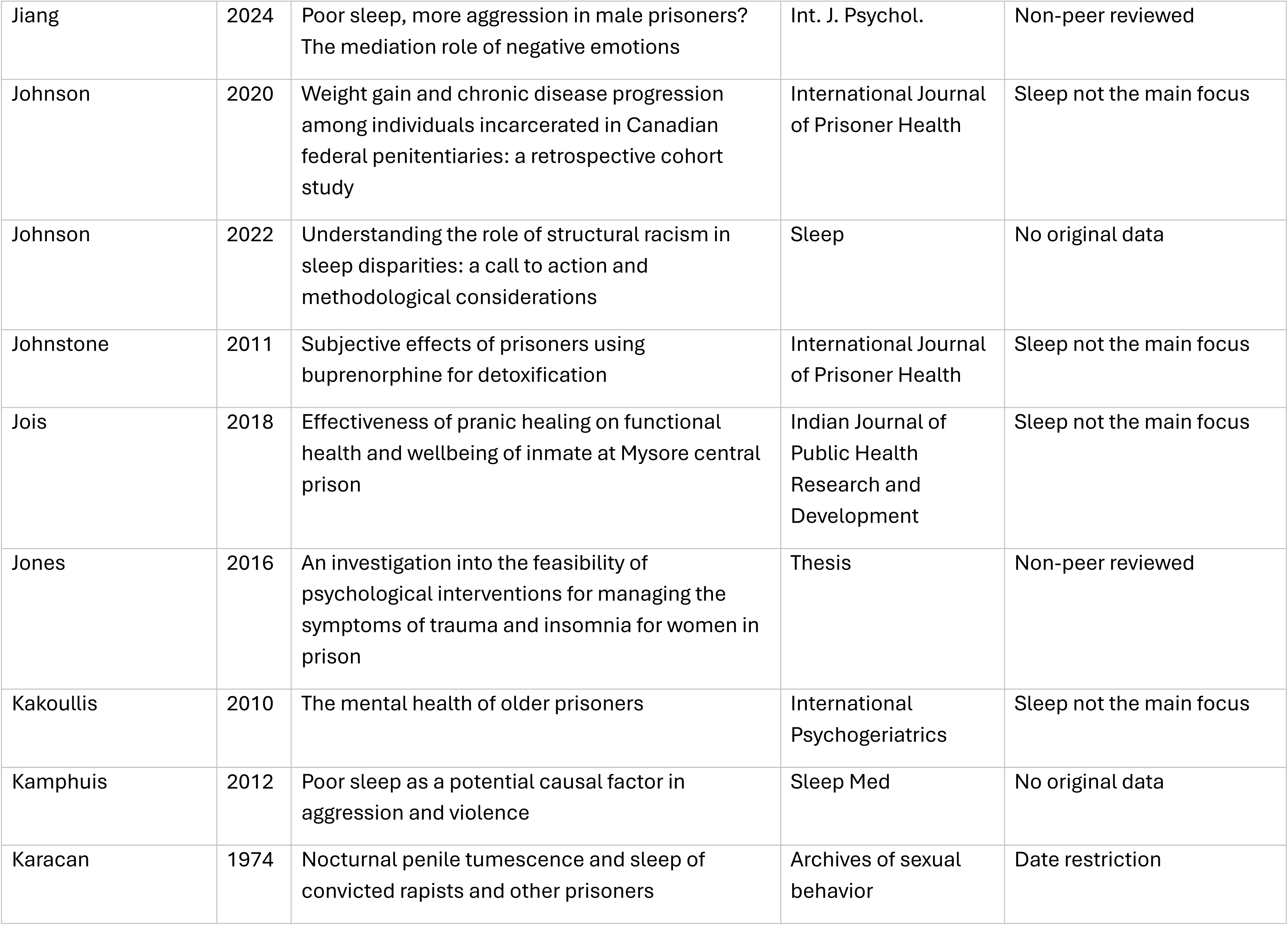
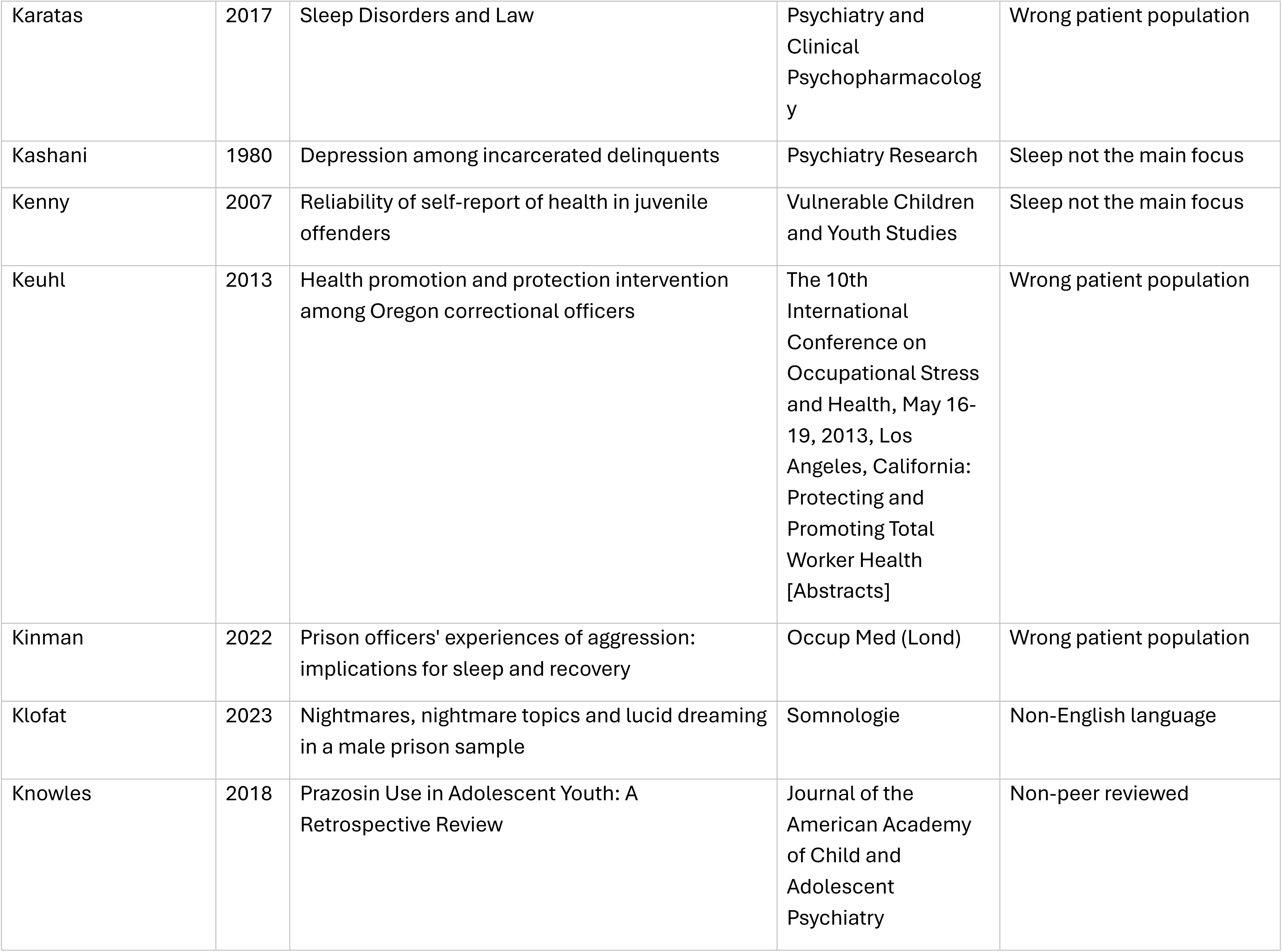
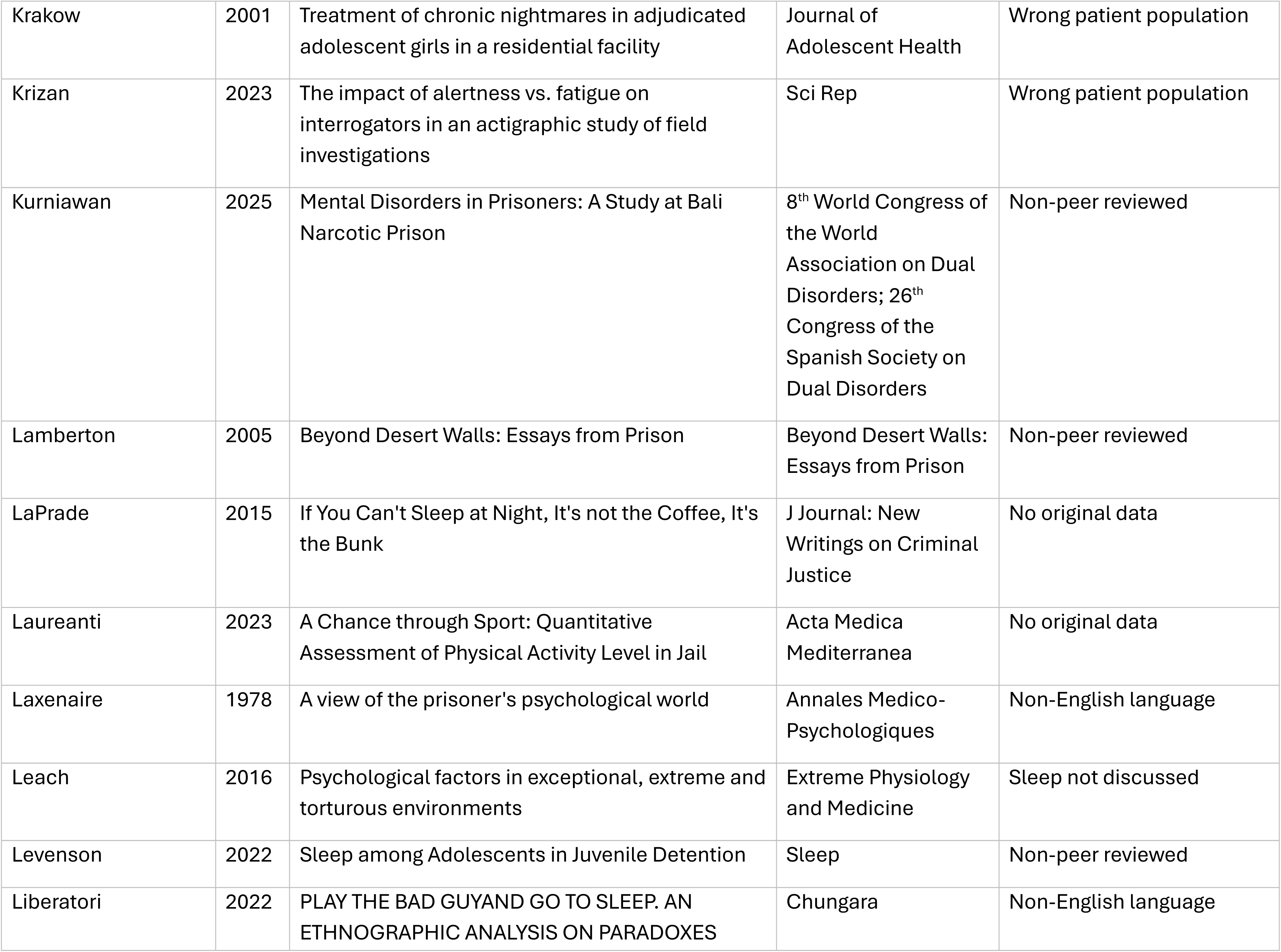
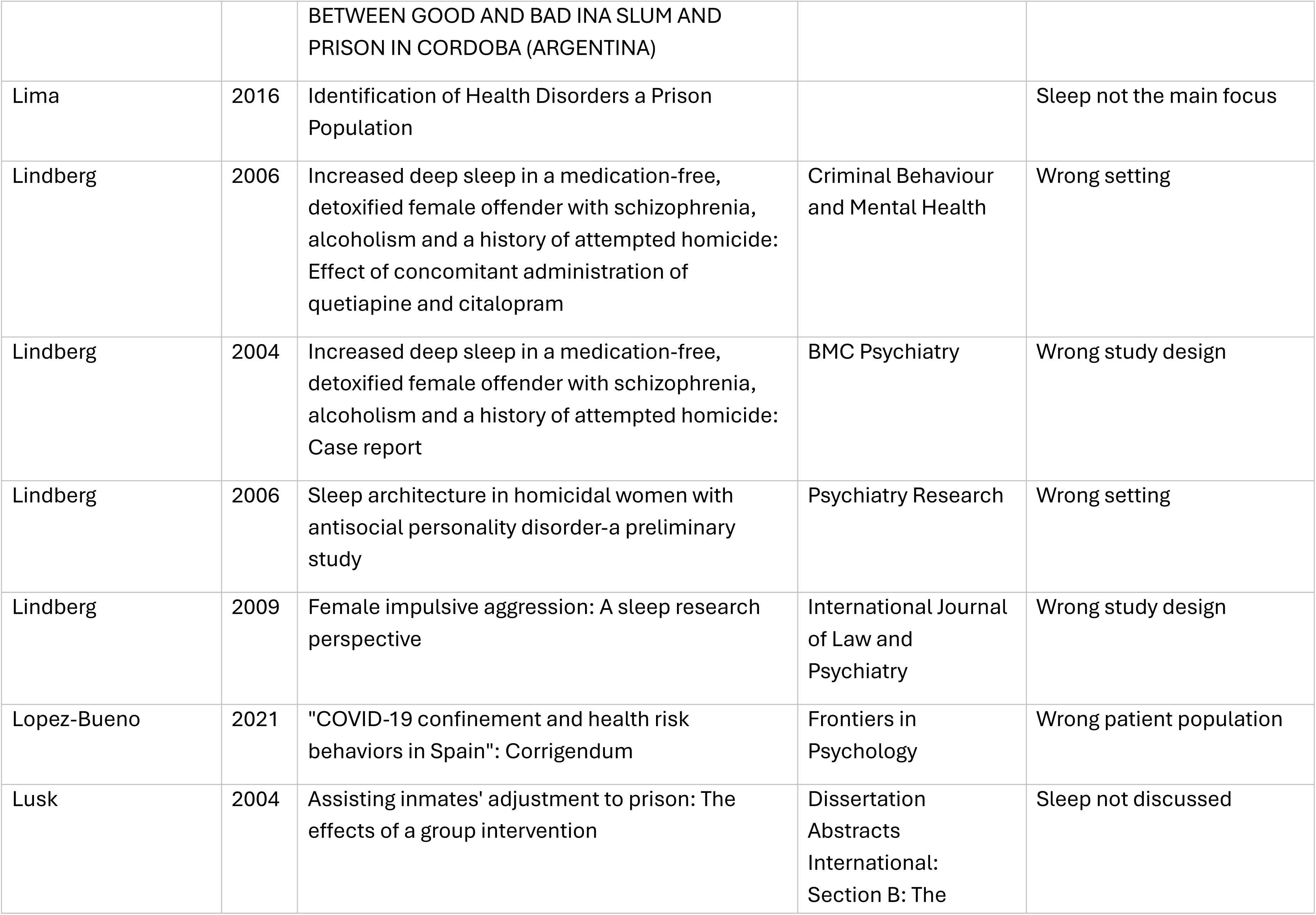
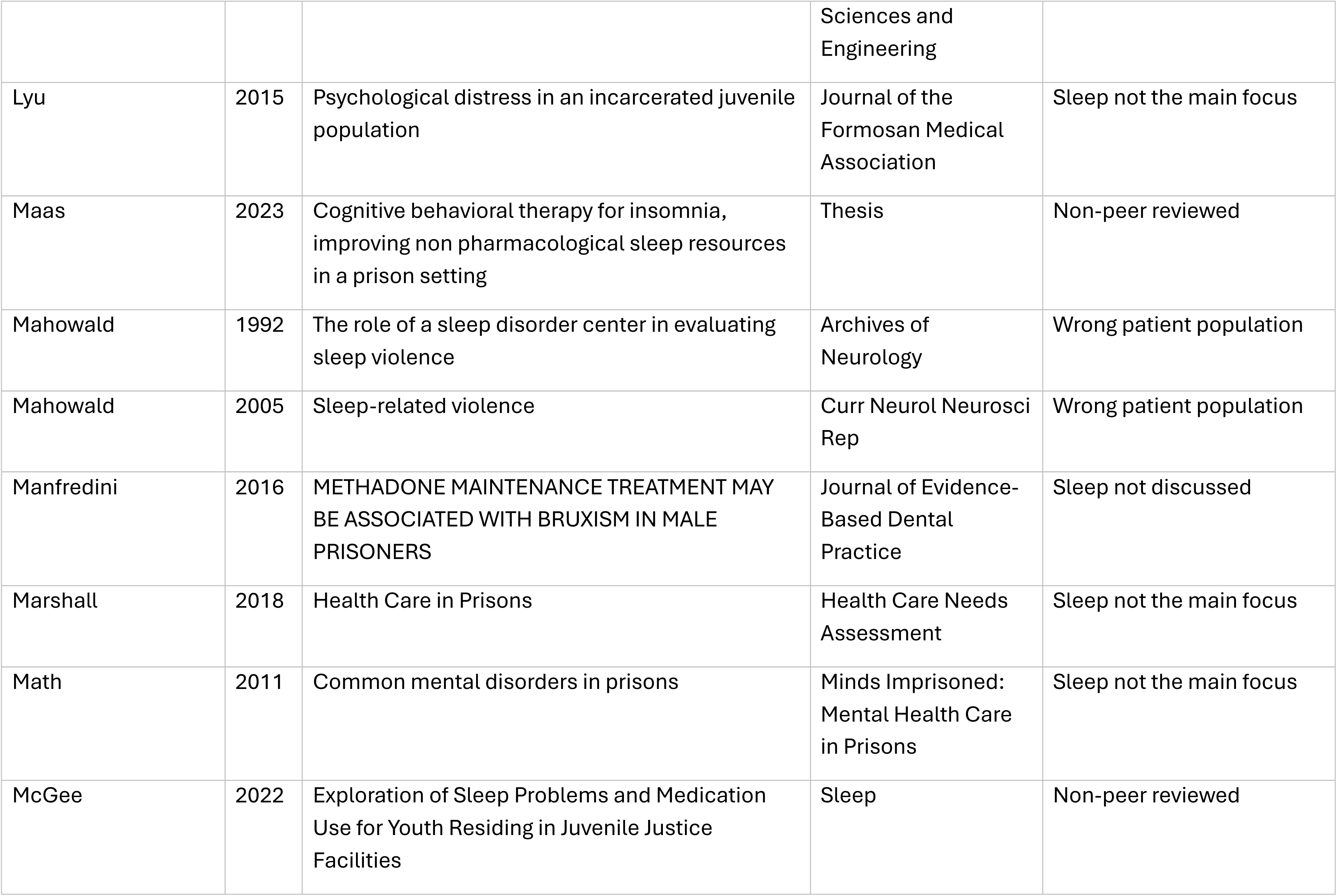
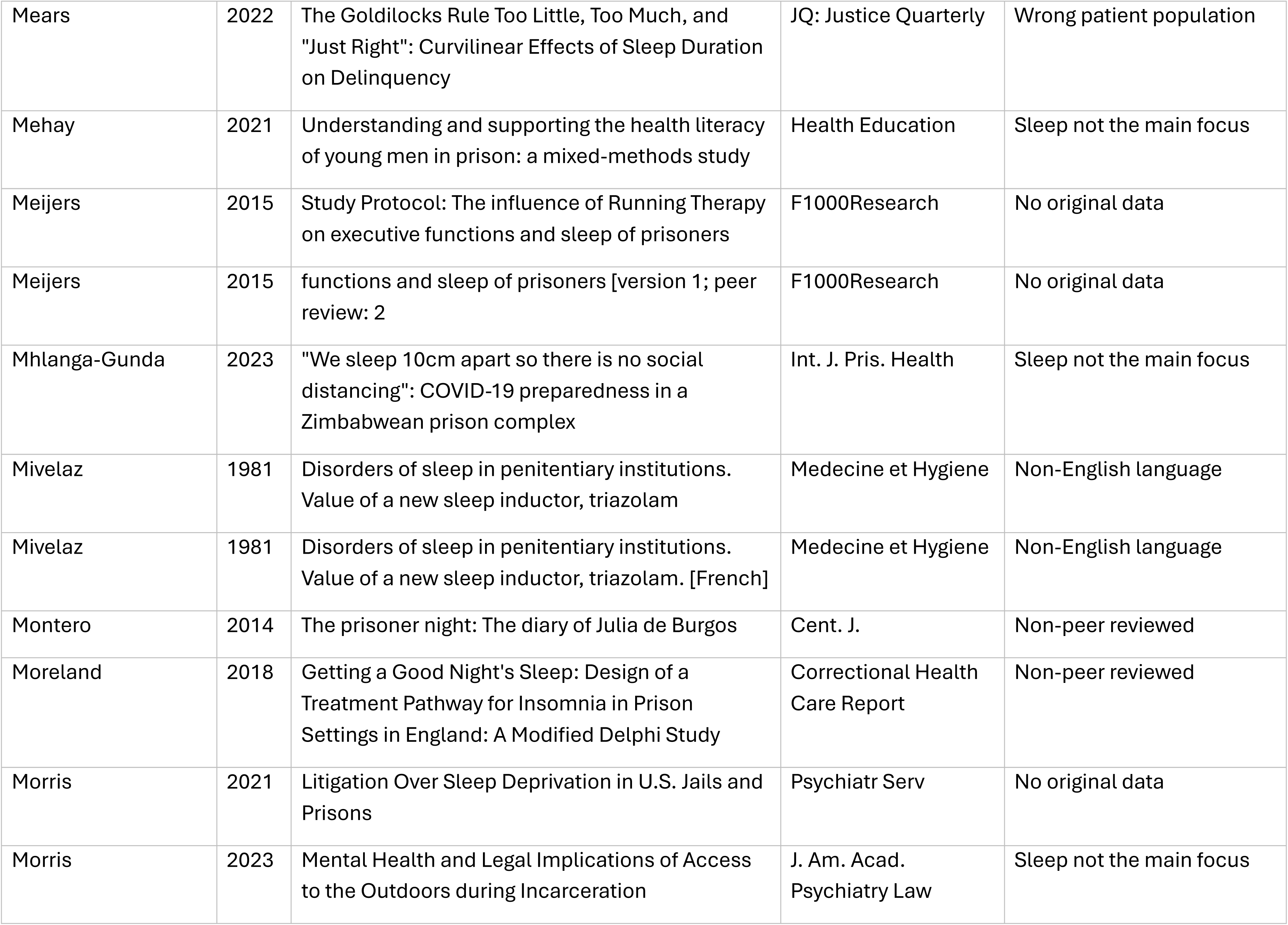
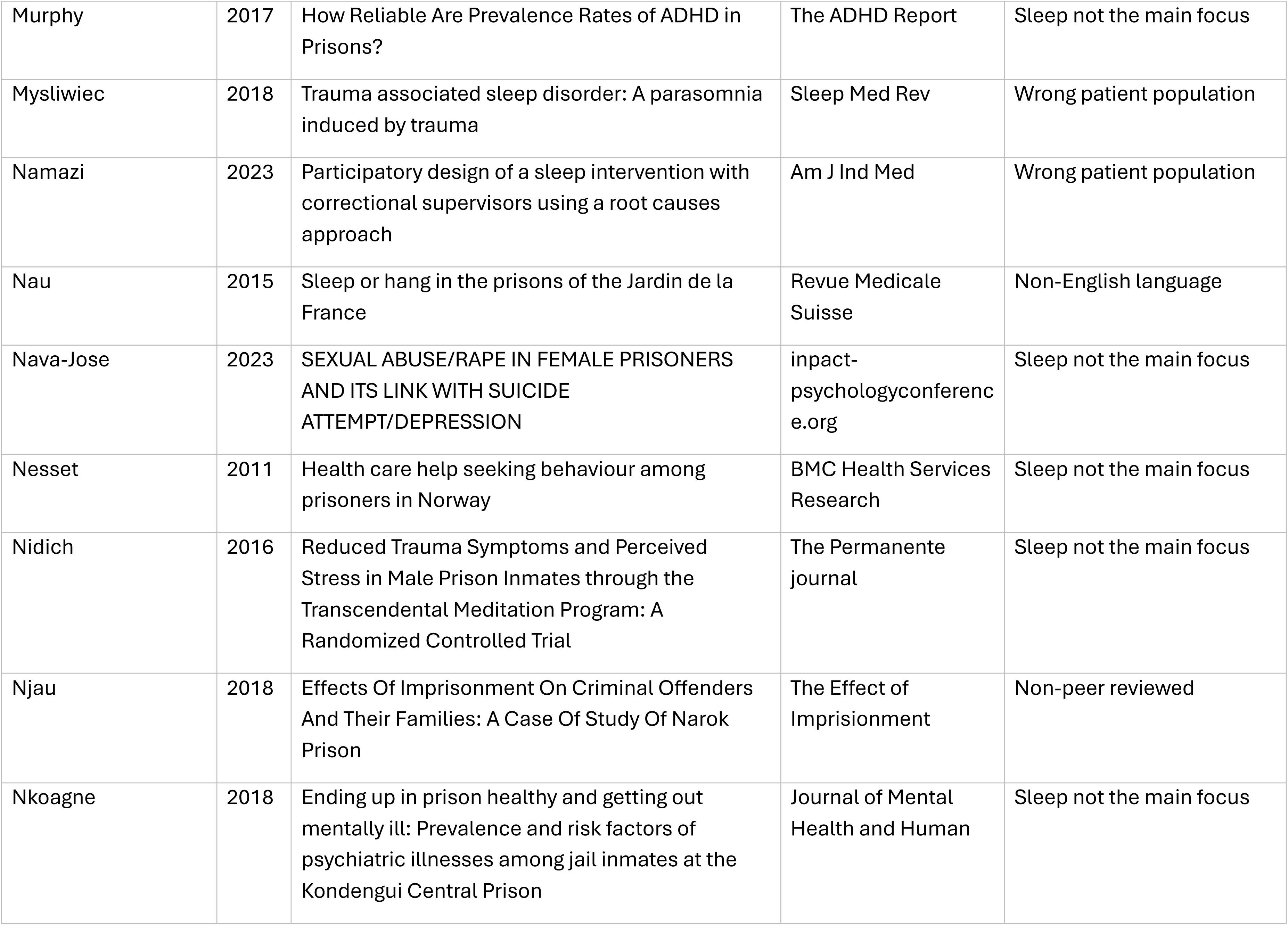
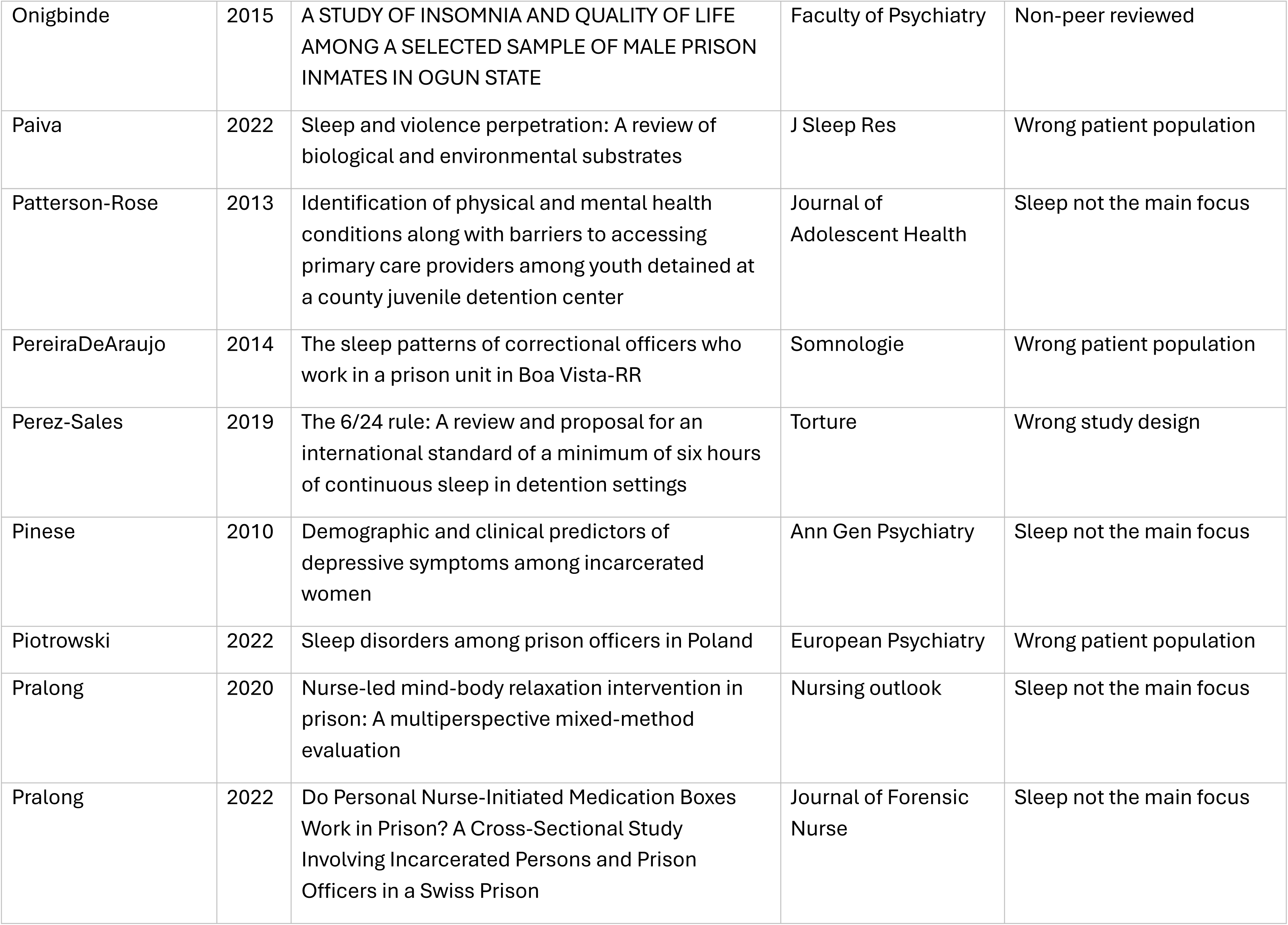
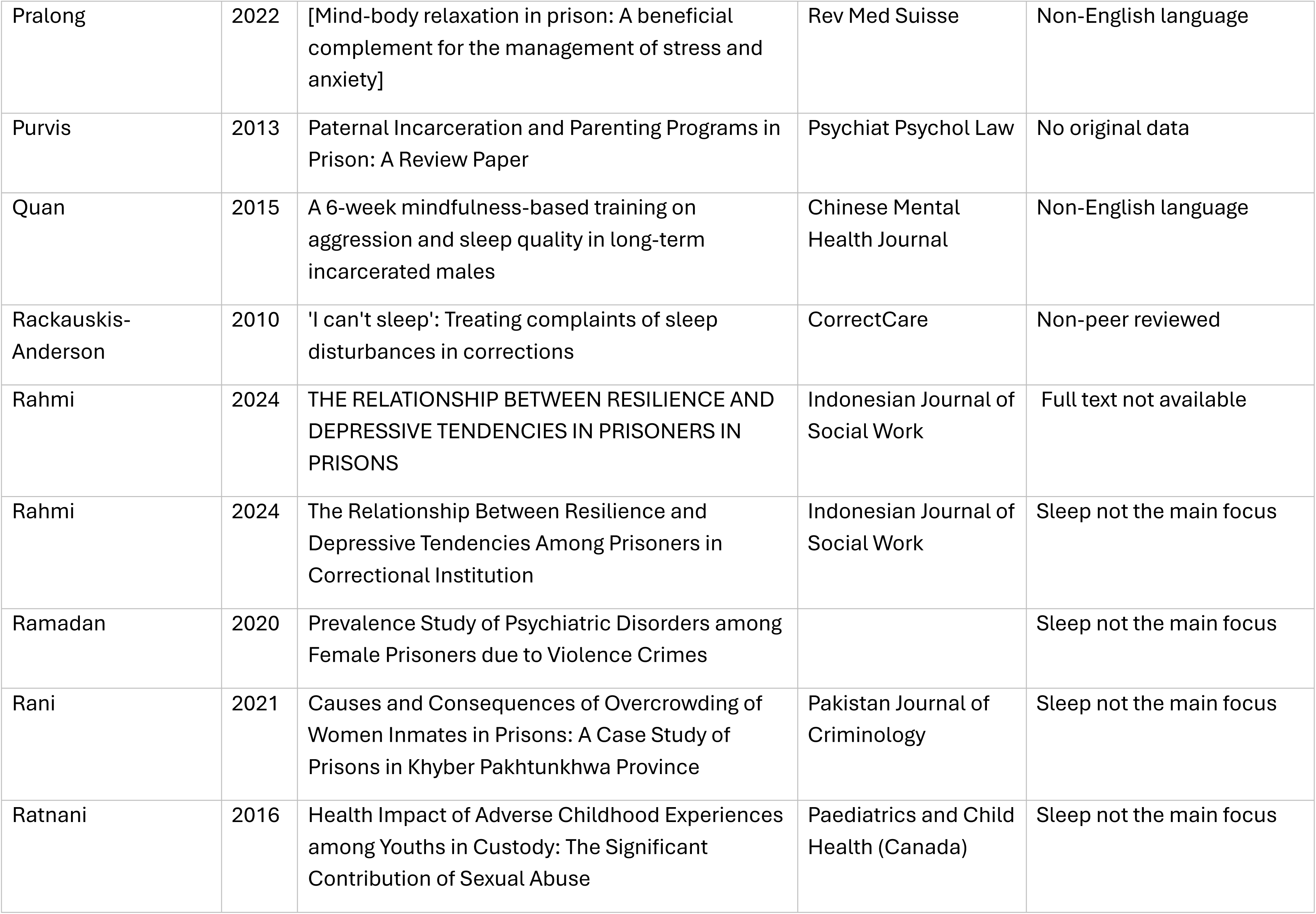
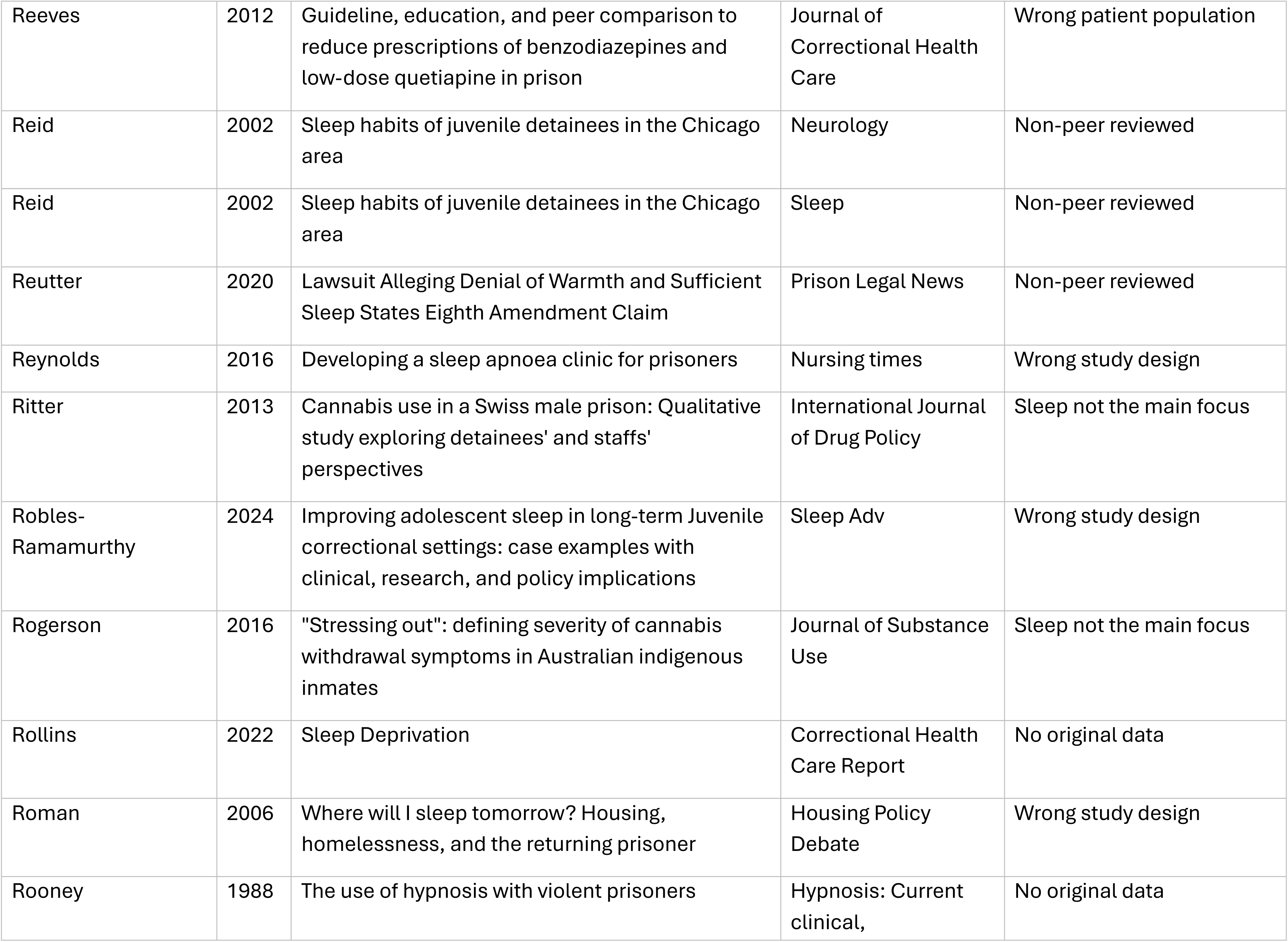
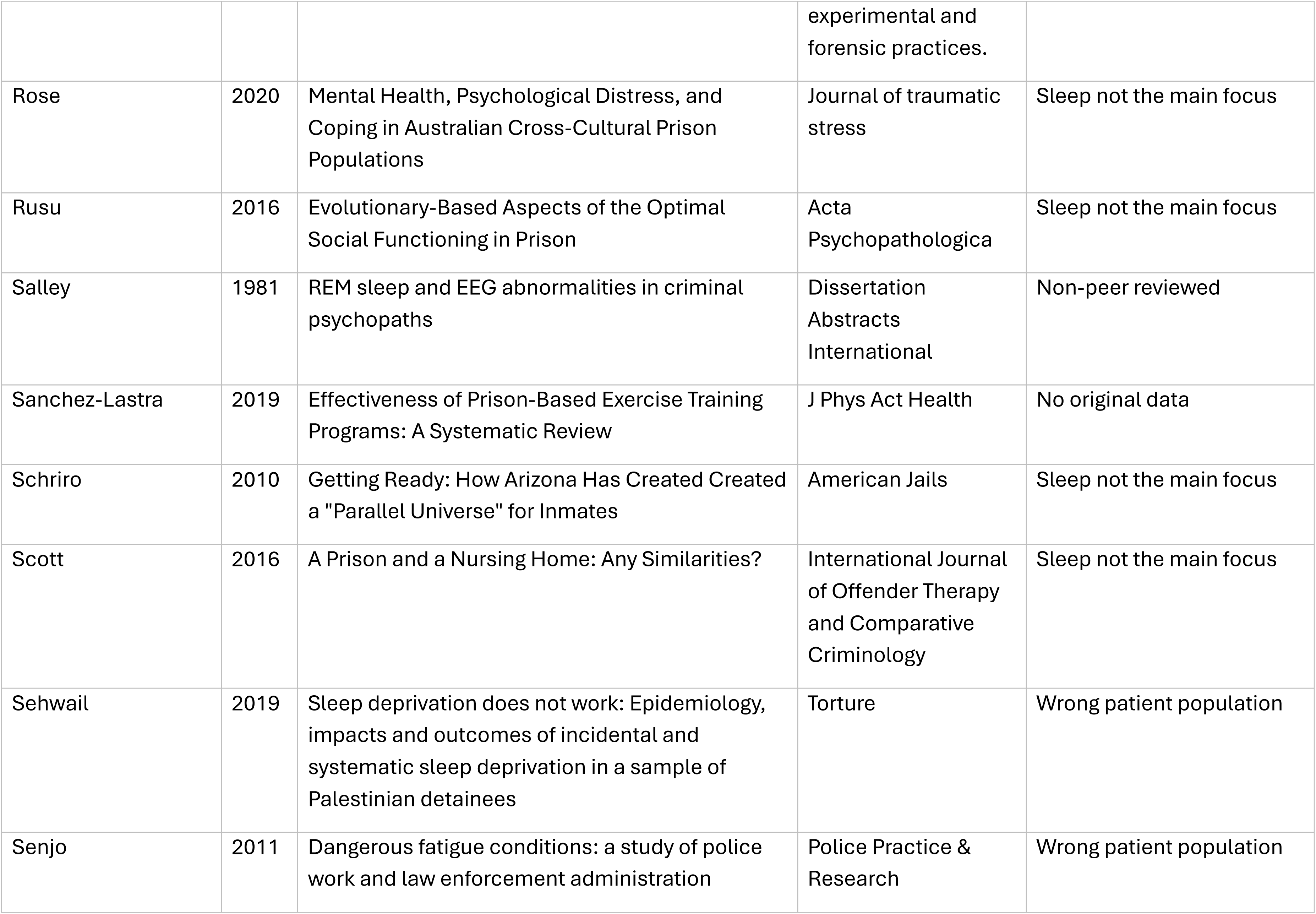
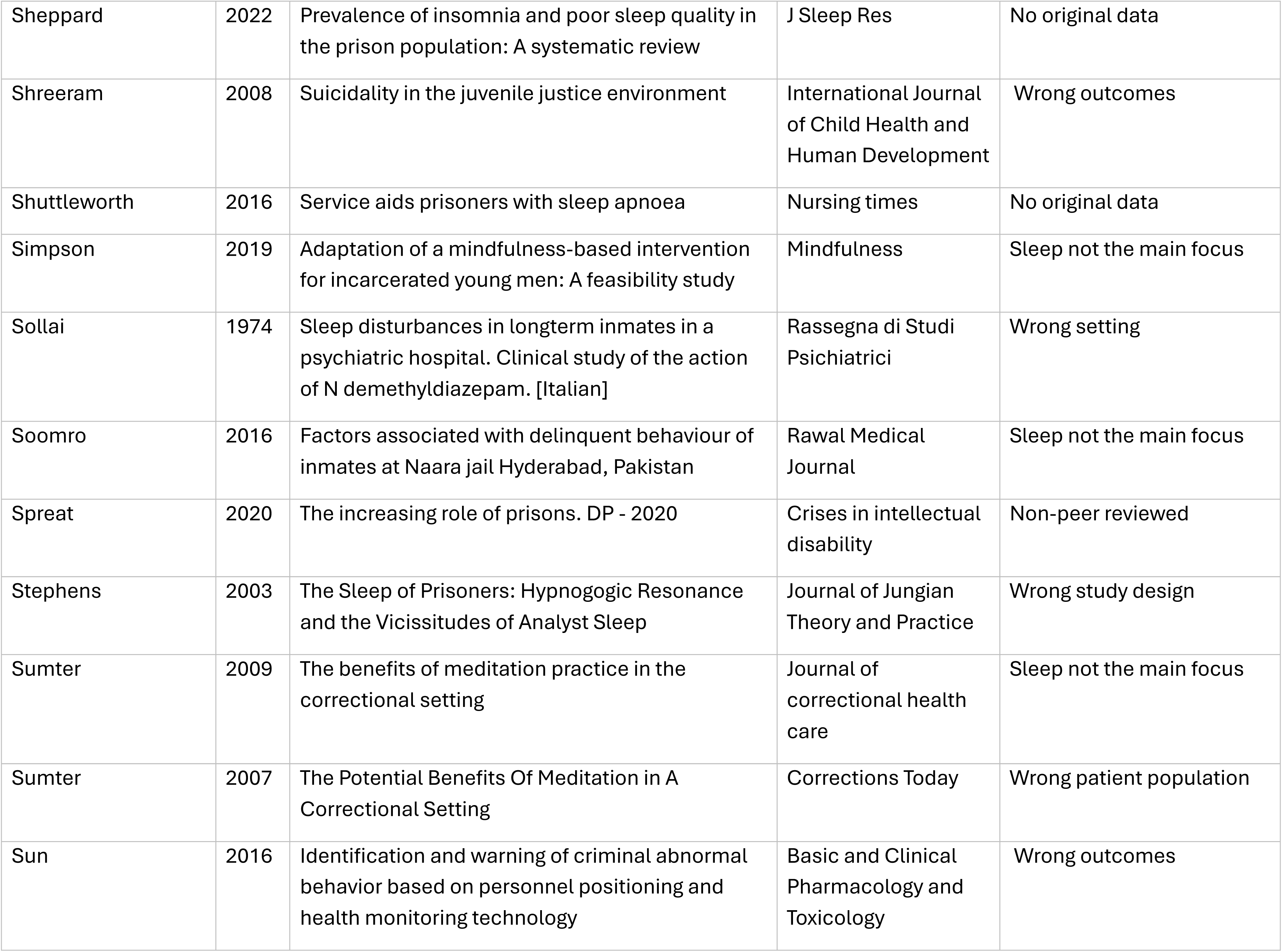
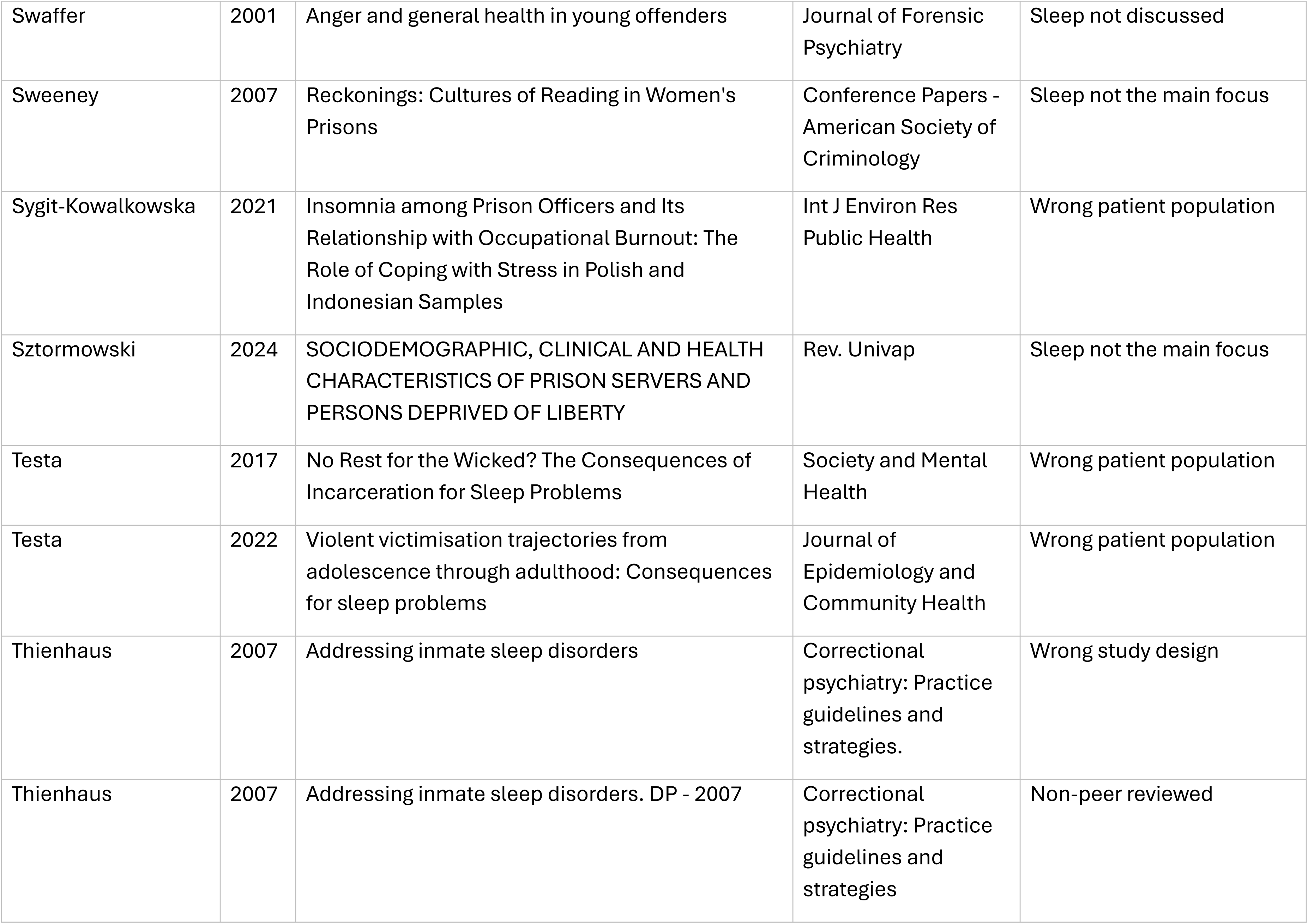
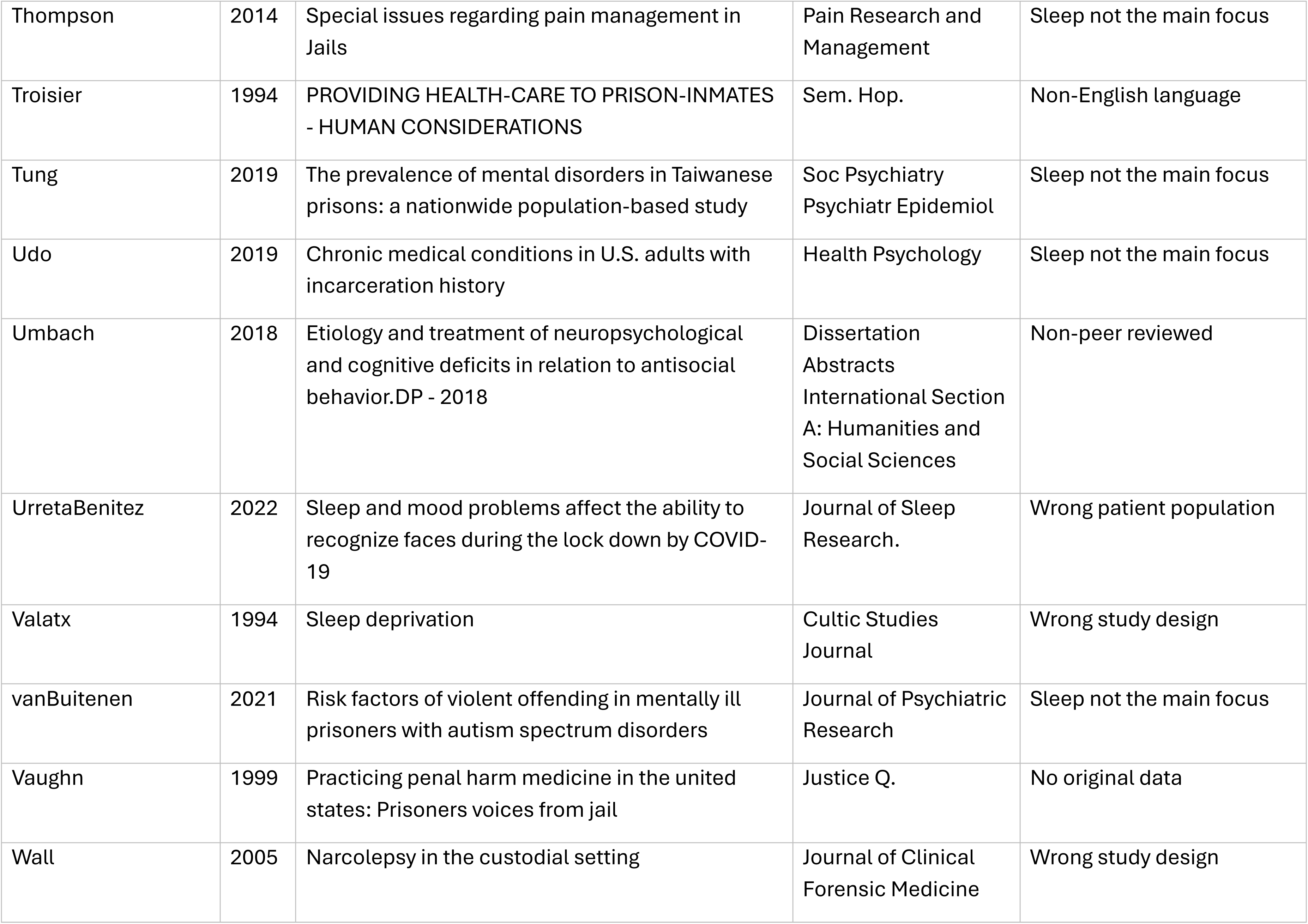
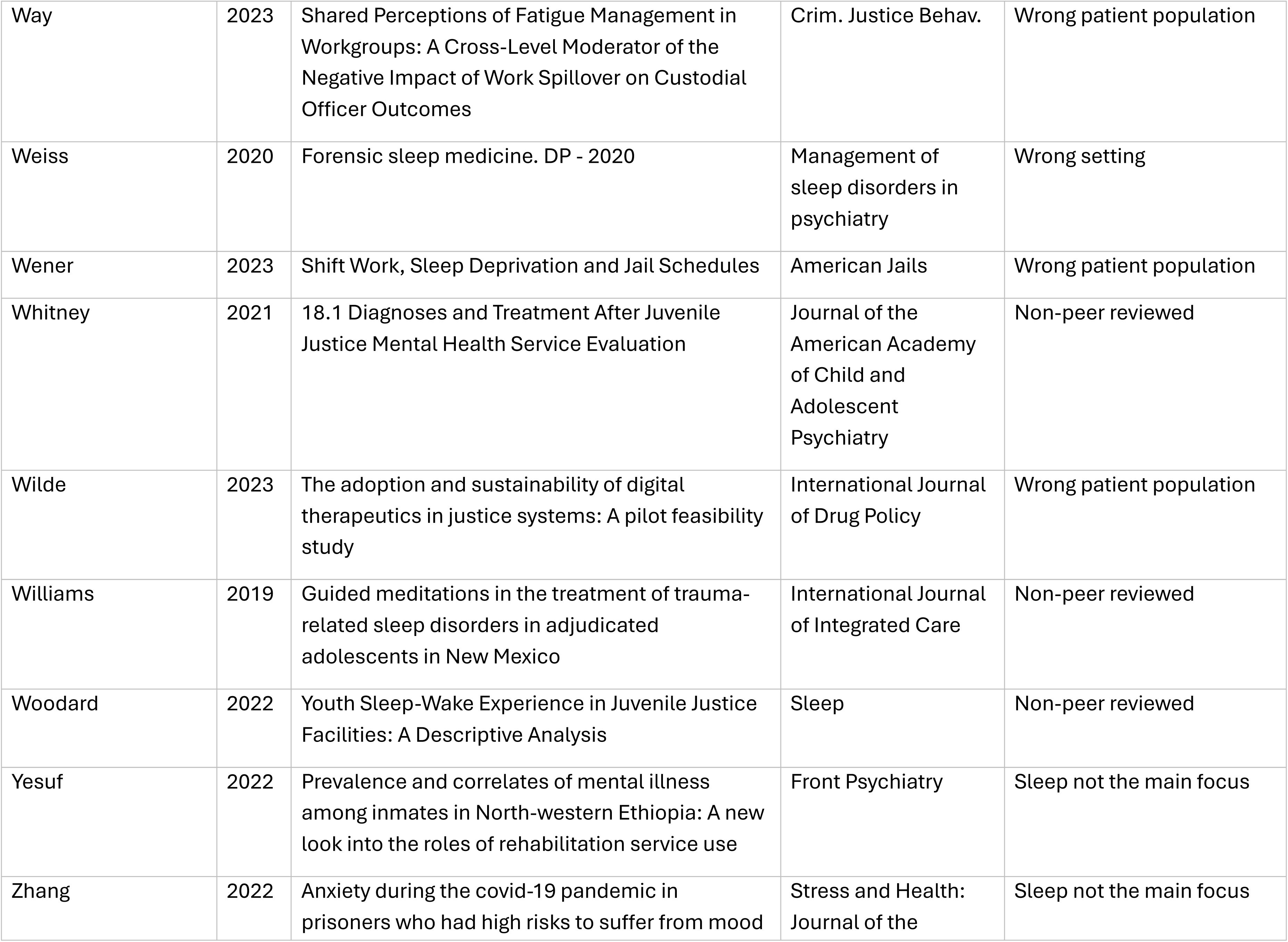
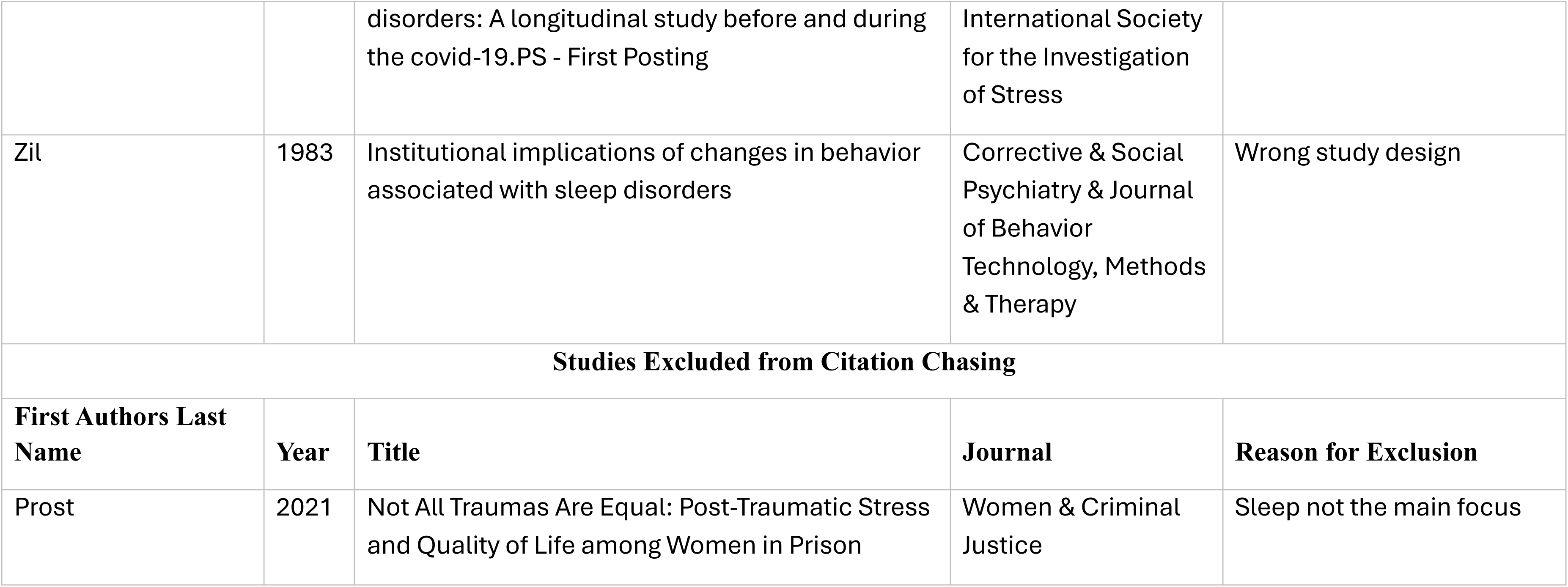
Excluded Studies Table.

| Lead Author | Year | Country | Study Focus | Study Aim | Study Design | Setting/Sample | Data Source | Study Outcomes | Main Findings |
| --- | --- | --- | --- | --- | --- | --- | --- | --- | --- |
| Abdu | 2020 | Ethiopia | Sleep Quality | To assess the prevalence and associated factors of poor quality of sleep among incarcerated people in Mettu town prison. | Cross-sectional study | **Setting:** Prison<br>**Sample size:** 310<br>**Age:** Adults<br>**Sex:** Female (n=19)<br>Male (n=291) | Survey<br>**Measure(s):** Pittsburg Sleep Quality Index | **Sleep Outcomes:** Sleep Quality | 77.1% of participants had poor sleep quality. Being single/widowed, history of previous incarceration, urban residence, illiteracy, lifetime alcohol use significantly associated with poor sleep quality |
| Abukari | 2024 | Ghana | Cardiovascular risk, Sleep duration/problems | Prevalence of cardiovascular disease (CVD) risk factors and sleep | Cross-sectional study | **Setting:** Prison<br>**Sample size:** 134<br>**Age:** Adults<br>**Sex:** Males | Survey<br>Clinical assessment<br>**Measure(s):** Questionnaire (dietary habits, tobacco use, alcohol consumption, sleep behavior, physical activity)<br>Clinical measures: Blood pressure<br>Body mass index (BMI)<br>Serum lipid profile<br>Fasting blood glucose (FBG) | **Sleep Outcomes:** Sleep duration<br>Difficulty sleeping | 48.1% of the participants had abnormal lipid levels. Average sleep duration at night was 5.54 ± 2.07 hours, 89.6% had difficulty sleeping at night, 85% reported overcrowding and 15% reported depression as the reason for their sleeplessness |
| Acar | 2019 | Turkey | Circadian preferences, Sleep Quality, PTSD | To explore whether DSM-5 posttraumatic stress disorder (PTSD) dissociative subtype could be differentiated from non-dissociative PTSD in sleep disturbances, circadian preferences, and posttraumatic cognitions. | Cross-sectional study | **Setting:** Prison<br>**Sample:** 399<br>**Age:** Adults<br>**Sex:** Female (n=10)<br>Male (n=389) | Survey<br>**Measure(s):** Pittsburg Sleep Quality Index; Insomnia; Morningness-Eveningness Questionnaire (MEQ), Dissociative Experiences Scale (DES), PTSD Checklist for DSM-5(PCL-5), Posttraumatic Cognitions Inventory (PTCI) | **Sleep Outcomes:** Sleep Quality<br>Sleep Duration<br>Sleep Latency<br>Sleep Disturbance<br>Sleep Apnea<br>Other Sleep Disorder<br>**Other Study Outcomes:** PTSD, dissociative experiences, post-traumatic cognitions, daytime dysfunction | 59.4% screened positive for DSM-5 PTSD with high prevalence of prior trauma exposure (92.0%). 36.8% had pathological levels of dissociation, 70.4% poor sleep quality, and 37.8% insomnia. |
| Adornetti | 2023 | United States | Juvenile, Sleep Health, Environmental Sleep Factors | To describe sleep and circadian health of adolescents living in detention and treatment facilities. | Cohort study, Qualitative | **Setting:** Juvenile detention/treatment<br>**Sample:** 62<br>**Age:** Juvenile<br>**Sex:** Female (n=9)<br>Male (n=53) | Survey<br>Semi-structured post study interview<br>**Measure(s):**<br>Novel Youth Sleep and Daytime Behavior Questionnaire (Sleep-Wake Behavior Problems Scale, Sleepiness Scale, Richards-Campbell Sleep Questionnaire), Daily sleep diary | **Sleep Outcomes:**<br>Sleep Quality<br>Sleep Duration<br>Sleep Latency<br>Sleep Disturbance<br>Other Sleep Disorder | Youth obtain recommended total sleep time but have twice the recommended sleep latency. Early facility sleep-wake schedules and overhead lights are possible factors. |
| Adri | 2022 | Indonesia | Physical Activity, Sleep Quality |  | Randomized control trial | **Setting:** Prison<br>**Sample size:** 24<br>**Age:** Adolescent<br>**Sex:** Males | Survey<br>**Measure(s):** Adapted-Sleep Quality Scale (SQS) | **Sleep Outcomes:**<br>Sleep Quality | Group based games improved sleep quality in adolescent prisoner population |
| Alaei | 2024 | Iran | Spiritual care, Aromatherapy, Sleep Quality | To assess the effects of spiritual care with and without aromatherapy on sleep quality | Randomized control trial | **Setting:** Prison<br>**Sample size:** 90<br>**Age:** Adult<br>**Sex:** Males | Survey<br>**Measure(s):** Pittsburg Sleep Quality Index | **Sleep Outcomes:**<br>Sleep Quality | Post-intervention there was significant improvement in sleep quality in the spiritual care + aromatherapy group $9.53 \pm 4.06$ to $7.86 \pm 4.09$ ($p=0.002$). The Spiritual Care with Aromatherapy was also showed significant improvement in sleep quality vs. the control group ($p=0.011$) |
| An | 2019 | China | Mindfulness Training | To examine the effects of a 6-week mindfulness training program on mental health, anxiety, depression, aggression, sleep quality, mood states, total mood disturbance, and perceived stress among male Chinese incarcerated people, and to explore whether the intervention effects differed in long-term and short-term incarcerated people. | Randomized control trial | **Setting:** Prison<br>**Sample size:** 83<br>**Age:** Adult<br>**Sex:** Males | Survey<br>**Measure(s):** Pittsburg Sleep Quality Index; FFMQ; SCL-90, Zung SAS, Zung SDS, PSQI, AQ, POMS, CPSS | **Sleep Outcomes:**<br>Sleep Quality<br>**Other Study Outcomes:**<br>Mental health, anxiety, depression, aggression, mood states, total mood disturbance, perceived stress | Mindfulness group showed improvement in mindfulness, aggression, sleep quality, and total mood disturbance and was more effective for long-term incarcerated people. |
| Atan | 2025 | Turkey | Obesity, Cardiometabolic risk | To evaluate the obesity, cardiometabolic disease risk and associated factors in incarcerated men | Cross-sectional study | **Setting:** Prison<br>**Sample size:** 139<br>**Age:** Adult<br>**Sex:** male | Survey<br>Clinical assessment<br>**Measure(s):** Pittsburgh Sleep Quality Index, Clinical measures: Body mass index, waist circumference | **Sleep Outcomes:** Sleep Quality<br>**Other Study Outcomes:** Obesity, BMI, Waist circumference | 53.2% were overweight, 13.7% were obese. Cardiometabolic risk was 47.5% (waist circumference), 53.2% (waist-to-hip ratio) and 73.4% (waist-to-height ratio) |
| Barker | 2016 | United Kingdom | Sleep quality, Sleep quantity, Aggression, Motivation | To examine the association between self-reported sleep quality and quantity and how these relate to aggression motivation and hostile cognition in a sample of incarcerated men sample. | Cross-sectional study | **Setting:** Prison<br>**Sample size:** Study 1 (n=95)<br>Study 2 (n=141)<br>**Age:** Adult<br>**Sex:** Male | Survey<br>**Measure(s):** Pittsburg Sleep Quality Index; Direct and Indirect Prisoner Checklist-Revised (Study1); Puzzle test; Hostile Interpretation Questionnaire, reactive-proactive Questionnaire (Study 2) | **Sleep Outcomes:** Sleep Quality<br>Sleep Duration<br>**Other Study Outcomes:** Aggression, non-aggression, victimization, perpetration, motivation, hostile vs. prosocial attribution bias | Participants with perceived poor sleep report greater reactive and proactive aggression |
| Camplain | 2022 | United States | Sleep quality, Housing status | To investigate sleep quality among individuals incarcerated in a rural county jail, by housing status before incarceration. | Cross-sectional study | **Setting:** Jail<br>**Sample:** 194<br>**Age:** Adult<br>**Sex:** Female (n=42)<br>Male (n=152) | Survey<br>**Measure(s):** Health survey with 2 questions about sleep quality | **Sleep Outcomes:** Sleep Quality | Non-permanent housing before incarceration predicted lower prevalence of worsening sleep quality while incarcerated compared to those with permanent housing. |
| Carli | 2011 | Italy | Insomnia, Suicidal behavior | To examine for a relationship between insomnia and suicidal behavior in a large | Cross-sectional study | **Setting:** Prison<br>**Sample size:** n=1427<br>**Age:** Adults<br>**Sex:** Male | Survey | **Sleep Outcomes:** Sleep Quality<br>Insomnia | Insomnia was significantly and independently associated with a history of attempting suicide |
|  |  |  |  | group of 1420 incarcerated people. |  |  | **Measure(s):** Pittsburgh Sleep Quality Index, Hamilton Depression Rating Scale | **Other Study Outcomes:** neuroticism, suicidal behavior, anger/impulsive aggression, violent behavior |  |
| Chinichian | 2020 | Iran | Substance use disorder, Sleep quality | To evaluate sleep quality and the related risk factors in incarcerated people with Substance Use Disorder in Iran. | Cross-sectional study | **Setting:** Prison<br>**Sample size:** 352<br>**Age:** Adults<br>**Sex:** Male | Survey<br>**Measure(s):** Pittsburg Sleep Quality Index; PSQI-P (Persian version of PSQI) | **Sleep Outcomes:** Sleep Quality<br>**Other Study Outcomes:** Substance Use | Poor sleep quality associated with total imprisonment duration, time till end of term, having a tattoo, history of sedative use, use of more than one substance, and use of more than one sedative. |
| Cope | 2003 | United Kingdom | Cannabis use, Prison length of stay, Time | To explores perspectives of time amongst incarcerated young men in prison and reflects on their cannabis use as a strategy that enabled them to manage their long sentences. | Qualitative longitudinal study | **Setting:** Prison<br>**Sample size:** 30<br>**Age:** Adolescents and young adults<br>**Sex:** Males | Interview<br>**Measure(s):** Unstructured qualitative interviews, Records, Staff interviews | **Sleep Outcomes:** Sleep Quality<br>Sleep Duration<br>Insomnia<br>**Other Study Outcomes:** Psychological coping mechanisms | Prisons present paradox where some aspects are highly structured while others are inherently uncertain (e.g. interactions with incarcerated people/staff, copious unstructured time). |
| D'Aurizio | 2020 | Italy | Anxiety, Depression, Sleep quality | To explore sleep quality and sleep habits in an Italian prison ward with three different security levels, and to attempt to clarify how anxiety state and the total time spent in prison can moderate insomnia complaints | Cross-sectional study | **Setting:** Prison<br>**Sample size:** 129<br>**Age:** Adults<br>**Sex:** Male | Survey<br>**Measure(s):** Pittsburg Sleep Quality Index; Insomnia Index; Beck Depression Inventory II; State-Trait Anxiety Inventory | **Sleep Outcomes:** Sleep Quality<br>Insomnia<br>**Other Study Outcomes:** Depression<br>Anxiety | Poor sleep quality, insomnia, mild to moderate depressive symptoms, and clinically significant anxiety symptoms were presented. |
| D'Aurizio | 2023 | Italy | Sleep quality, Sleep quantity | To assess the quality and quantity of sleep by using, for the first time, actigraphy. | Cross-sectional study | **Setting:** Jail<br>**Sample size:** 60 | Survey<br>Actigraphy<br>**Measure(s):** Actiwatch | **Sleep Outcomes:** Sleep Quality<br>Sleep Duration | Incarcerated people exhibited a longer sleep latency and a shorter sleep than controls, confirming |
|  |  |  |  |  |  | (30 incarcerated, 30 volunteers-control group)<br>**Age:** Adult<br>**Sex:** Male | Pittsburg Sleep Quality index, Insomnia Index, PSQI, Beck Depression Inventory II Edition, State-Trait Anxiety Inventory | Sleep Latency<br>Sleep Disturbance<br>Insomnia<br>**Other Study Outcomes:**<br>Depression<br>Anxiety | subjective measures. |
| Dewa | 2017 | United Kingdom | Insomnia | To investigate the prevalence of insomnia and identify associated demographic, clinical and forensic risk factors in adult incarcerated people in England. | Cross-sectional study | **Setting:** Prison<br>**Sample size:** 237<br>**Age:** Adults<br>**Sex:**<br>Females (n=119)<br>Male (n=118) | Survey<br>**Measure(s):** Pittsburgh Sleep Quality Index; Sleep Hygiene Index; Dysfunctional Beliefs and Attitudes About Sleep (DBAS-16); Sleep Disorders Screener; Sleep Condition Indicator; Brief Psychiatric Rating Scale (BRPS); Prison Environment Sleep questionnaire (PESQ) | **Sleep Outcomes:**<br>Sleep Quality<br>Sleep Latency<br>Insomnia | 61.6% of prison participants had possible insomnia, compared to 45.7% of a large UK general population sample |
| Dewa | 2018 | United Kingdom | Insomnia, Insomnia Treatment, Prison-based Intervention | To design a treatment pathway for insomnia in prisons | Modified Delphi study | **Setting:** Prison<br>**Sample size:**<br>Round 1 = 15 stakeholders; Rounds 2 and 3 = 13 stakeholders<br>Incarcerated people, sleep researchers, prison staff were included in the study.<br>**Age:** Adults<br>**Sex:** Male<br>Female | Interview, Survey<br>**Measure(s):**<br>Modified Delphi technique | **Sleep Outcomes:**<br>Sleep Quality<br>Insomnia | A standardized stepped-care pathway for insomnia in prison populations, addressing transition, assessment, and short- and long-term treatment, was developed with high stakeholder consensus (>80%) despite initial contention to promote early detection of insomnia and reduce sleep-related complaints. |
| Dewa | 2024 | United Kingdom | Peer Support, Environmental Aids, Cognitive Behavioral Therapy for Insomnia | To assess the feasibility of a treatment pathway for insomnia in a high-secure prison to inform a future randomized controlled trial (RCT) and initial efficacy data for sleep and mental health outcomes | Cohort study | **Setting:** Prison<br>**Sample size:** 36<br>**Age:** Adults<br>**Sex:** Male | Interview<br>Survey<br>Actigraphy<br>**Measure(s):** Actiwatch<br>Pittsburg Sleep Quality Index; Insomnia Index; Epworth Sleepiness Scale; | **Sleep Outcomes:**<br>Sleep Quality<br>Sleep Durations<br>Sleep Latency<br>Sleep Disturbance<br>Insomnia | Large treatment benefits found for insomnia severity, depression, anxiety, well-being, and cognitive functioning, with a medium benefit on suicidal ideation. No overall change in actigraphy-measured sleep. |
|  |  |  |  |  |  |  | Sleep condition indicator; Warwick-Edinburg mental well-being scale; PHQ-0, Generalized anxiety disorder assessment, Beck scale for suicide ideation (BSS); PROMIS-10; Flinders Fatigue Scale (FSS); Cognitive Failures Questionnaire (CFS) | **Other Study Outcomes:** Well-being, Mood, Anxiety, Suicidality, Overall health, Sleepiness, Fatigue, Cognitive functioning |  |
| Dos Santos | 2017 | Brazil | Sleep<br>Mental health | To identify the factors affecting the mental health of women in a prison in Rio de Janeiro. | Cross-sectional study<br>Qualitative | **Setting:** Prison<br>**Sample size:** n=40<br>**Age:** Adult<br>**Sex:** Females | Qualitative interviews<br>Medical record review | **Sleep Outcomes:** Sleep disturbances<br>Sleep medications<br>Sleep as an escape<br>Environment and sleep<br>**Other Outcomes:** Mental health | The women identified altered sleep patterns as a factor that impacted their mental health during incarceration. They used psychotropic medication to help them sleep and for some sleep was a way to escape. |
| Eichelman | 2021 | United States | Prazosin Treatment, Traumatic Dreams, PTSD | To evaluate patient and clinician assessment of the efficacy of prazosin in the treatment of recurring traumatic dreams in a female prison population. | Retrospective cohort study | **Setting:** Prison<br>**Sample size:** 81<br>**Age:** Adults<br>**Sex:** Females | Survey<br>**Measure(s):** Clinical Global Impression; Clinician Administered PTSD Scale, Pittsburgh Sleep Quality Index (PSQI) | **Sleep Outcomes:** Sleep Quality<br>Sleep Latency<br>Nightmares | Percent of participants waking from dream decreased from 100% to 57% after prazosin treatment |
| Eichelman | 2022 | United States | Traumatic Dreams, PTSD | To (a) ascertain the incidence and character of traumatic dreams in a women's prison, (b) determine whether this was associated strongly with a diagnosis of posttraumatic stress disorder (PTSD); and (c) determine whether this symptom was being treated. | Cross-sectional study | **Setting:** Prison<br>**Sample size:** 483<br>**Age:** Adults<br>**Sex:** Females | Survey<br>**Measure(s):** Clinician Administered PTSD Scale | **Sleep Outcomes:** Sleep<br>Disturbance<br>Nightmares<br>**Other Study Outcomes:** Post-traumatic stress disorder (PTSD) | 282 (74%) reported traumatic dreams, of which 85 (30%) were nightly and 107 (38%) weekly. |
| Elger | 2003 | Switzerland | Insomnia | To find out whether subjective sleep quality improves during time and which factors influence improvement. | Cohort study | **Setting:** Prison<br>**Sample size:** 52<br>**Age:** Adult<br>**Sex:** Female (n=3)<br>Male (n=37) | Survey<br>**Measure(s):** Pittsburg Sleep Quality Index; General Health Questionnaire | **Sleep Outcomes:** Insomnia<br>**Other Study Outcomes:** anxiety, depression, somatic symptoms | PSQI insomnia significantly improvements in the first one to two weeks of incarceration but stagnates after |
| Elger | 2004 | Switzerland | Insomnia | To determine the prevalence of insomnia complaints and possible causes among incarcerated people. | Prospective study | **Setting:** Prison<br>**Sample size:** 215<br>**Age:** Adults<br>**Sex:** Males (n=197)<br>Females (n=18) | Survey<br>Medical records review | **Sleep Outcomes:** Insomnia | Insomnia is frequent and chronic, causing medical and psychological problems. |
| Elger | 2004 | Switzerland | Insomnia | To examine the clinical management of insomnia in non-substance misusing people in prison. | Retrospective study | **Setting:** Prison<br>**Sample size:** 112<br>**Age:** Adults<br>**Sex:** Female (n=11)<br>Male (n=101) | Chart review | **Sleep outcomes:** Insomnia<br>Medications prescribed for sleep | Patients were overwhelmingly prescribed hypnotics for insomnia (n=111 participants) with 80% receiving benzodiazepines or Zolpidem. More than 60% of participants reported insomnia lasting more than 3 weeks. 37% showed completed improvement, while 30% had partial improvement and 18% no improvement. 55% of the 112 patients were still being prescribed hypnotics when they left prison. |
| Elger | 2009 | Switzerland | Insomnia Risk Factors, Daily Activities, Stress | To compare how incarcerated people complaining of insomnia and incarcerated people without sleep problems describe their daily activities, stress factors and the perceived reasons for their sleep quality in prison. | Cross-sectional study | **Setting:** Prison<br>**Sample:** 163<br>**Age:** Adults<br>**Sex:** Female (n=12)<br>Male (n=151) | Survey<br>**Measure(s):** Pittsburg Sleep Quality Index; GHQ | **Sleep Outcomes:** Sleep Quality<br>Insomnia | Those with insomnia and those reporting good sleep had significant differences in their daily activities and stressful events. Exercise, medical care, and psychological support can help reduce sleep problems. |
| Elger | 2009 | Switzerland | Insomnia, Subjective Sleep Quality | To measure the subjective sleep quality of incarcerated people complaining of insomnia and to compare it to the subjective sleep quality of incarcerated people who report "good sleep", in order to determine factors that predict insomnia and severity of insomnia. | Cross-sectional study | **Setting:** Prison<br>**Sample size:** 147<br>**Age:** Adults<br>**Sex:** Female (n=10)<br>Male (n=137) | Interview Survey<br>**Measure(s):** Pittsburg Sleep Quality index; Goldberg's General Health Questionnaire | **Sleep Outcomes:**<br>Sleep Quality<br>Sleep Duration<br>Insomnia | Insomniacs and good sleepers present significant differences in subjective sleep quality. Causes of insomnia are listed. |
| Fakorede | 2015 | Nigeria | Sleep Efficiency, Sleep Quality, Sleep Pattern | To determine the sleep efficiency and quality of Nigerian incarcerated people as well as a reversal in their sleep pattern. | Cross-sectional study | **Setting:** Prison<br>**Sample size:** 300<br>**Age:** Adults<br>**Sex:** Male | Survey<br>**Measure(s):** Pittsburg Sleep Quality Index | **Sleep Outcomes:**<br>Sleep Quality<br>Other Sleep Disorder | Approximately two-thirds (67%) of the respondents were awaiting trial, and this group was significantly younger, lived in more crowded cells, shared sleeping materials, and had higher rates of psychoactive substance use compared to incarcerated people convicted of a crime. Most incarcerated people awaiting trial (95%) had spent less than five years in prison, while only 20% of incarcerated people convicted of a crime had the same duration of imprisonment. Sleep-related issues were prevalent, with 94% of respondents experiencing sub-normal sleep efficiency and 37.7% reporting poor sleep quality, though only 1% experienced a reversal of their sleep-wake cycle. Among the factors investigated, self-perception of a sleep problem was the only significant correlate of poor sleep quality. |
| Fakorede | 2021 | Nigeria | Insomnia, Quality of Life | To determine the prevalence of insomnia as well as its association with the quality of life of incarcerated people in a Nigerian prison. | Cross-sectional study | **Setting:** Prison<br>**Sample size:** 300<br>**Age:** Adults<br>**Sex:** Male | Survey<br>**Measure(s):** Insomnia module for the Schedule for Clinical Assessment in | **Sleep Outcomes:**<br>Insomnia | 45.7% of respondents met insomnia diagnostic criteria. Insomnia and some sleep-related variables were significantly associated with quality of life. |
|  |  |  |  |  |  |  | Neuropsychiatry, Brief World Health Organization Quality of Life questionnaire |  |  |
| Fakorede | 2023 | Nigeria | Family Support, Sleep Environment, Sleep Quality | To determine the association between prison circumstances, perceived family support and sleep quality among men incarcerated at the Ibara prison, Ogun State. | Cross-sectional study | **Setting:** Prison<br>**Sample size:** 300<br>**Age:** Adults<br>**Sex:** Male | Survey<br>**Measure(s):** Pittsburgh Sleep Quality Index | **Sleep Outcomes:**<br>Sleep Quality<br>Sleep Duration<br>Sleep Latency<br>Sleep Disturbance<br>Other Sleep Disorder | Overcrowding, inappropriate sleeping materials, and self-awareness of sleep problem associated with poor sleep quality, but no predictor of the latter was identified. |
| Ferszt | 2015 | United States | Mindfulness Intervention, Stress, Anxiety, Depression | To examine the impact of mindfulness-based program on the stress, anxiety, depression, and sleep of women | Pre-post Non-Experimental | **Setting:** Prison<br>**Sample size:** 33<br>**Age:** Adults<br>**Sex:** Female | Survey<br>**Measure(s):** Pittsburg Sleep Quality Index; Center for Epidemiological Study; State Trait Anxiety Scale; Perceived Stress Scale | **Sleep Outcomes:**<br>Sleep Quality | First group showed significant decrease in stress and depression scores. Second group was decreased scores but was not significant. |
| Geng | 2021 | China | Sleep Problems, Depression, PTSD | To investigate the prevalence and correlates of sleep problems in a large prison in China. | Cross-sectional study | **Setting:** Prison<br>**Sample size:** 1491<br>**Age:** Adults<br>**Sex:** Male | Survey<br>**Measure(s):** Sleep duration and sleep problems assessed based on author's prior studies, Life Events Checklist, PCL-5, PHQ-9 | **Sleep Outcomes:**<br>Sleep Quality<br>Sleep Duration<br>Sleep Latency<br>Sleep Disturbance<br>Other Sleep Disorder | Short sleep, insomnia, and poor sleep quality highly co-occurred |
| Getachew | 2020 | Ethiopia | Sleep Quality associated factors | To assess the quality of sleep and related factors among incarcerated people of the Diredawa correctional facility, Diredawa, eastern Ethiopia. | Cross-sectional study | **Setting:** Prison<br>**Sample size:** 300<br>**Age:** Adults<br>**Sex:** Female (n=28)<br>Male (n=393) | Survey<br>**Measure(s):** Pittsburg Sleep Quality Index; Patient Health Questionnaire-9; Sleep Hygiene Index<br>Generalized Anxiety Disorder scale (GAD-7) | **Sleep Outcomes:**<br>Sleep Quality | Depression; cigarettes, cannabis, and hashish use; coffee, poor sleep hygiene, and crime types were associated with poor sleep. |
| Goudard | 2017 | France | Sleep Disorders, Therapeutic Management | To examine sleep disorders and therapeutic management. | Cross-sectional study | **Setting:** Prison<br>**Sample size:** 358<br>**Age:** Adults<br>**Sex:** Female (n=39)<br>Male (n=319) | Survey<br>Prescription review | **Sleep Outcomes:**<br>Sleep Quality<br>Sleep Duration<br>Sleep Disturbance | More than half of participants (56%) were not satisfied with their sleep. Most patients reported that their sleeping problems began or worsened after incarceration. A quarter of |
|  |  |  |  |  |  |  |  | Insomnia | the incarcerated people were following a hypnotic treatment, and most of these treatments began in prison. Only 42% of patients were satisfied with its effectiveness. |
| Habibzadeh | 2024 | Iran | Well-being, Emotion Regulation, Sleep Quality, Substance use | To evaluate the effectiveness of yoga training on psychological well-being, emotion regulation, and sleep quality. | Quasi-experimental study | **Setting:** Prison<br>**Sample size:** 30<br>**Age:** Adults with substance use<br>**Sex:** Not reported | Survey<br>**Measure(s):** Pittsburgh Sleep Quality Index Emotion regulation scale<br>Psychological well-being scale | **Sleep Outcomes:** Sleep Quality<br>**Other Study Outcomes:** Psychological well-being<br>Emotional Regulation | Yoga training significantly impacted psychological well-being, emotional regulation, and sleep quality of incarcerated adults with a history of drug abuse (P<0.05). |
| Haghighat | 2014 | Iran | Anxiety, Substance Abuse, Sedative Drugs, Sleep Disorders | To investigate the relationship of anxiety, sleep disorders and drug abuse with sedative drugs in male patients of Semnan prison. | Cross-sectional study | **Setting:** Prison<br>**Sample size:** 123<br>**Age:** Adult<br>**Sex:** Male | Survey<br>**Measure(s):** Pittsburg Sleep Quality Index, Spiel Berger State-Trait Anxiety Questionnaires | **Sleep Outcomes:** Sleep Quality<br>Sleep Duration<br>Sleep Latency<br>Sleep Disturbances<br>Other Sleep Disorder<br>**Other Study Outcomes:** Anxiety | A correlation between trait (but not state) anxiety and sedative drug use was approved. |
| Harner | 2014 | United States | Sleep Apnea, Sleep Quality | To describe incarcerated women's sleep quality; document incarcerated women's risk for sleep apnea; and identify other factors that contribute to poor sleep quality during incarceration. | Cross-sectional study, Qualitative | **Setting:** Prison<br>**Sample size:** 438<br>**Age:** Adult<br>**Sex:** Female | Survey<br>**Measure(s):** Pittsburg Sleep Quality Index; Multivariate Apnea Prediction Score | **Sleep Outcomes:** Sleep Quality<br>Sleep Duration<br>Sleep Latency<br>Sleep Disturbance<br>Sleep Apnea<br>Insomnia | 72% of the sample met the PSQI criteria for “poor sleepers. Approximately 10% of sample had probability for sleep apnea higher than .50 |
| Harty | 2010 | United States | Borderline Personality, Psychopathy, Antisocial | To investigate the relationship between Borderline Personality Disorder (BPD) and sleep | Cross-sectional study | **Setting:** Jail<br>**Sample size:** 513<br>**Age:** Adult<br>Female (~359) | Survey<br>**Measure(s):** | **Sleep Outcomes:** Sleep problems | Borderline personality disorder, antisocial personality, cocaine and opiate dependence, depression, and psychopathy |
|  |  |  | Personality, Sleep Problem | problems, independent of depression and substance dependence |  | Male (~154) | Created a five item Sleep Problems Index from sleep questions in the PAI; Personality Assessment Inventory (PAI), Psychopathy Checklist-Screening Version (PCL:SV) | Sleep disturbance<br>**Other Study Outcomes:** Depression Personality Disorders | show significant positive correlation with sleep problems |
| Ireland | 2006 | United Kingdom | Aggression, Anger, Impulsivity, Sleep Problems | To examine both the quantity and quality of sleep reported by male adolescents detained in prison, with a focus on exploring the association between the quantity and quality of sleep with aggression, impulsivity, or anger. | Cross-sectional study | **Setting:** Prison<br>**Sample size:** 184<br>**Age:** Adolescents/Young Adult (14-20)<br>**Sex:** Males | Survey<br>**Measure(s):** Sleep Complaints Scale (SCS), Sleep Behavior in and before prison; Aggression Questionnaire (AQ), The Anger Management Assessment Questionnaire (AMA); The Barratt Impulsivity Scale-II (BIS-II) | **Sleep Outcomes:** Sleep Quality Sleep Duration Sleep Latency Sleep Disturbances Sleep Apnea Sleep Disorder<br>**Other Study Outcomes:** Aggression, anger, impulsivity | Overall aggression is predictive of sleep quantity and quality. Hostility is the most important component of aggression to account for. |
| Jacob | 2019 | India | Anxiety, Depression, Insomnia, Muscle x | To examine efficacy of progressive muscle relaxation on irritability, depression, anxiety, and insomnia in opioid dependent incarcerated patients | Quasi-randomized control trial | **Setting:** Psychiatric unit during incarceration<br>**Sample size:** 54<br>**Age:** Adult<br>**Sex:** Male | Survey<br>**Measure(s):** Irritability, Depression, Anxiety Scale (IDAS), Athens Insomnia Scale (AIS) | **Sleep Outcomes:** Insomnia<br>**Other Study Outcomes:** irritability, depression, anxiety | Progressive muscle relaxation produced significant difference in pre and post scores for irritability, anxiety, depression and insomnia. |
| Johnson | 2019 | Canada | Daily Movement Behaviors, Physical Activity, Screen Time, Sleep, Weight Gain | To examine how 24-h movement/nonmovement behaviors contribute to weight gain during incarceration. | Retrospective cohort study | **Setting:** Prison<br>**Sample size:** 754<br>**Age:** Adult<br>**Sex:** Female (n=82), Male (n=672) | Survey<br>Clinical assessment<br>**Measure(s):** Canadian Health Measures Survey<br>Clinical measures: Weight, BMI, waist circumference | **Sleep Outcomes:** Sleep<br>**Other Study Outcomes:** Changes in BMI, waist circumference, weight | Incarcerated people engaged in at least 60 minutes of daily physical activities gained less weight compared to non-exercising incarcerated people. Sleep and screen time were not associated with weight gain outcomes. |
| Kerekes | 2017 | Sweden | Anger, Anxiety, Depression, Stress, Yoga | To investigate effect of yoga in the reduction of depressive symptoms, anxiety, stress, anger, and increased ability of behavioral control In correctional settings. | Randomized Control Trial | **Setting:** Prison<br>**Sample size:** 152<br>**Age:** Adults<br>**Sex:** Female (n=19)<br>Male (n=133) | Survey<br>Computerized test (attention/impulsivity)<br>**Measure(s):**<br>Pittsburg Sleep Quality Index; Conner's Continuous Performance Test II; Perceived Stress Scale; Prison Adjusted Measure of Aggression; Positive and Negative Affect Schedule | **Sleep Outcomes:**<br>Sleep Disturbance<br>**Other Study Outcomes:**<br>Perceived stress, aggression, positive and negative affect, attention and impulse control, psychological distress | Yoga had positive effects on well-being and risk factors associated with recidivism (e.g. impulsivity, antisocial behavior) |
| Li | 2019 | Taiwan | Insomnia associated factors | To determine the prevalence and factors associated with insomnia among incarcerated people in a large prison in Taiwan. | Cross-sectional study | **Setting:** Prison<br>**Sample size:** 1490<br>**Age:** Adults<br>**Sex:** Males | Survey<br>**Measure(s):** Insomnia Index; Hospital Anxiety and Depression Scale (HADS), self-rated health Presence of chronic diseases and communicable diseases, smoking status | **Sleep Outcomes:**<br>Insomnia<br>**Other Study Outcomes:**<br>Anxiety, depression, self-reported health status, chronic diseases, communicable diseases | Insomnia prevalence was 26.9%, higher than the adult population in the community (5–15%), but much lower than in sampled USA female prison population (61.6%) (Dewa et al., 2017). |
| Lindberg | 2003 | Finland | Antisocial Personality Disorder, Habitual Violence | To characterize the relationship between earlier-mentioned different categorical psychiatric diagnosis describing human | Cross-sectional study | **Setting:** Psychiatric unit during incarceration<br>**Sample size:** 27<br>**Age:** Adult | Survey<br>Clinical assessment<br>**Measure(s):**<br>Polysomnography, Power | **Sleep Outcomes:**<br>Sleep Quality<br>Sleep Duration<br>Sleep Latency | Antisocial disorder persons without borderline personality disorder did not differ from healthy controls in number of awakenings. |
|  |  |  |  | impulsive aggression and sleep using polysomnography and spectral power analysis & to study the relationship between serum testosterone and sleep in persons with severe aggressive behavior. |  | **Sex:** Male | spectrum, Hormone assays, SCID structured clinical interview SCID I and II, Beck Depression Inventory Scale (BDI) | **Other Study Outcomes:** testosterone levels, Depression, Intermittent explosive disorder |  |
| Lindberg | 2003 | Finland | Antisocial Personality Disorder, Habitual Violence | To characterize the subjective and objective sleep and sleep quality in habitually violent offenders with DSM-IV diagnosis of antisocial personality disorder using a sleep questionnaire, actigraphy, polysomnography and power spectral analysis. | Cross-sectional study | **Setting:** Psychiatric unit during incarceration<br>**Sample size:** 19<br>**Age:** Adult<br>**Sex:** Male | Survey<br>Clinical assessment<br>**Measure(s):** Polysomnography, Actigraphy, Power spectral analysis, Basic Nordic Sleep Questionnaire, Sleep Diary | **Sleep Outcomes:** Sleep Quality<br>Sleep Duration<br>Sleep Latency<br>Sleep Disturbance | Antisocial personality disorder participants showed more problems in falling asleep, frequent awakenings, and greater daytime sleepiness |
| Lutz | 1990 | United States | Relaxation Training State, Anxiety, Sleep, Sickness | To study the effect of relaxation teaching on sleep, anxiety, and sick call in a prison population. | Quasi-experimental study | **Setting:** Prison<br>**Sample size:** 40<br>**Age:** Adults<br>**Sex:** Males | Survey<br>**Measure(s):** Richard Campbell Sleep Questionnaire, State Anxiety Inventory | **Sleep Outcomes:** Sleep Quality<br>**Other Study Outcomes:** Anxiety, stress, sick calls | Null hypothesis supported, but experimental group showed trend toward increased sleep and decreased anxiety. |
| Manzoor | 2021 | Pakistan | Gender, Gendered Health | To assess the gender differences in the health status of the imprisoned population of Lahore. | Cross-sectional study | **Setting:** Jail<br>**Sample size:** 320<br>**Age:** Adults<br>**Sex:** Female (n=124)<br>Male (n=196) | Survey | **Sleep Outcomes:** Insomnia<br>**Other Study Outcomes:** Physical health<br>Mental health<br>Social health | Participants assessed for depression (50%), anxiety (62.2%), mood swings (43.1%), violent behavior (19.4%), suicidal thoughts (15%), and insomnia (29.4%). Females suffered from mental health issues more than men. |
| Meijers | 2015 | Netherlands | Aggression<br>Psychosis<br>Sleep Disruption | To examine whether a relationship exists between disturbed sleep and aggression in 19 male | Cross-sectional study | **Setting:** Prison<br>**Sample size:** 19<br>**Age:** Adults<br>**Sex:** Males | Survey<br>**Measure(s):** Actigraphy | **Sleep Outcomes:** Sleep Duration | Nocturnal restlessness was a significant predictor of aggression/agitation with a large |
| | | | | incarcerated people with a psychotic illness, incarcerated in the Penitentiary Psychiatric Centre in Amsterdam, the Netherlands. | | | Social Dysfunction and Aggression Scale (SDAS) | **Other Study Outcomes:** Aggression | effect size ($R^2=.639$, adjusted $R^2=.536$). |
| Oniszczenko | 2020 | Poland | Anxiety Insomnia | To assess the relationship between anxious temperament and mood dimensions and to examine insomnia as a mediator in this relationship in male incarcerated people. | Cross-sectional study | **Setting:** Prison<br>**Sample size:** 245<br>**Age:** Adult<br>**Sex:** Male | Survey<br>**Measure(s):** Polish Adaptation of Athens Insomnia Scale (AIS); Temperament Evaluation of Memphis, Pisa, Paris, and San Diego Auto-questionnaire (TEMPS-A), Polish adaptation of UWIST Mood Adjective Checklist (UMACL) | **Sleep Outcomes:** Sleep Quality Insomnia<br>**Other Study Outcomes:** Affective Temperaments, Mood | Low energetic arousal tends to increase tense arousal, and likely anxiety, in incarcerated people. |
| Osasona | 2018 | Nigeria | Sleep Quality Factors | To assess the prevalence and associated factors of poor quality of sleep among incarcerated people in Mettu town prison, 2019. | Cross-sectional study | **Setting:** Prison<br>**Sample size:** 252<br>**Age:** Adults<br>**Sex:** Female (n=23) Male (n=229) | Survey<br>**Measure(s):** Pittsburg Sleep Quality Index; Hospital Anxiety and Depression Scale; Self-Reporting Questionnaire | **Sleep Outcomes:** Sleep Quality | 63% of respondents met PSQI criteria for poor sleep quality. Anxiety, depression, general psychopathology, age, marital status, self-reported poor physical and mental health, and self-rating of the prison food and medical services associated with sleep quality. |
| Poorebrahim | 2020 | Iran | Insomnia, Mindfulness | To investigate whether dispositional mindfulness can be beneficial in helping relieve insomnia among Iranian incarcerated people & whether psychological distress and perceived stress are potential mediators. | Prospective cohort study | **Setting:** Prison<br>**Sample size:** 208<br>**Age:** Adult<br>**Sex:** Male | Survey<br>**Measure(s):** Pittsburgh Sleep Quality Index (PSQI) Insomnia Severity Index (ISI) Hospital Anxiety and Depression Scale, 10-item Perceived Stress Scale, Mindful Attention Awareness Scale (MAAS); | **Sleep Outcomes:** Sleep Quality Sleep Disturbances Insomnia<br>**Other Study Outcomes:** Mindfulness Psychological distress Perceived stress | Greater dispositional mindfulness correlated with significantly improved sleep quality and reduced insomnia. |
| Poorebrahim | 2021 | Iran | Insomnia, Mindfulness | To test the construct validity (i.e., factor structure) of the | Cross-sectional study | **Setting:** Prison<br>**Sample size:** 361 | Survey | **Sleep Outcomes:** | The Persian Mindful Attention |
|  |  |  |  | Persian Mindful Attention Awareness Scale (MAAS) on a sample of male incarcerated people. |  | **Age:** Adults<br>**Sex:** Males | **Measure(s):**<br>Persian Mindful Attention Awareness Scale (MAAS),<br>Insomnia Severity Index (ISI),<br>General Health<br>Questionnaire (GHQ-12) | Insomnia<br>**Other Study Outcomes:**<br>Mindfulness<br>General Health | Awareness Scale is a valid to measure mindfulness in Iranian male incarcerated people |
| Raha | 2018 | India | Insomnia, Depression, Suicidality | To compare the prevalence of insomnia, depression, and suicidality between male and female incarcerated people, and to probe how the internal turmoil and the psychological burden consequent of the committed crime influences these morbid occurrences. | Cross-sectional study | **Setting:** Jail<br>**Sample size:** 80<br>Adult female (n=40)<br>Adult male (n=40) | Survey<br>**Measure(s):**<br>Diagnostic and Statistical Manual of Mental Disorders (DSM-IV-TR), Beck Depression Inventory-II (BDI-II); Pittsburgh Insomnia Rating Scale (PIRS) Columbia Suicide Severity Rating Scale (C-SSRS); Sociodemographic Variable Questionnaire | **Sleep Outcomes:**<br>Insomnia<br>**Other Study Outcomes:**<br>Depression<br>Suicidality | Depression, insomnia, suicidality significantly associated with longer conviction, convict status, presence of physical illness, substance abuse, and with higher age of their children in female incarcerated people. |
| Randall | 2019 | United Kingdom | Cognitive Behavioral Therapy Insomnia | To examine the preliminary efficacy and effectiveness of a one-shot session of cognitive behavioral therapy for insomnia (CBT-I) for incarcerated people with acute insomnia. | Quasi-experimental open trial | **Setting:** Prison<br>**Sample size:** 30<br>**Age:** Adult<br>**Sex:** Male | Survey<br>**Measure(s):**<br>Insomnia Severity Index (ISI); Patient Health Questionnaire (PHQ); Generalized Anxiety Disorder (GAD); Consensus Sleep Diary | **Sleep Outcomes:**<br>Insomnia<br>Sleep latency<br>Wake after sleep onset<br>Total sleep time<br>Sleep efficiency<br>**Other Study Outcomes:**<br>Depression<br>Anxiety | Single shot of CBT-I effective in reducing severity of insomnia, depression, and anxiety |
| Salley | 1980 | United States | REM Sleep, EEG, EEG Abnormalities, Psychopathy | To investigate REM deficit theory in psychopathy | Cross-sectional study | **Setting:** Prison<br>**Sample size:** 23<br>**Age:** Adult<br>**Sex:** Male | Psychological measure<br>Clinical sleep measures<br>**Measure(s):**<br>Rorschach Psychodiagnostic Inkblot Test, Polysomnography, EEG recordings<br>Sleep recordings | **Sleep Outcomes:**<br>REM sleep<br>REM latency<br>Total REM time<br>Slow wave sleep<br>Light sleep | No significant different sleep patterns among men diagnosed with psychopathy with normal waking EEGs, men diagnosed with psychopathy with abnormal EEGs, and men not diagnosed with psychopathy with normal EEGs. Those diagnosed with psychopathy with abnormal EEGs showed higher REM time and REM %, contradicting Hare's theory, suggesting REM sleep is unrelated to the EEG slowing observed in those diagnosed with psychopathy. |
| Sylvia | 2021 | United States | Mind-body stress management, Veterans, Veterans Health | To determine the acceptability and effectiveness of a mind-body stress reduction program for incarcerated veterans. | Quasi-experimental pilot study | **Setting:** Prison<br>**Sample size:** 24<br>**Age:** Adult<br>**Sex:** Male | Survey<br>**Measure(s):**<br>Patient Health Questionnaire-8 (PHQ-8); Perceived Stress Scale (PSS-10); General Self-Efficacy Scale (GSE); PROMIS-8B; Sleep Disturbance Scale; Cognitive and Affective Mindfulness Scale-Revised (CAMS-R); Measure of Current Status (MOCS); Current Expectancies Scale (CES); Post-program feedback survey | **Sleep Outcomes:**<br>Sleep Disturbance<br>**Other Study Outcomes:**<br>Depression, Self-efficacy, Resilience; Perceived stress, Stress reactivity, Mindfulness | 6 week mind-body stress management course improved self-efficacy, depressive symptom severity, and resilience. |
| Toler | 1978 | United States | Insomnia, Relaxation Training, Stimulus Control, Sleep Disturbance | To determine if relaxation training was an effective short-term treatment procedure for incarcerated males reporting serious sleeping disturbances and to evaluate whether additional benefits would be achieved by the use of stimulus control instructions. | Randomized control trial | **Setting:** Prison<br>**Sample size:** 27<br>**Age:** Adult<br>**Sex:** Male | Survey<br>**Measure(s):**<br>Self-reported sleep log Kincannon Mini Personality Inventory; Locus of Control Questionnaire; State-Trait Anxiety Inventory | **Sleep Outcomes:**<br>Sleep Latency<br>Sleep Disturbances<br>**Other Study Outcomes:**<br>Anxiety<br>Relaxation | Both relaxation and stimulus-control groups report decrease in the number of times night awakenings compared to the control. |
| Tussey | 2024 | United States | Mental Health, Sleep Environment, Sleep Quality | To characterize sleep quality and insomnia prevalence in women in jail in the United States. | Cross-sectional study | **Setting:** Jail<br>**Sample size:** 176<br>**Age:** Adult<br>**Sex:** Female | Survey<br>**Measure(s):**<br>Demographics<br>Life Stressors Checklist-Revised (LSC-R)<br>Posttraumatic Stress Disorder Checklist-5 (PCL-5) | **Sleep Outcomes:**<br>Sleep Quality<br>Sleep Duration<br>Sleep Latency<br>Sleep Disturbances<br>Insomnia<br>**Other Study Outcomes:** | 76% reported poor sleep quality and 70% insomnia. Depressive symptoms and PTSD significantly related to both. Excessive noise, poor bedding, and mental health cited disrupting sleep. |
|  |  |  |  |  |  |  | Center for Epidemiological Studies Depression Scale Revised (CESD-R)<br>Pittsburgh Sleep Quality Index (PSQI)<br>Insomnia Severity Index (ISI) | PTSD<br>Depression<br>Stress |  |
| Vogler | 2014 | Switzerland | Aggression, Psychosocial Adjustment | To investigate sleep quantity and quality and their association with anger-related reactions, symptoms of ADHD, depressive symptoms, physical health complaints, and life satisfaction in incarcerated men & whether good sleep hygiene in the prison context (physical exercise during the day, switching the television off at night, limiting caffeine and cigarette consumption) is related to sleep quantity and quality. | Cross-sectional study | **Setting:** Prison<br>**Sample size:** 49<br>**Age:** Adult<br>**Sex:** Males | Survey<br>**Measure(s):**<br>Insomnia Severity Index (ISI)<br>Sleep hygiene checklist<br>Self-reported sleep duration<br>Anger-Related Reactions and Goals Inventory (ARGI)<br>ADHD Self-Report Scale (ADHD-SR)<br>Patient Health Questionnaire (PHQ-15, German version, 13 items)<br>Allgemeine Depressionsskala – short form (ADS-K)<br>Berne Questionnaire of Subjective Well-Being (5 items) | **Sleep Outcomes:**<br>Sleep Quality<br>Sleep Duration<br>Sleep hygiene<br>Insomnia<br>**Other Study Outcomes:**<br>Aggression<br>Rumination<br>ADHD symptoms<br>Physical health complaints<br>Depression<br>Life satisfaction | Sleep duration of 6hr or less correlated with aggressive reactivity and physical health complaints. Poor sleep quality marked higher levels of aggressive reactivity, ADHD symptoms rumination, and physical health complaints. |
| Wadey | 2009 | United Kingdom | Valerian Tea bags, Subjective Sleep Quality | To establish whether Valerian teabags improve subjective sleep quality for men with sleep disturbance and dual diagnosis in a remand prison setting within a 14-day period. | Randomized control trial | **Setting:** Prison<br>**Sample size:** 15<br>**Age:** Adult<br>**Sex:** Males | Survey<br>**Measure(s):**<br>Pittsburgh Sleep Quality Index; General Health Questionnaire; Functional Analysis of Mental Health and Addiction | **Sleep Outcomes:**<br>Sleep Quality<br>Sleep Duration<br>Insomnia<br>**Other Study Outcomes:**<br>Psychological wellbeing<br>Functioning | Although teabags may be helpful for certain individuals, results between control and experimental groups were not statistically significant. |
| Woolhouse | 2018 | New Zealand | Traumatic Brain Injury | To study the prevalence of Traumatic Brain Injury (TBI) and mental health issues in an exclusively female prison population in New Zealand. | Cross-sectional study | **Setting:** Prison<br>**Sample size:** 38<br>**Age:** Adolescent, Adult<br>**Sex:** Female | Survey<br>**Measure(s):**<br>Ohio State University TBI Identification Method (OSU-TBI-ID), | **Sleep Outcomes:**<br>Sleep quality<br>**Other Study Outcomes:** | Traumatic brain injury is highly prevalent among female offenders and may be linked to increased mental health problems |
|  |  |  |  |  |  |  | Pittsburg Sleep Quality Index (PSQI), Depression, Anxiety and Stress Scale (DASS-21), Achenbach System of Empirically Based Assessment (ASEBA-Adult Behavior Checklist), Wechsler Abbreviated Scale of Intelligence (WASI) | TBI, Depression, Anxiety, Stress, Behavioral health, Cognitive functioning |  |
| Zar | 2017 | Iran | Exercise, General Health, Sleep Quality | To investigate the Effectiveness of exercise activities on the sleep quality and the general health of incarcerated people. | Cross-sectional study | **Setting:** Prison<br>**Sample size:** 97<br>**Age:** Adult<br>**Sex:** Male | Survey<br>**Measure(s):**<br>Pittsburg Sleep Quality Index; General Health Questionnaire (GHQ_28); Goldberg & Hild's General Health Questionnaire, U Mann Whitest Test | **Sleep Outcomes:**<br>Sleep latency<br>Sleep duration<br>Habitual sleep efficiency<br>Sleep disturbances<br>Use of sleep medication, Sleep quality, Daytime dysfunction<br>**Other Study Outcomes:**<br>TBI, Depression, Anxiety, Stress, Behavioral health, Cognitive functioning | Active incarcerated people showed significant improvement in general health, physical symptoms, anxiety, depression |
| Zhang | 2022 | China | Short-Term Mindfulness Intervention, Stress | To test the effects of two different 4-day interventions (i.e., instructor-guided and audio-based) with content similar to Mindfulness-Based Cognitive Therapy for newly incarcerated males, and to compare the effectiveness of the two interventions relative | Randomized control trial | **Setting:** Prison<br>**Sample size:** 90<br>**Age:** Adult<br>**Sex:** Male | Survey<br>**Measure(s):**<br>Insomnia Severity Index; 3-item Daily Stress Questionnaire, Negative Affectivity (NA) subscale in the Positive and Negative Affect Schedule | **Sleep Outcomes:**<br>Insomnia<br>**Other Study Outcomes:**<br>Stress<br>Negative affect<br>Mindfulness | A very brief, 4-day mindfulness based intervention reduced perceived stress during early incarceration. |
|  |  |  |  | to a no-intervention control group. |  |  | Daily mindfulness practice logs |  |  |
| Zhang | 2023 | China | Anxiety, COVID-19, Mood Disorders | To compare the anxiety levels in incarcerated people before and after the COVID-19 outbreak and analyzed the causes of the changes in anxiety. | Longitudinal cohort study | **Setting:** Prison<br>**Sample size:** 803<br>**Age:** Adult<br>**Sex:** Male | Survey<br>**Measure(s):**<br>Insomnia Severity Index; Generalized Anxiety Disorder-7(GAD-7); Patient Health Questionnaire-9<br>COVID-19 questions | **Sleep Outcomes:**<br>Insomnia<br>**Other Study Outcomes:**<br>Anxiety<br>Depression<br>COVID-19 | Prevalence of anxiety among incarcerated people decreased 4 months into the COVID-19 pandemic. |

